# COVID-19 Vaccine Safety Studies among Vulnerable Populations: A Systematic Review and Meta-analysis of 120 Observational Studies and Randomized Clinical Trials

**DOI:** 10.1101/2024.12.27.24319417

**Authors:** Sima Mohammadi, Malede Mequanent Sisay, Putri Widi Saraswati, Alhadi Khogali Osman, Nicolaas.P.A Zuithoff, Daniel Weibel, Miriam Sturkenboom, Fariba Ahmadizar

## Abstract

**BACKGROUND:** The COVID-19 vaccines were rapidly developed and tested, but concerns about vaccine-related adverse events remain, especially in vulnerable groups like pregnant women, children, and those with certain health conditions. This review aims to summarize rates of such adverse events in individuals often not included in randomized clinical trials (RCT).

**METHOD:** From December 2019 to February 2022, we searched Embase and Medline for observational studies and RCTs on adverse events post-COVID-19 vaccination in vulnerable groups. We examined serious and non-serious events in individuals with specific medical conditions, infants, children, pregnant individuals, and socioeconomically disadvantaged individuals. Cumulative risks for all events were calculated. The Incidence rate (IR) and 95% confidence intervals were reported for those studies that met the follow-up period criteria based on the referenced literature. For events with data on exposed and unexposed groups, we calculated the odds ratio. Pooled incidence rates were calculated per 1000 person-days using a random-effects model. Sub-group analyses were conducted based on vaccine types and doses, with heterogeneity assessed using I^2^.

**FINDINGS:** Of the 4,254 papers, 235 met eligibility criteria, including 120 studies with 171,073 participants (113 observational, eight RCTs. We examined 17 severe and 7 non-severe adverse event categories. Lymphadenopathy (IR: 1.95[1.20;3.19]), autoimmune disease and multiple sclerosis flare-up (1.13 [0.47;2.68]), and cardiac symptoms (0.26[0.00;10.58]) were the most severe events. Allergic reactions were more common among autoimmune (7.03[4.10;12.06]) and cancer (4.87[2.21;10.76]) groups. vaccinees who received the second dose of vaccine had higher proportions of disease flare-ups (39.27 [18.08;85.31] vs 22.13 [10.22; 47.93]); cardiac symptoms (6.11[2.05-18.22] vs 3.78[2.53-5.65]); and cardiac events (5.34[1.69;16.90] vs 5.05[1.40,18.19]) in observational studies.

**INTERPRETATION:** This review highlights COVID-19 vaccine safety in vulnerable populations, enhancing vaccination strategies. Further real-world research is needed to validate and extend our findings, especially in addressing safety gaps among vulnerable groups.

## Introduction

The rapid development and global deployment of COVID-19 vaccines have been pivotal in the ongoing battle against the pandemic. At the start of the vaccination rollout, the WHO prioritizes the vaccination of populations at high risk of severe COVID-19 to ensure the protection of vulnerable individuals, such as pregnant and lactating women, children, those with underlying health conditions like immunocompromised and autoimmune diseases, who are more likely to experience severe complications from COVID-19^1^. While pivotal clinical trials and randomized clinical trials (RCTs) provided crucial safety data for vaccine authorization, including these vulnerable groups was limited, necessitating further investigation into the safety profiles of COVID-19 vaccines within these populations^2,3^.

This review addresses this gap by comprehensively analysing adverse events of special interest (AESIs) following COVID-19 vaccination, focusing specifically on vulnerable populations. We determined the incidence rate (IR) and prevalence of COVID-19 vaccine-related AESIs within the overall vulnerable population. Additionally, we conducted subgroup analyses for specific groups, such as individuals with particular medical conditions, infants, children, and pregnant individuals, and performed analyses based on vaccine type and dose. These efforts aim to inform vaccination strategies and enhance safety monitoring.

## Method

### Search strategy and selection criteria

This study is a systematic review and meta-analysis following the Preferred Reporting Items for Systematic Reviews (PRISMA) requirements. A comprehensive compilation of keywords utilized in each database (Tables S1-2). Embase and Medline databases were established for our study from 14^th^ December 2019 until 21^st^ February 2022. To minimize the possibility of missing relevant literature, all titles and abstracts underwent rigorous independent reviews by three reviewers (A.K.O, P.W.S, M.M.S) using the Rayyan software^4^. Disagreement between reviewers was resolved through the review process involving the fourth and fifth researchers (S.M, F.A).

We included all observational studies (cohort, cross-sectional, and case-control studies) and pivotal clinical trials/RCTs in Phase III/IV, as of December 14th, 2021, that reported AESIs or reactogenicity associated with COVID-19 vaccinations within a specified time window after vaccination. We included all severe and non-severe adverse events associated with the COVID-19 vaccine. Severe adverse events entail those causing considerable harm, requiring medical attention or hospitalization, including systemic allergic reactions, cardiac or neurological symptoms, respiratory symptoms, lymphadenopathy, maternal or neonatal events, and death. Non-severe adverse events include mild effects that do not harm significantly, such as arthralgia, myalgia, fever, fatigue, and gastrointestinal symptoms. We excluded all studies that did not report rates of adverse events, non-English papers, literature, editorial publications, systematic reviews, animal studies, clinical trials in phase I or II, and heterologous vaccine studies.

Our study’s inclusion criteria for the target population encompassed immunocompromised individuals (people with HIV, solid organ transplant recipients, hematopoietic cell transplant recipients, and individuals receiving immunosuppressive therapies), cancer (individuals with any malignancy), pregnant and breastfeeding, children under 12 years old and adolescents aged 12-17 years, individuals living with disabilities (such as Down syndrome), migrants (including refugees, internally displaced persons, asylum seekers, undocumented migrants and transgenders).

### Data extraction

Four researchers (S.M, M.M.S, P.W.S, A.K.O) retrieved data, assessed the studies’ quality, and verified them. An independent investigator confirmed the accuracy of the extracted data (S.M.). We extracted numerical data and information on the subgroups. Study features included study design, sample size, number of vaccinated, number of events in vaccinated, population type, and characteristics (sex, age, country), vaccine platform, dose and brand, adverse event, and follow-up period (days). Vaccines’ outcomes were classified according to their organ/system involvement. We selected 24 main outcomes for reporting based on the clinical impact of the outcomes and available studies. We grouped various terms, synonyms, and closely related outcomes under broader specific categories (Table S3). When multiple outcomes from the same class were presented, we included all of them.

### Quality assessment

The New Castle Ottawa checklist and ROBIN-I tool were utilized to evaluate the risk of bias for observational and trials, respectively^5,6^. Two reviewers (M.M.S, P.W.S) did the risk of bias assessment, verified by a third reviewer (S.M).

### Data analysis

We used descriptive statistics to characterize and summarize the included studies. To mitigate the impact of varying follow-up durations across the studies and maintain consistency in follow-up duration for accurate comparison and pooling of data, we assessed each study’s follow-up period against predefined criteria derived from referenced guidelines and literature^7–17^ (Table S4). Studies meeting the follow-up period criteria based on referenced literature were included in calculating incidence rates. We calculated cumulative risks (incidence proportion) for all events. For events with available data on exposed and unexposed groups, we calculated the odds ratio (OR) with a 95% CI.

Pooled IRs and 95% CI were calculated on the scale of 1000 person days, and risk was calculated per 1000 observations. We employed the inverse variance method to weigh the effect sizes of the studies based on their respective variances. We used a Random-Effects Model because all studies included in our analysis did not represent a homogeneous population. This model accounts for variations between studies that arise from factors beyond simple sampling error, allowing us to incorporate and address the diversity in study designs, populations, and settings. We used the I^2^ statistic to assess heterogeneity among studies, with values indicating no (0%), low (25%), moderate (50%), or high (75% or more) heterogeneity. Rates or risks without an associated I^2^ value indicate no meta-analysis was conducted for these outcomes. Instead, these values represent descriptive findings from individual studies. We explored asymmetry and P value to assess the publication bias using a funnel plot and an Egger’s test. We initially performed analysis to estimate the cumulative risk in the overall vulnerable population for each adverse event and subsequently conducted subgroup analyses based on vaccine dose, vaccine type, and specific study populations. Pooled effects were reported for outcomes with data from at least two studies or individual studies where data could be pooled meta-analytically. We conducted all meta-analyses using the “Meta” package in R Studio 3.6.0 software.

## Results

Following the PRISMA guidelines, we initially retrieved 4,763 articles. After deduplication, 4,254 unique articles underwent screening, 3,957 were excluded based on title and abstract, and 62 were not retrievable due to the unavailability of full-text access. Among the 235 full-text screened studies, 11 were excluded due to mismatch with the target population, three had incorrect exposure, 14 had incorrect outcome, 27 had wrong study design, 28 had wrong publication type, 14 had incomplete data, and two were non-English languages. A total of 136 studies remained eligible for inclusion in the review (Figure 1). Among these, 120 studies^3,18–135^ were included in the meta-analysis. Table S5 provides further details on the descriptive characteristics of the studies included in the analysis.

**Figure 1.**
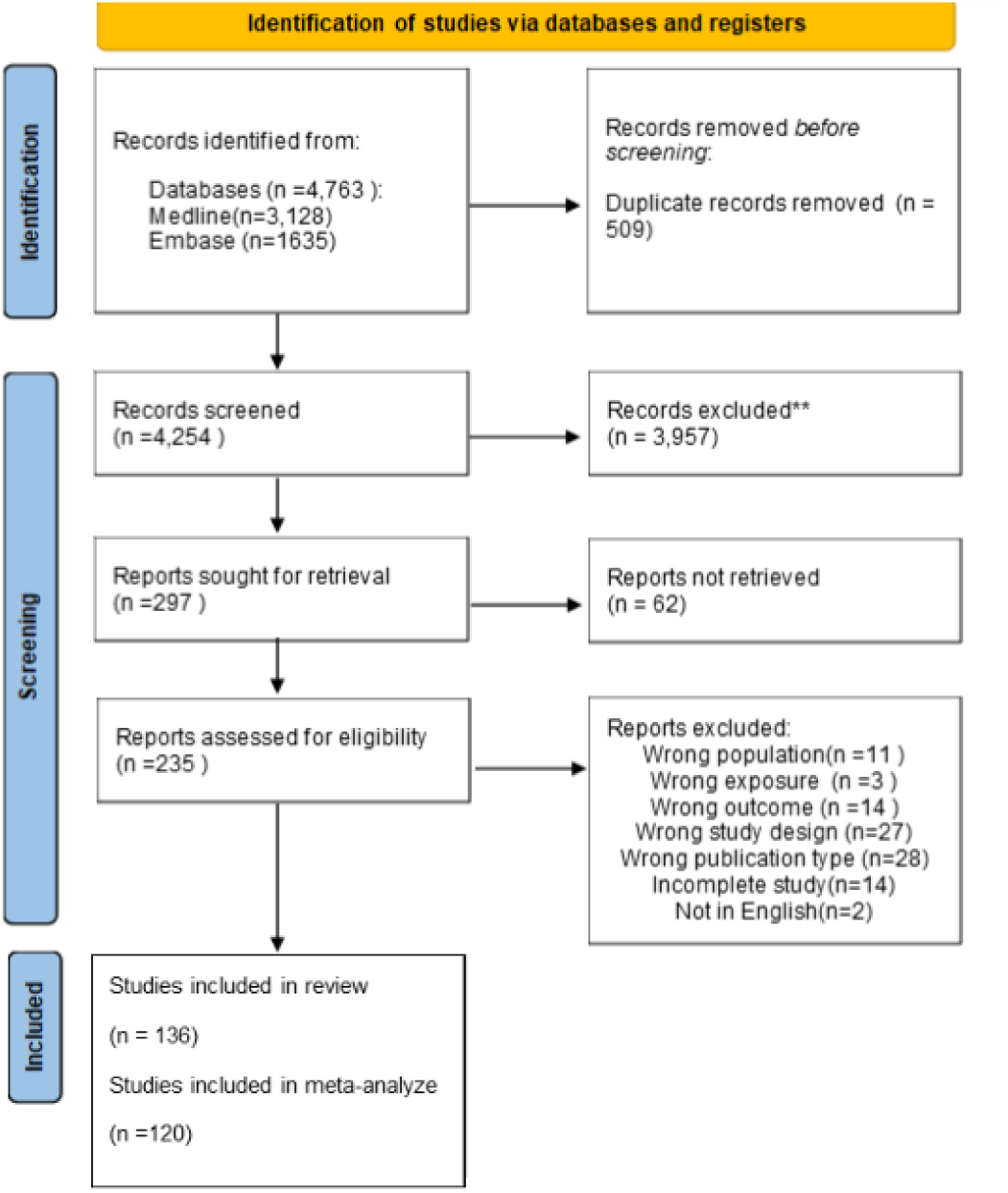
PRISMA.

These studies included populations from over 33 countries/regions (Figure S1). The predominant study design (67.8 %) was cohort studies, followed by (25%) cross-sectional studies, (6.6%) RCTs, and (1.6%) case-control studies.

Using the Newcastle Ottawa scale, among observational studies, 112(28.8%) received a good quality assessment score, while 36% were rated as fair quality and 35.2 as poor quality. Using the ROBIN-I tool, 66.7 of RCTs were rated as having a low risk of bias, 8.3% had some concerns and 25% had a high risk of bias.

Among observational studies, 55.8% reported data for dose 1, 27.5% on dose 2, and 2.5% on doses 3, while 28.3% were unspecified for the vaccine dose. Most studies (64.1%) provided data on mRNA vaccines, while only 10.8% focused on inactivated vaccines and 5% on viral vector vaccines. The remaining studies did not specify the data by vaccine type. A total of 24.1% of studies included populations with auto-immune disease, 24.1% cancer patients, 20% immunocompromised, 13.3% pregnant, 7.5 % children and adolescents, 5% multiple sclerosis (MS), 4.1% lactating and 1.6% Down and Dravet syndrome (Table S6).

### Allergic reaction

In both cohorts and cross-sectional studies, most allergic reactions occurred among individuals with autoimmune diseases with a pooled proportion of (cohort:7.03[4.10;12.06], I^2^=0.00^68,77,123^; cross-sectional: 9.93[4.77;18.19]^47^) followed by cancer individuals (cohort: 2.68,[1.01;10.00], I^2^=0.00^109,112^; cross-sectional: 2.91[0.48;17.67], I^2^=0.00)^32^. The cohort study revealed that patients with cancer exhibited a higher pooled proportion of allergic reactions after the second vaccine dose (4.87[2.21; 10.76], I^2^= 0.00)^67,104,109,112^ compared to the first dose (2.68[1.01;10.00], I^2^= 0.00)^109,112^. Additionally, individuals with autoimmune diseases demonstrated a higher pooled proportion of allergic reactions after the first dose compared to the second dose respectively, in both cohorts (7.03[4.10;12.06], I^2^ =0.00^68,77,123^ vs 5.53[3.60;8.48], I^2^ =0.00^66,68,77,118,123^) and the individual cross-sectional studies (9.93[4.77;18.19] vs 8.94 [4.09;16.90])^47^ (Figure 2 and Table S7).

**Figure 2.**
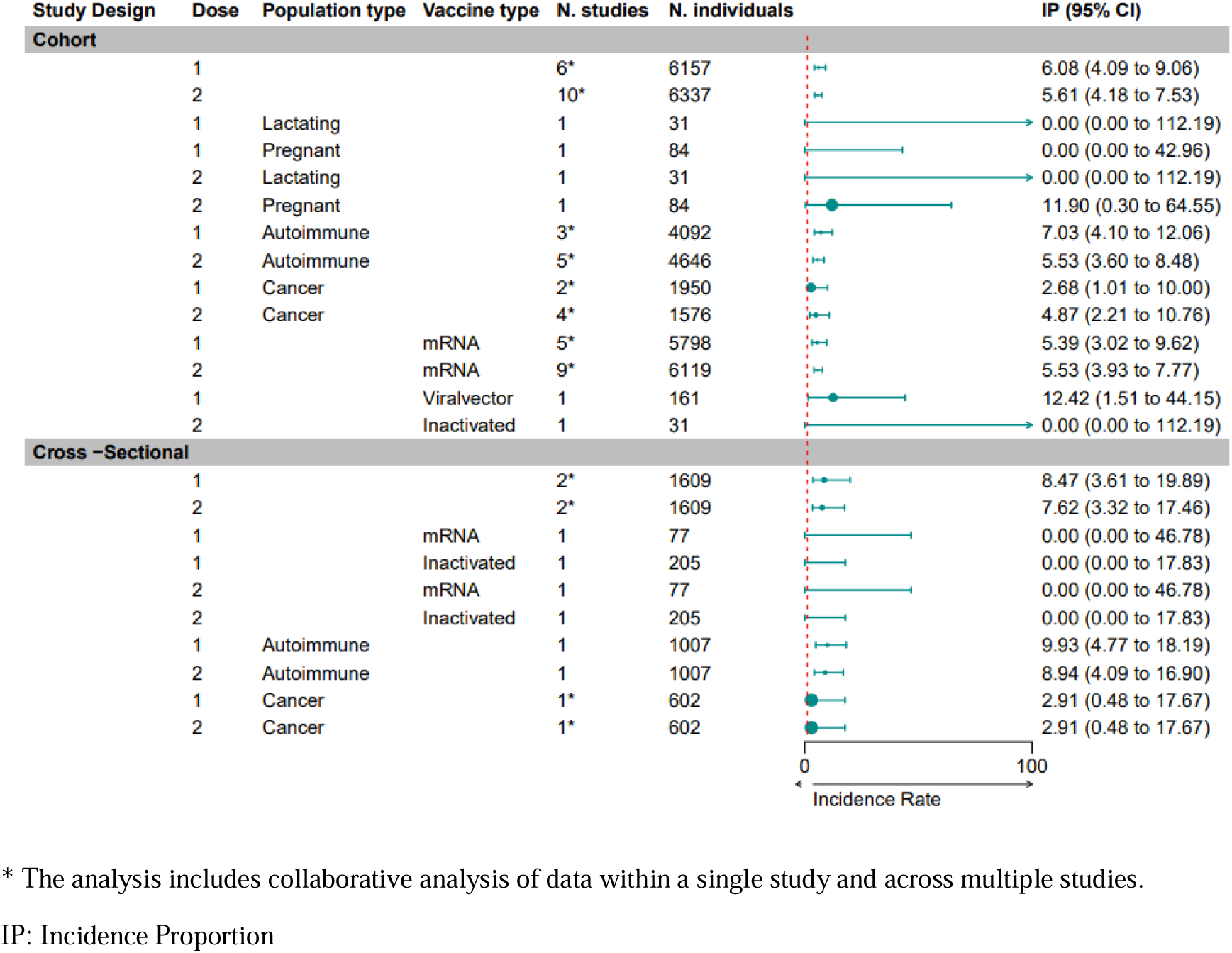
Allergic Reaction Pooled Incidence Proportion.

### Accelerated allergic reactions

Three studies assessed accelerated allergic reactions. The data analysis from a study on autoimmune patients, unspecified for dose, showed an incidence rate of 1.82[0.81-2.83], I^2^=0.00^49^. According to an individual cross-sectional study on MS patients^31^ and one on lactating women^46^, the IR of allergic reaction within 48 hours is higher in MS than in lactating women (27.81[15.62;40.00] vs 1.28[0.00;16.39]). Accelerated allergic reaction IR is higher in dose 1 than dose 2 in an individual cross-sectional study of 720 MS patients. (27.81[15.62;40.00] vs 13.91[5.29;22.53]) ^31^. There were no significant differences in the pooled IR between vaccination types (mRNA:1.59[0.00;10.21], I^2^=0.00 vs viral vector: 1.66 [0.00;19.72], I^2^=0.00)^49^ (Figure 3).

**Figure 3.**
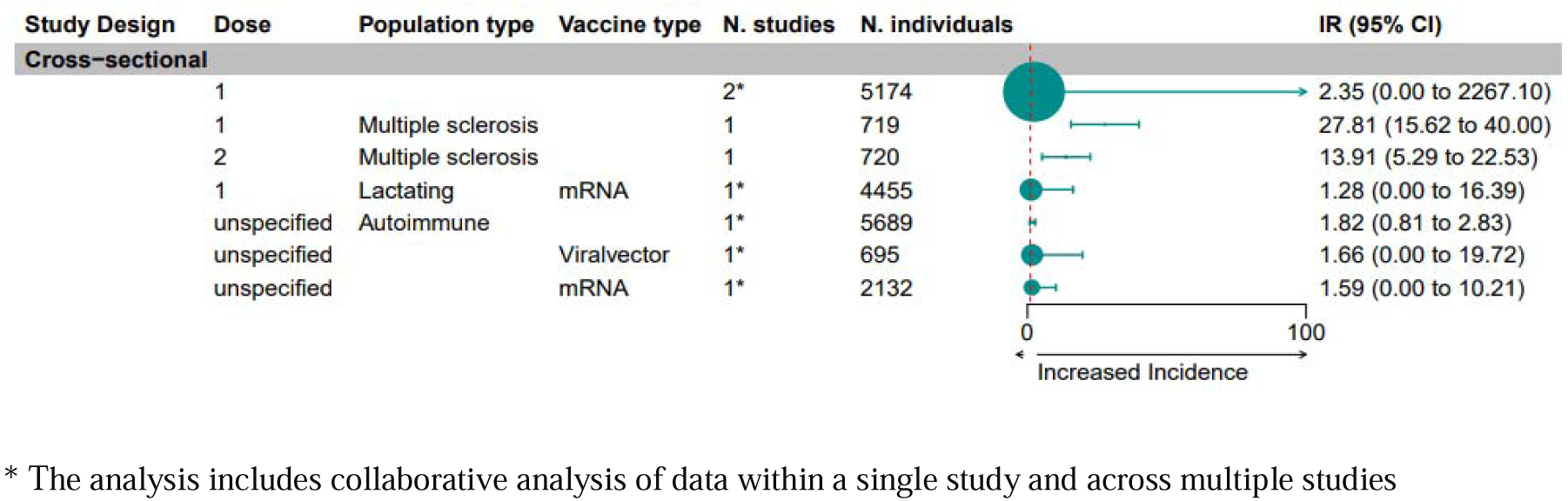
Accelerated Allergic Reaction Pooled Incidence Rate.

### Cardiac event

A prospective, multicenter RCT revealed that the IR of myocardial infarction is 0.13 (0.03;0.53)^21^. The IR of cardiac events in a big population-based cohort of 2,494,282 children and adolescents vaccinated with mRNA is higher after the second dose (0.01[0.01;0.01], vs first dose 0.0024[0.0004;0.0043])^61^ (Table S7). The pooled IR of two cohorts on autoimmune patients represent the IR of 0.02[0.00;4.14], I^2^=0.95 for myocarditis and pericarditis^80,97^, and the pooled IR in cross-sectional studies on autoimmune patients revealed that mRNA vaccines had greater IR of the cardiac event than viral vector; (viral vector dose 1: 0.00[0.00;0.00], I^2^=0.00% vs mRNA dose 1: 1.19[0.00;0.56], I^2^=0.00%; viral vector dose 2: 0.00[0.00;80.42], I^2^=0.00 vs. mRNA dose 2: 0.36[0.00;0.78], I^2^=0.00)^38^ (Figure 4)

**Figure 4.**
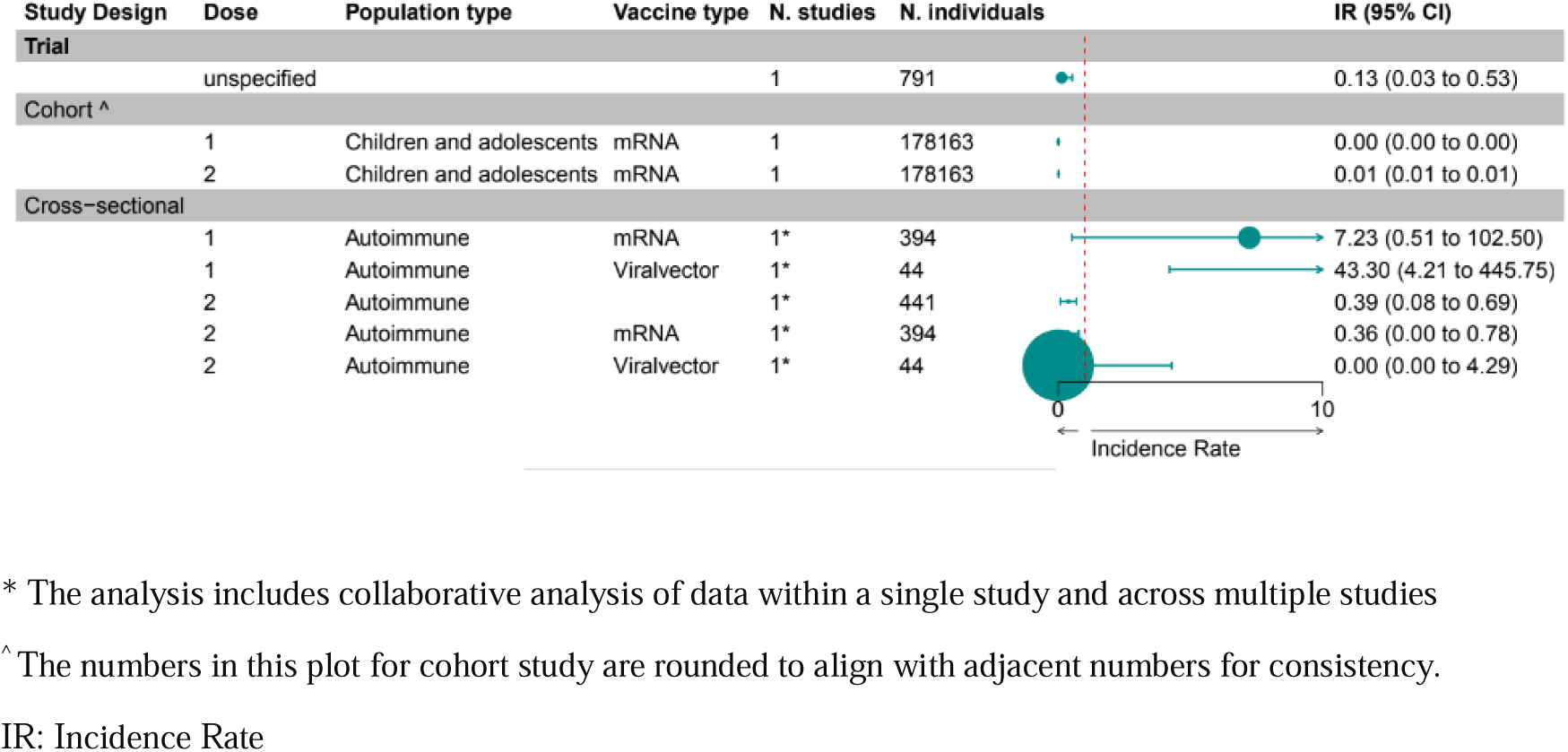
Cardiac Event Pooled Incidence Rate.

### Cardiac symptoms

Similar to cardiac events, the cardiac symptoms such as chest pain and palpitation, have a higher pooled IR following dose 2 (0.75[0.00;5175.62], I^2^=0.00)^85,121^ compared to dose 1 (0.26[0.00;10.58], I^2^=43%)^68,85,121^ in the overall pooled analysis of cohort studies. Similarly, a cross-sectional study on children vaccinated with an mRNA vaccine supports this finding, reporting higher cardiac symptom rates after dose 2 (277.11 [170.58; 283.64]) compared to dose 1 (184.56 [135.78; 233.34])^26^ (Figure 5).

**Figure 5.**
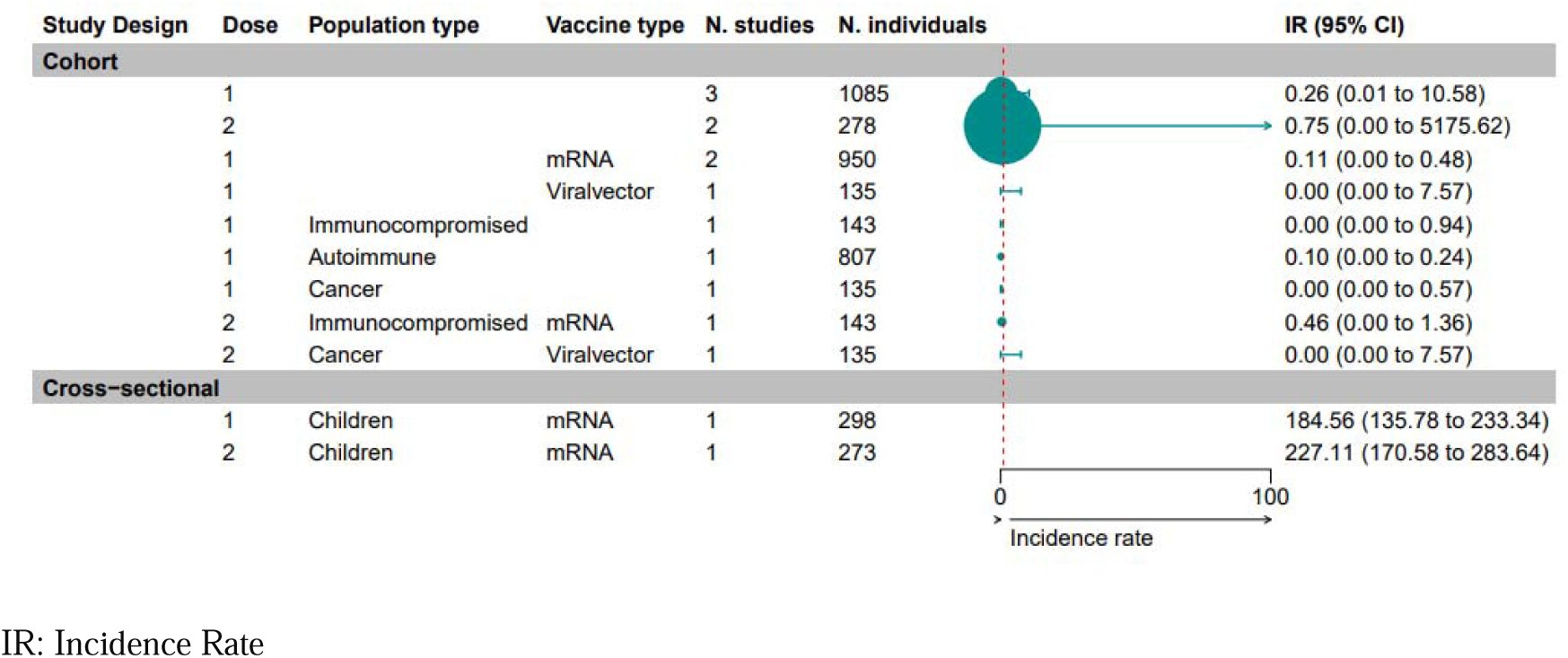
Cardiac Symptoms Pooled Incidence Rate.

### Disease Deterioration/Flare/Recurrence

In cohort studies, the pooled IR of disease deterioration/flare /recurrence in the MS population is 0.79[0.00;2.40], I^2^=0.00 for vaccine dose 1^55^. The pooled IRs comparing doses 1 and 2 revealed that other autoimmune diseases experience more flares than the MS Population. (dose1: 0.92[0.00;2.01], I^2^=33%; vs 0.79[0.00;2.40], I^2^=0.00)^68,77,123^ and; (dose2: 2.73[0.56;4.89], I^2^=97% vs 1.42 [0.00;12.55], I^2^=90%)^68,77,118,123^. Based on the findings of cohort studies, there is a higher pooled IR of disease recurrence/flare/deterioration after receiving the second dose (2.40[0.81;3.99], I2=96%)^55,68,77,118,123^of the mRNA vaccine than after the first dose; (0.85[0.36;1.34], I^2^=18%)^55,68,77,123^. However, the pooled effect from the analysis of autoimmune patients vaccinated with mRNA vaccine in cross-sectional studies showed the opposite trend (dose 1: 4.02[0.00;8.92], I^2^=0.00; vs dose 2: 1.61[0.42;2.80])^39^ (Figure 6).

**Figure 6.**
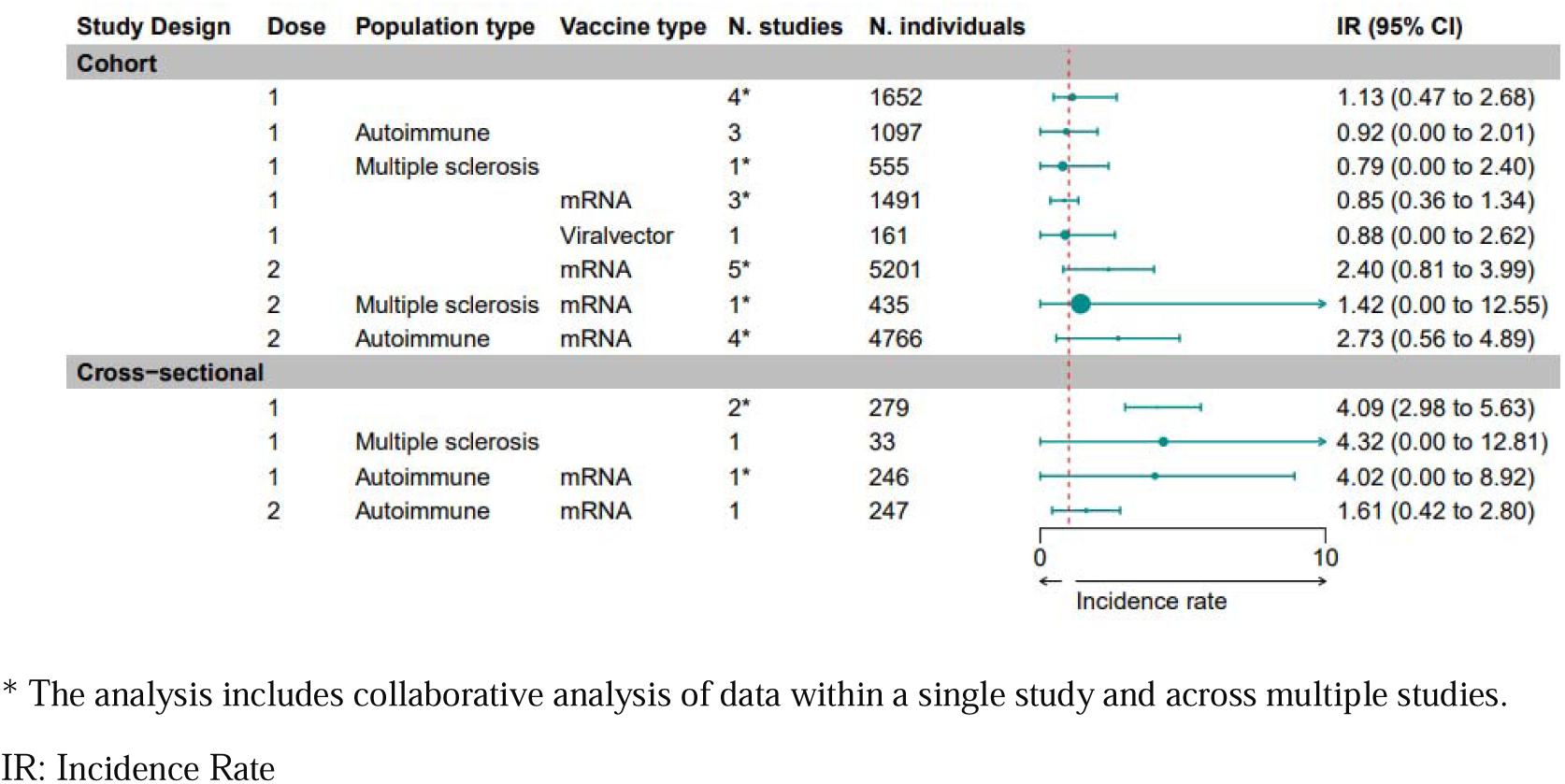
Disease Flare-up Pooled Incidence Rate.

### Respiratory symptoms

The analysis of two RCTs on autoimmune vaccinated with inactivated vaccine following dose 1 represented the pooled IR of 2.15 [0.01;4.29], I^2^=56% for respiratory symptoms^22,23^. Pooled IR of respiratory symptoms in the overall vulnerable population is higher in dose 2 compared to the dose, in both cohorts (1.73[0.28;10.41], I^2^=0.00^85,119^ vs 0.06[0.00;0.19], I^2^=0.00^68,85,119^) and cross-sectional (5.83[0.21;158.58], I^2^=94% vs 4.13[0.02;712.07], I^2^=94%)^26,38^ studies. The analysis of cross-sectional studies exhibits a greater IR of respiratory symptoms in mRNA vaccinated compared to the viral vector vaccinated; (mRNA, dose1: 60.58[0.00;320.09], I^2^=96%^26,38^, vs viral vector dose1: 1.20[0.00;14.60)], I^2^=0.00^38^); (mRNA, dose2: 74.52[0.00; 394.91], I^2^=97%^26,38^, vs viral vector dose2: 0.00 [0.00; 4.29])^38^ (Figure 7).

**Figure 7.**
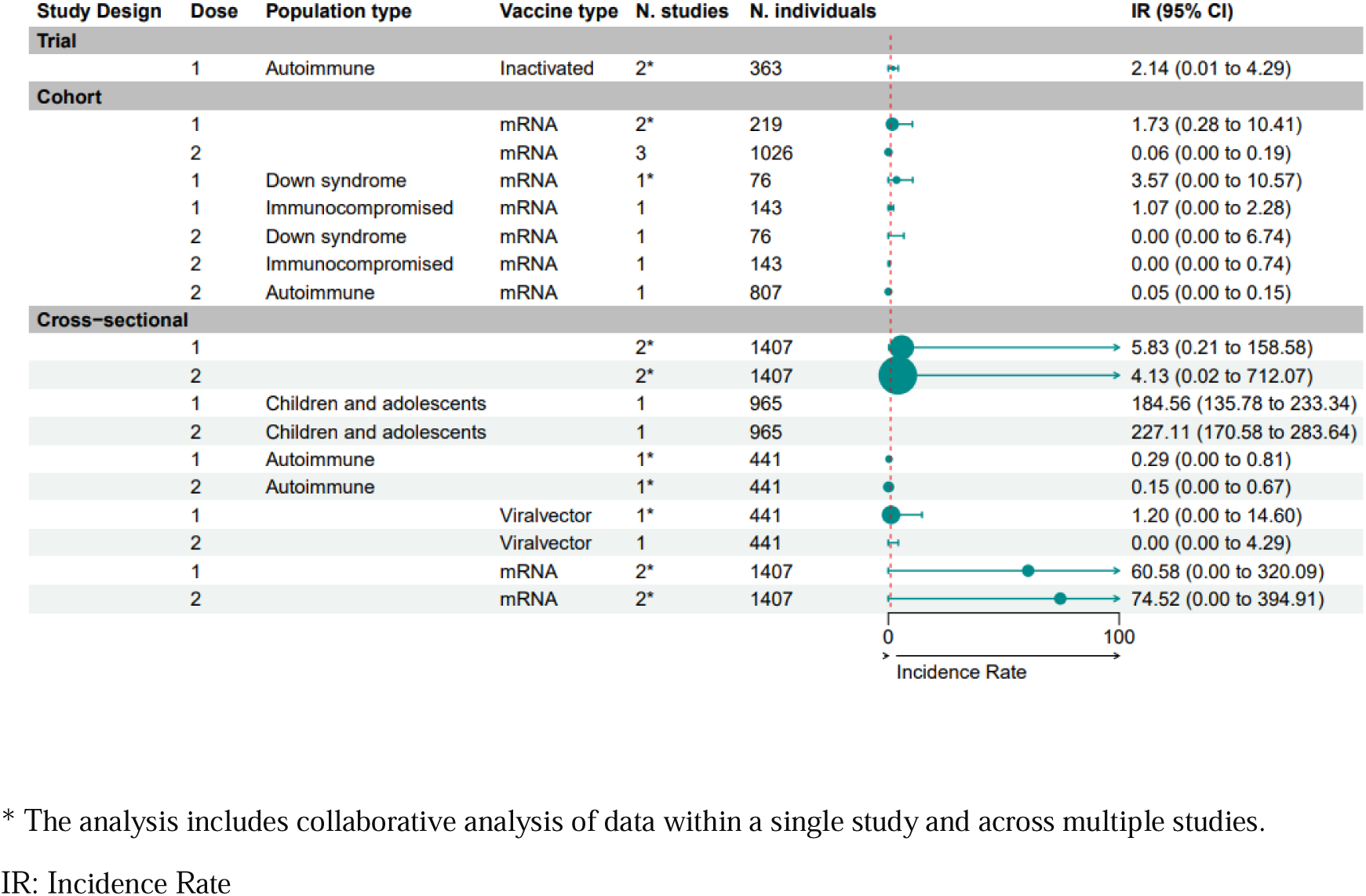
Respiratory Symptoms Pooled Incidence Rate.

### Hospitalization

Two separate cohort studies, one involving 90 individuals with autoimmune diseases^77^ and another encompassing approximately 444,000 children and adolescents^80^, found that the proportion of hospitalization is notably higher among autoimmune patients in comparison to adolescents who have received mRNA vaccines. (dose1: 22.22[2.70;77.98] vs 0.08[0.06;0.11]); (dose2: 11.11[0.28;60.36] vs 0.16[0.13;0.20]). In both cohort and cross-sectional studies, adolescents who received the mRNA vaccine experienced double the number of reported hospitalizations after the second dose compared to the first dose (cohort dose1: 0.08[0.05;0.11] vs dose 2: 0.16[0.13-0.20])^80^; (cross-sectional dose1: 53.69[31.00,85.73], dose 2: 120.88[84.69;165.55])^26^, (Figure 8 and Table S7).

**Figure 8.**
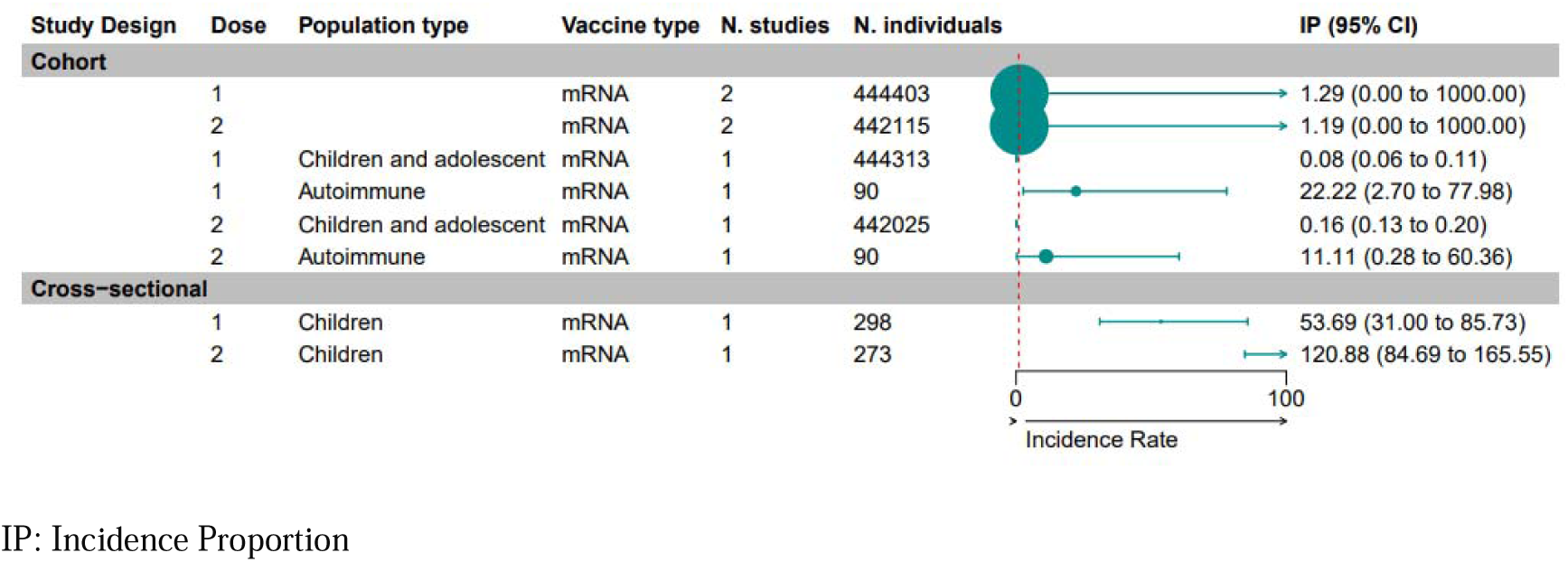
Hospitalization Pooled Incidence Proportion.

### Lymphadenopathy

The pooled IR of lymphadenopathy following dose 2 is higher than the first dose in the cohort, cross-sectional, and case-control studies (cohort of cancer patients: 1.80[0.00;3.67], I^2^=0.00^93,116^; vs 0.77[0.00;1.66], I^2^=0.00^93,110,116^); (cross-sectional of autoimmune patients: 0.57[0.00;1.43], I^2^=0.00 vs 0.15[0.00;0.54], I^2^=0.00)^38^; individual case-control of pregnant women: 2.93[0.90;4.96] vs 0.37[0.00;1.08])^134^ (Figure 9 and Table S7).

**Figure 9.**
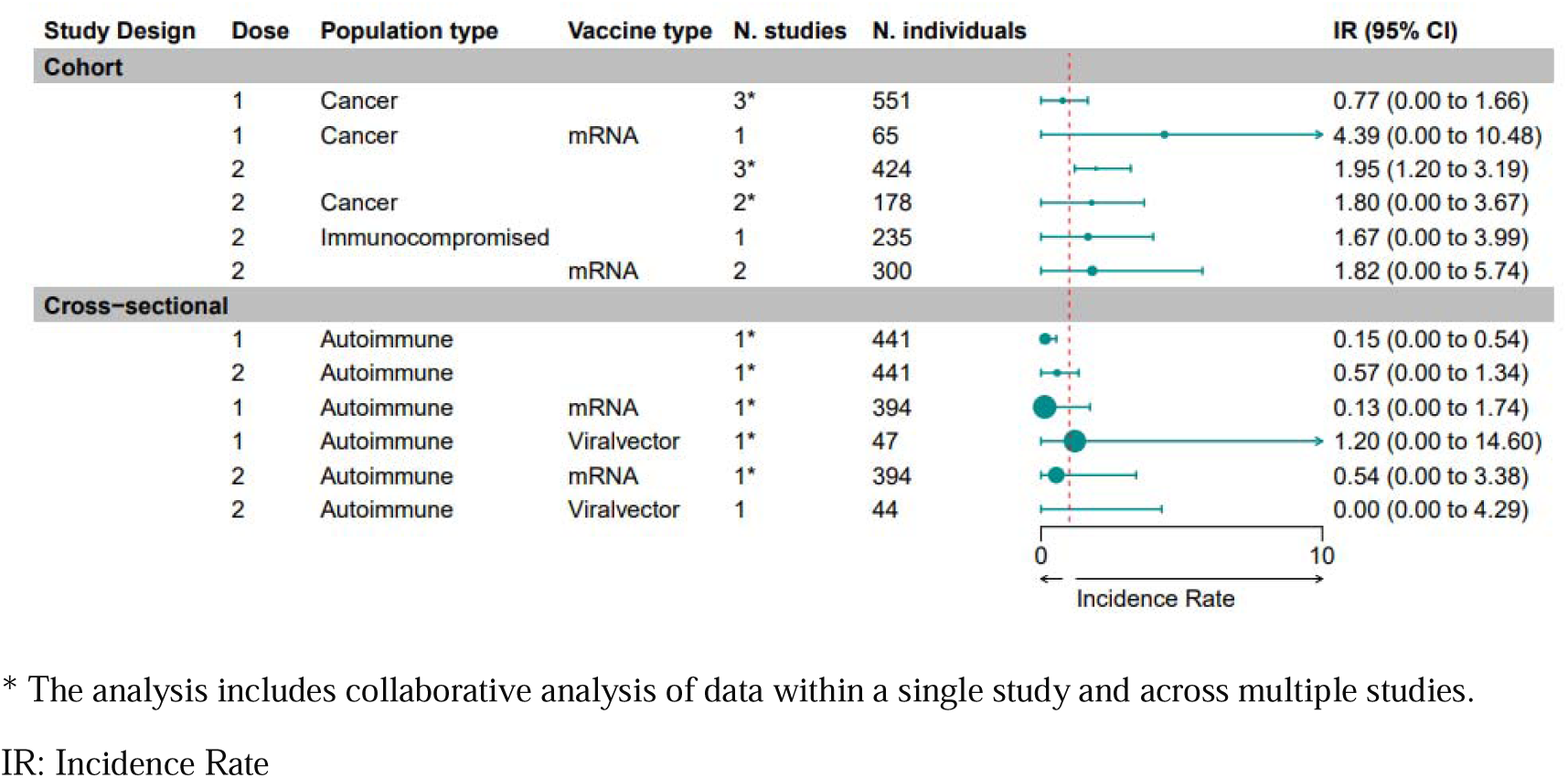
Lymphadenopathy Pooled Incidence Rate.

### Hypertensive disorder

There was no difference in the pooled proportion of hypertensive disorder observed across different doses in immunocompromised patients in cohort studies (dose 1: 16.39[0.41;87.99]^92^ vs dose 2: 18.71[8.84;39.59], I^2^=0.00^67,92^ vs dose 3: 16.39[0.04;116.38])^92^. The pooled proportion of hypertensive disorder among organ transplant recipients after the second dose was higher compared to that among individuals with autoimmune diseases (18.71[8.84;39.59], I^2^=0.00^67,92^; vs 3.31[0.40;11.89]^118^). An individual cohort study on transplant recipient reported no cases of hypertension after receiving the inactivated vaccine^67^. All studies reporting this outcome were conducted among mRNA-vaccinated individuals^67,92,118^ (Figure 10).

**Figure 10.**
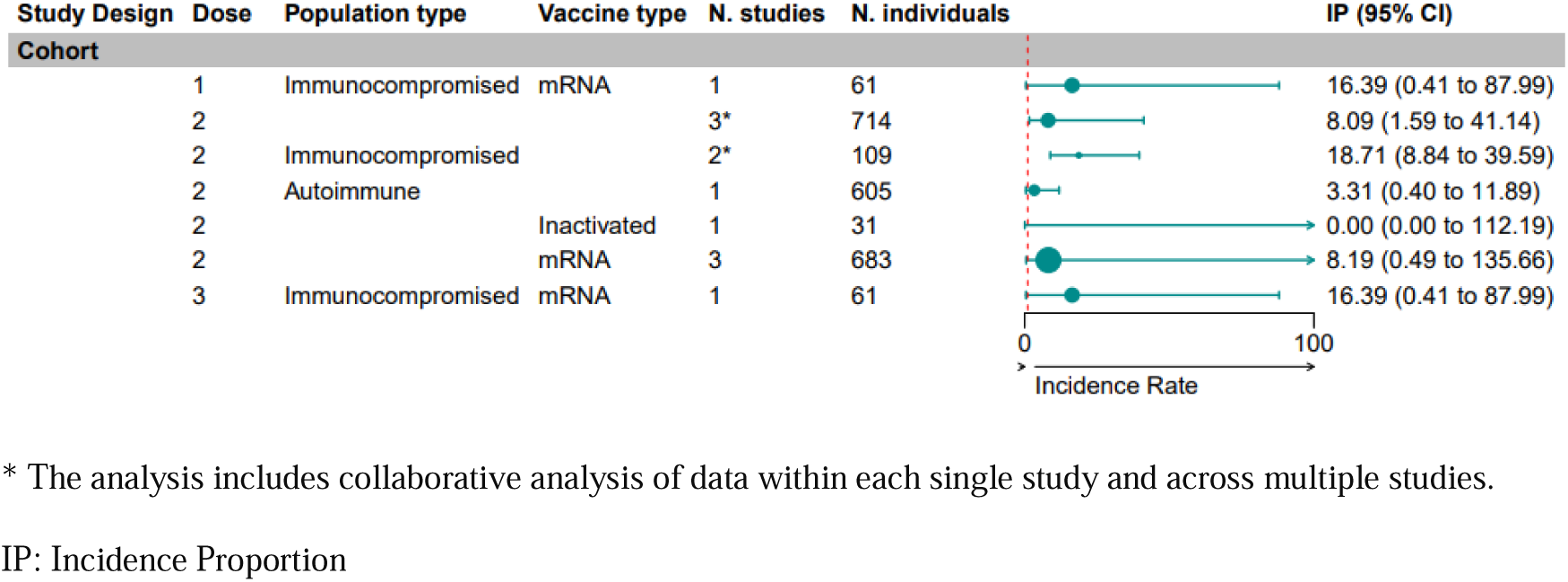
Hypertensive Disorder Pooled Incidence Proportion.

### Neurological symptoms

Analysis of cohort and case-control studies separately indicates that the pooled proportion of neurological symptoms including Paresthesia, numbness, facial tingling, tremor, and tinnitus per 1000 observations in overall vulnerable population is higher after the second dose compared to the first dose (cohort: 16.43[2.32;116.19], I^2^=15%^55,67^; vs 10.83[0.00;1000.00], I^2^=63%^55,121^); (case-control: 27.69[0.03;1000.00], I^2^=87% vs 24.59[13.45;44.95], I^2^=0.00)^133,134^; however, the pooled effect in cross-sectional studies and RCTs showed the opposite pattern (cross-sectional dose 1: 17.69[1.68;185.93], I^2^=45%, vs dose2: 8.23[0.96;70.59], I^2^=0.00)^38^; (trial dose1: 47.63[0.00;1000.00], I^2^=67% vs dose 2: 36.89[0.00;1000.00], I^2^=32%)^22,23^ (Figure 11). Subgroup analyses by population or vaccine type did not reveal any clear trends in proportions.

**Figure 11.**
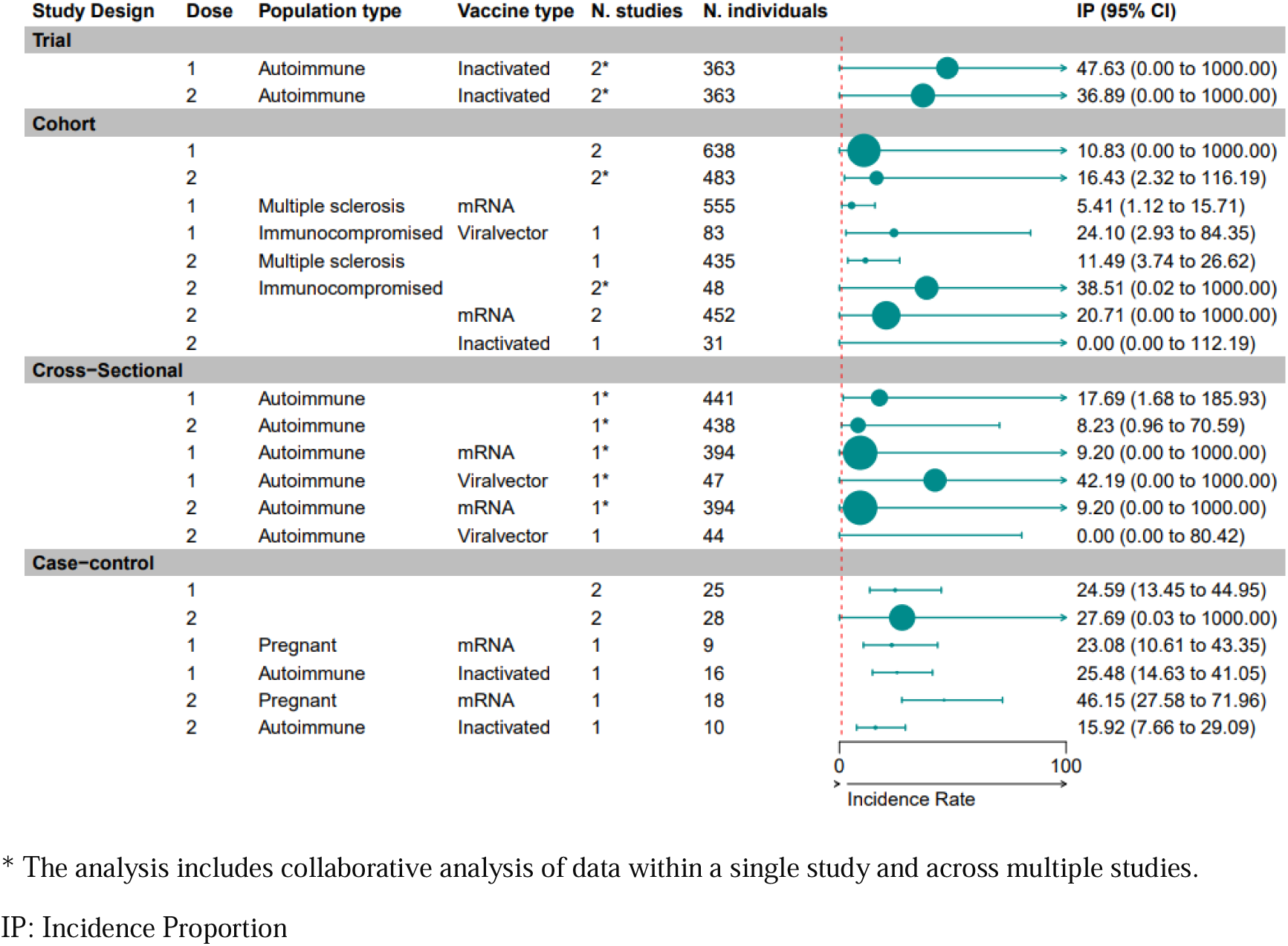
Neurological Symptoms Pooled Incidence Proportion.

### Neurological event

Analysis of cohort studies in cancer patients showed no significant difference between the pooled proportion of neurological events in doses 1 and 2 (dose1: 5.05[1.40;18.19], I^2^=0.00 vs dose2: 5.34[1.69;16.90], I^2^=0.00)^112^ (Figure 12).

**Figure 12.**
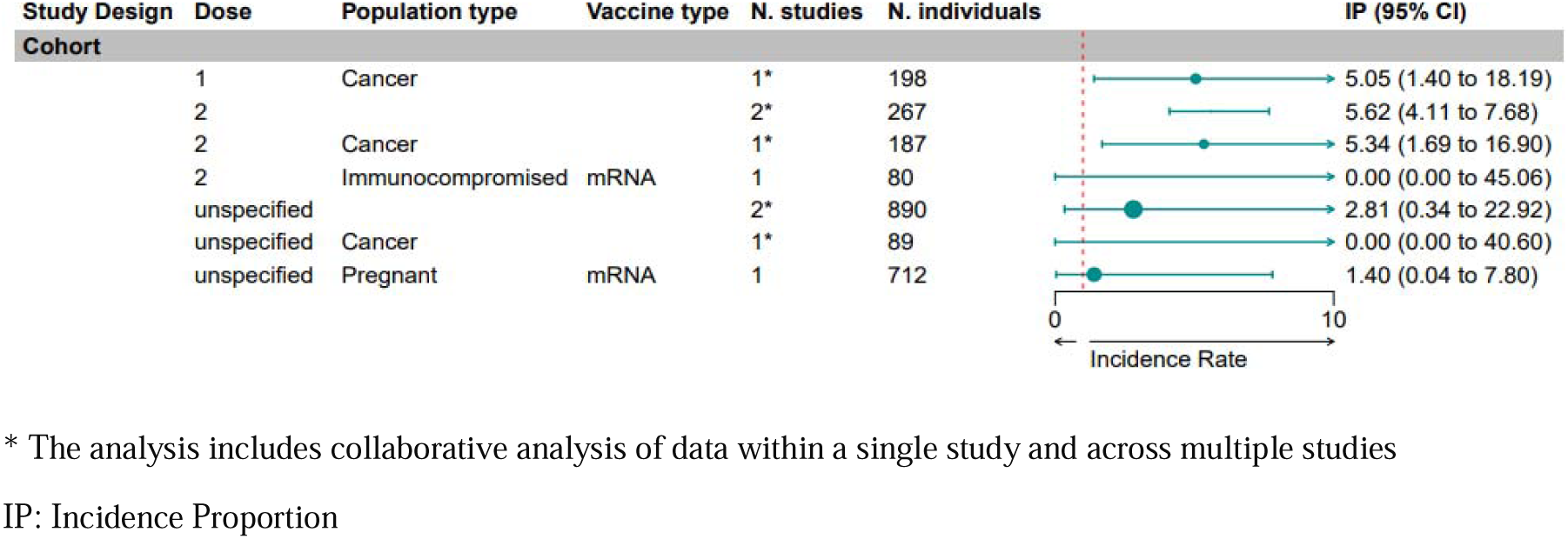
Neurological Events Pooled Incidence Proportion.

### Maternal and neonatal outcome

The most prevalent maternal outcomes following COVID-19 vaccination were gestational hypertension by the pooled proportion of 36.57[0.00;1000.00], I^2^=88%^115^ per 1000 observations followed by spontaneous or induced abortion (8.55[3.59;20.37], I^2^=96%)^126^ and stillbirth (3.65[2.64;5.05], I^2^=0.00)^115,123^. The most common neonatal outcomes were small for gestational age by a proportion of 73.18[9.67;553.68], I^2^=95%^106,120,123^, followed by neonatal hospitalization (33.89[4.10;280.14], I^2^=43%)^106,115,123^ (Figure 13-14-15).

**Figure 13.**
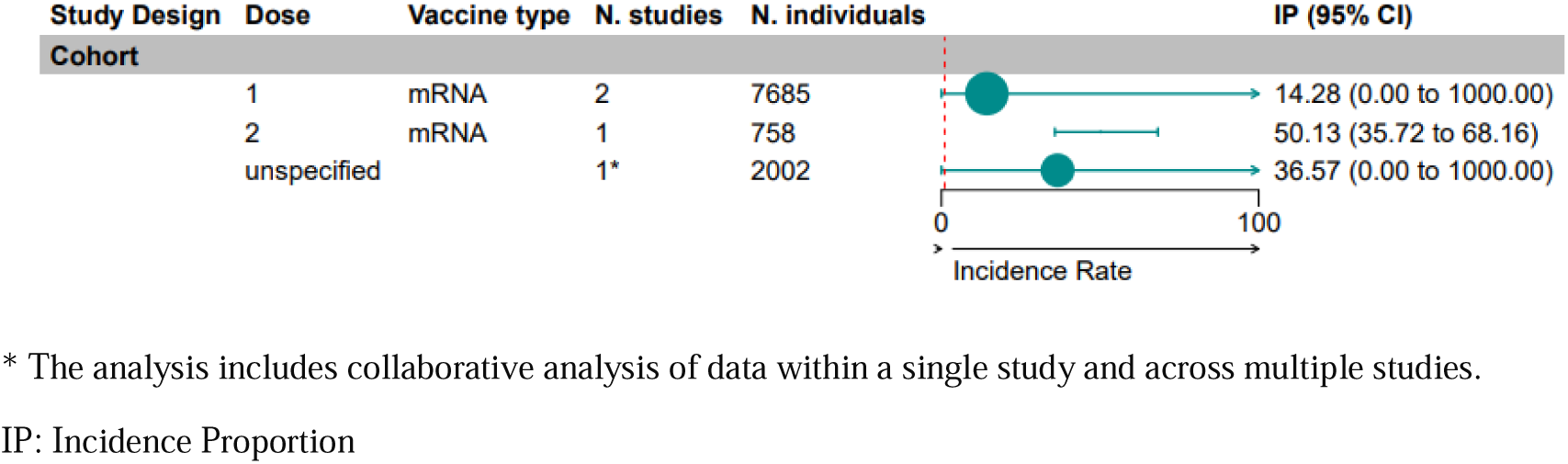
Gestational Hypertension Pooled Incidence Proportion.

**Figure 14.**
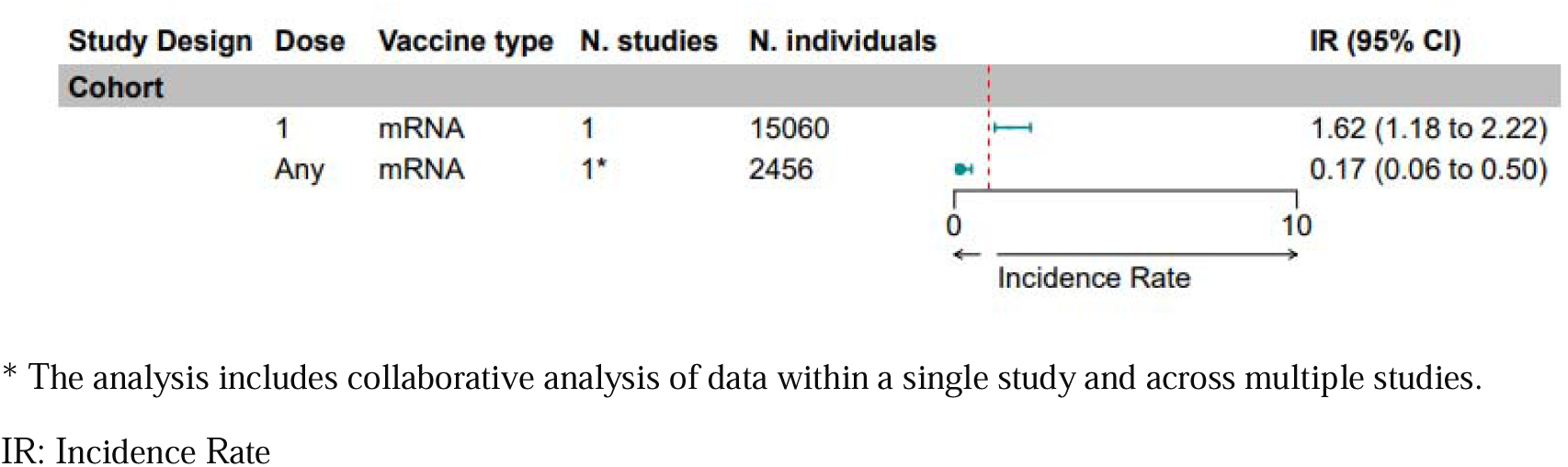
Spontaneous or Induced Abortion Pooled Incidence Rate.

**Figure 15.**
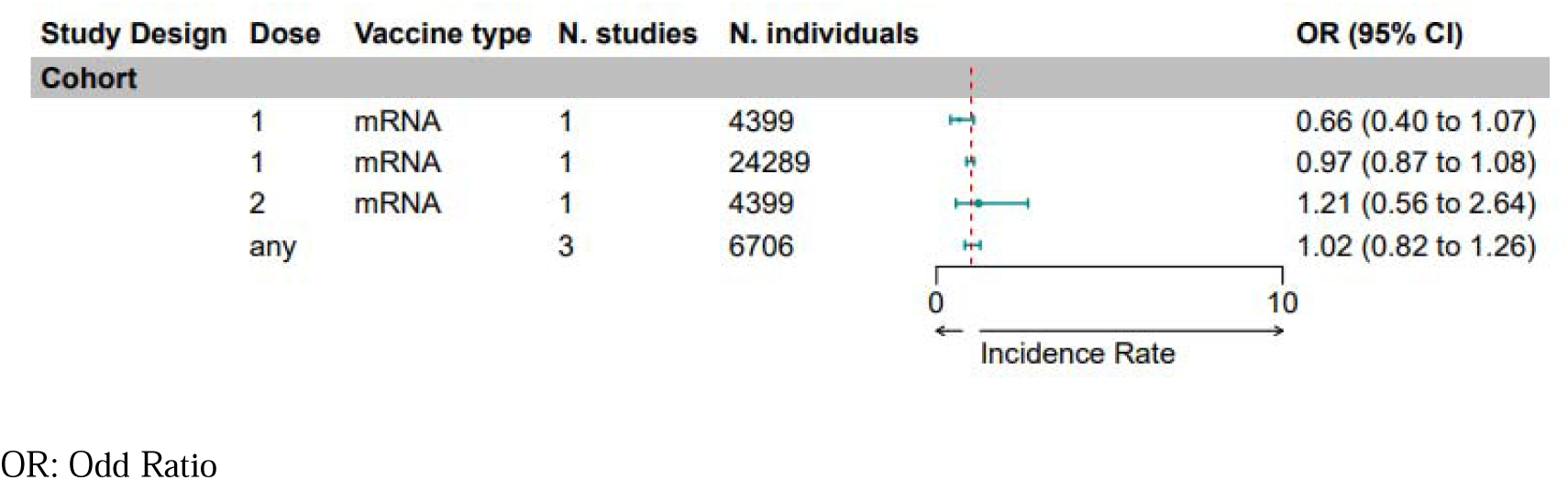
Small for Gestational Age Odd Ratio.

### Gestational hypertension

Cohort studies investigating maternal hypertensive disorders after mRNA vaccination indicate a higher pooled proportion of hypertensive disorders after the second dose (50.13[35.71;68.1]) compared to the first dose (14.28[0.00;1000.00], I^2^=99%^70,120^) (Figure 13).

### Abortion

In cohort studies of pregnant women vaccinated with mRNA vaccines, the pooled IR of spontaneous or induced abortion is 0.17[0.06;0.5], I^2^=95%^126^ (Figure 14).

### Small for gestational age (SGA)

The odd ratio (OR) of SGA infants born from mothers vaccinated with the first dose of COVID-19 mRNA vaccines in the two respective cohorts were 0.66[0.4;1.07]^120^ and 0.97[0.87;1.08]^71^, and the OR of SGA following dose 2 in one of these cohorts was 1.21[0.56;2.64]^120^ (Figure 15).

### Stillbirth

No stillbirth cases were reported in the two cohorts of 140 pregnant women who received either the mRNA or viral vector vaccine^115^. Among 7,530 cases in a separate cohort^70^ and 827 cases in a cross-sectional study^51^ receiving mRNA vaccine during pregnancy, only one stillbirth case was reported (Table S7).

### Thromboembolism

An RCT of 544 cancer patients vaccinated with mRNA vaccines presented that the proportion of thromboembolism events was 9.19[3.83-22.08] per 1000 observations^21^. A cohort study of 140 pregnant women showed no case of events in this population^115^. A cross-sectional study of over 2500 inflammatory bowel syndrome patients showed that the proportion of thromboembolic events was higher following dose two compared to dose one (1.16[0.37;3.58] vs 0.77[0.19;3.08])^35^ (Table S7).

### Deaths

Two cohort studies involving pregnant women who received mRNA/viral vector vaccines found no reported deaths^70,115^. In another cohort of 435 MS patients who received the second dose of the mRNA vaccine, one death was reported^55^. Additionally, in a trial of 1,902 cancer patients who received the mRNA vaccine, one death was reported^24^. In a study of over 16,600 newborns born from mothers who received the COVID-19 vaccine, the OR of neonatal deaths was 0.84[0.43;1.72], suggesting no significant differences between neonatal death in vaccinated and unvaccinated mothers^71^ (Supplementary Table 7).

### Non-sever outcomes

Within the scope of our analysis, fatigue, myalgia, arthralgia, and gastrointestinal symptoms were the most prevalent symptoms. Figures 16 to 22 encapsulate the data about non-severe systemic outcomes and their sub-grouped analysis (Table S8).

**Figure 16.**
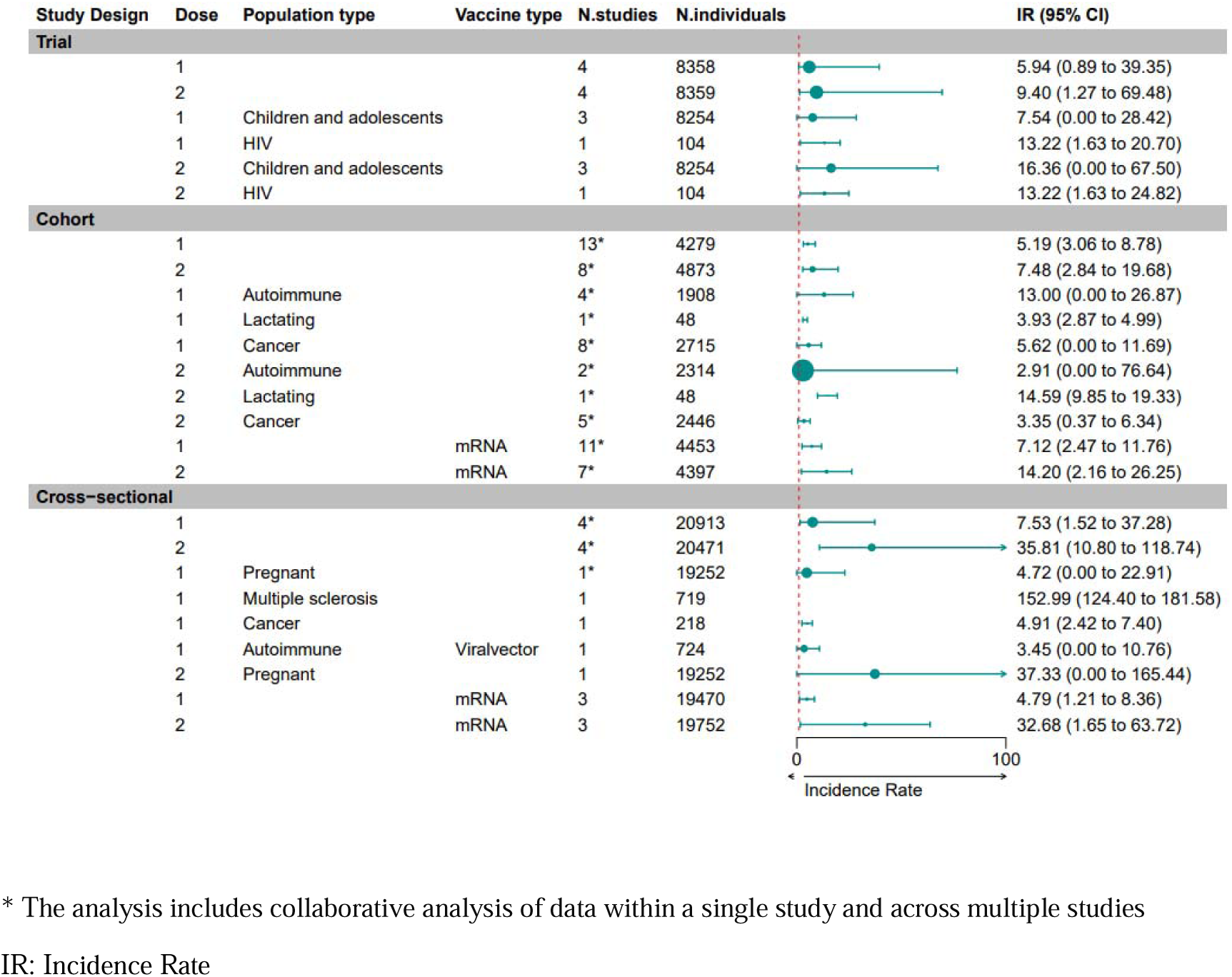
Arthralgia Pooled Incidence Rate.

**Figure 17.**
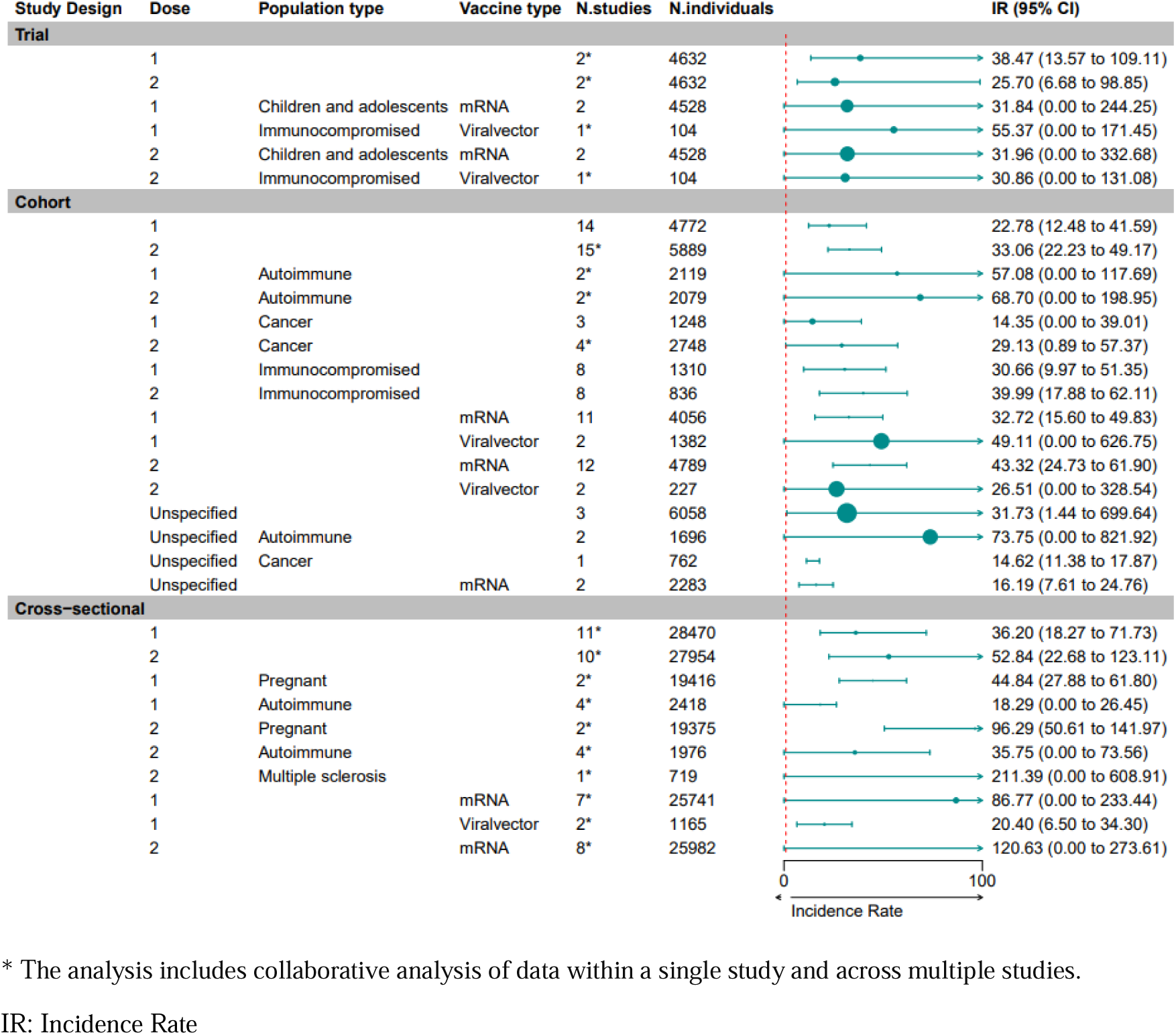
Fatigue Pooled Incidence Rate.

**Figure 18.**
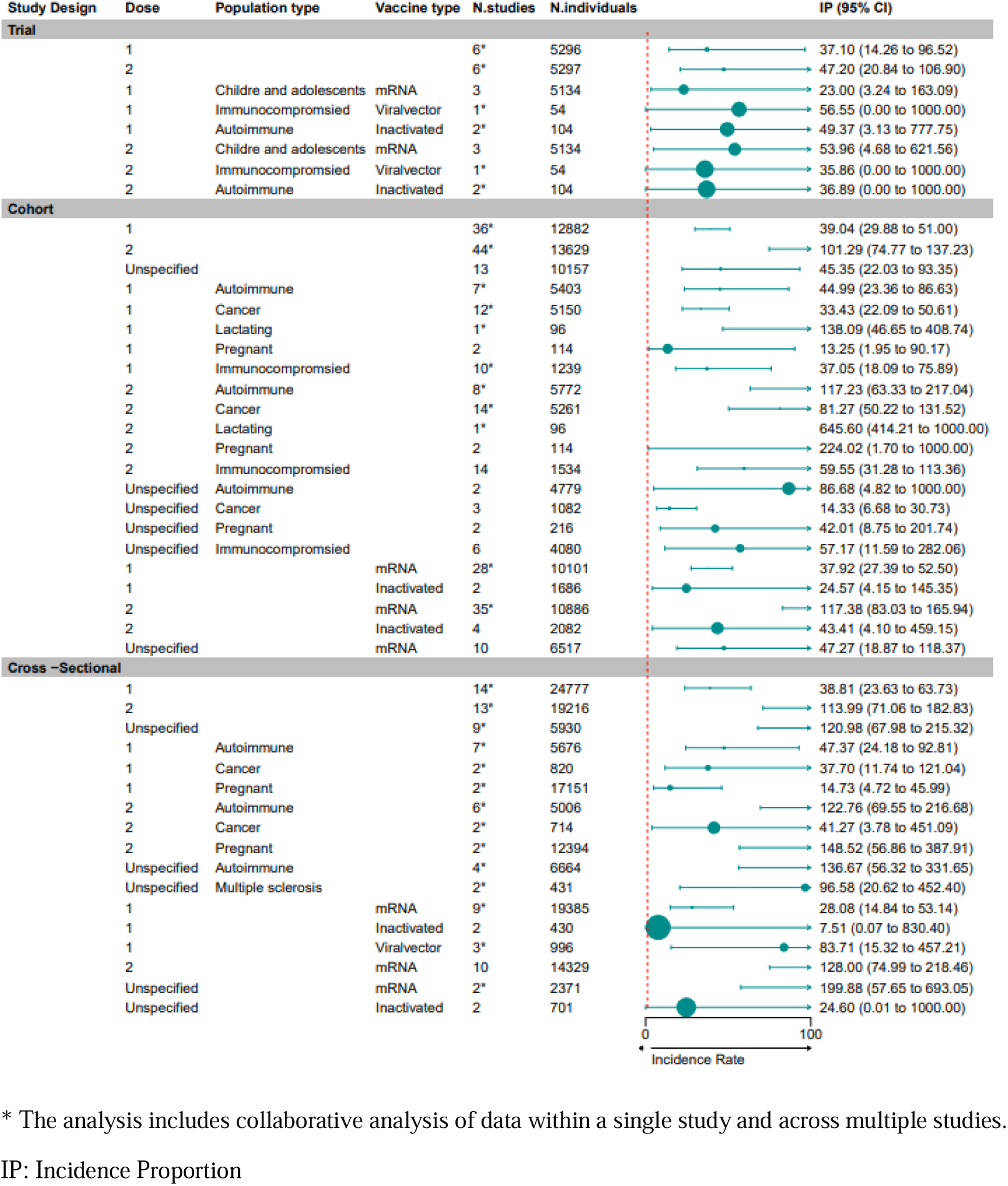
Fever Pooled Incidence Proportion.

**Figure 19.**
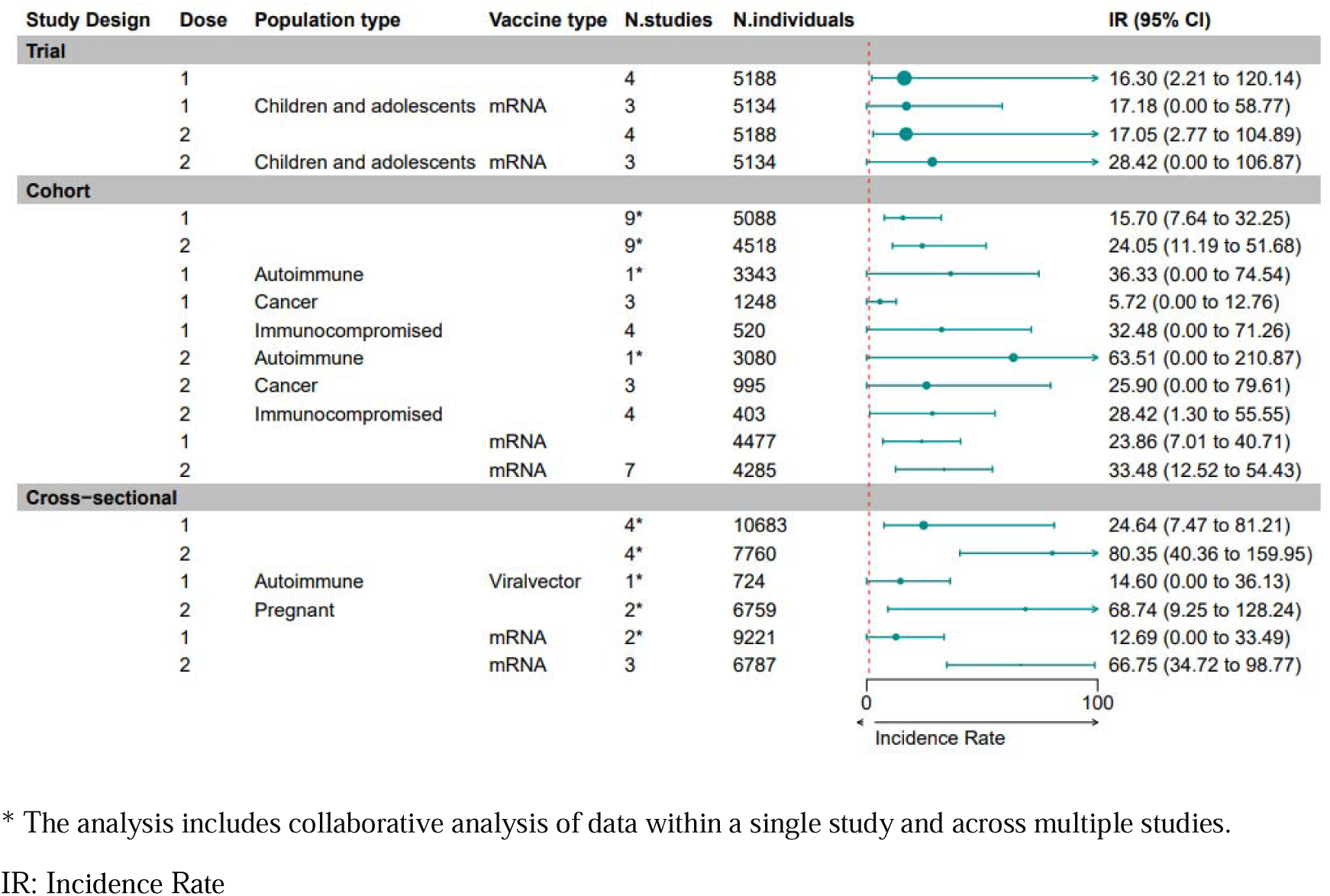
Myalgia Pooled Incidence Rate.

**Figure 20.**
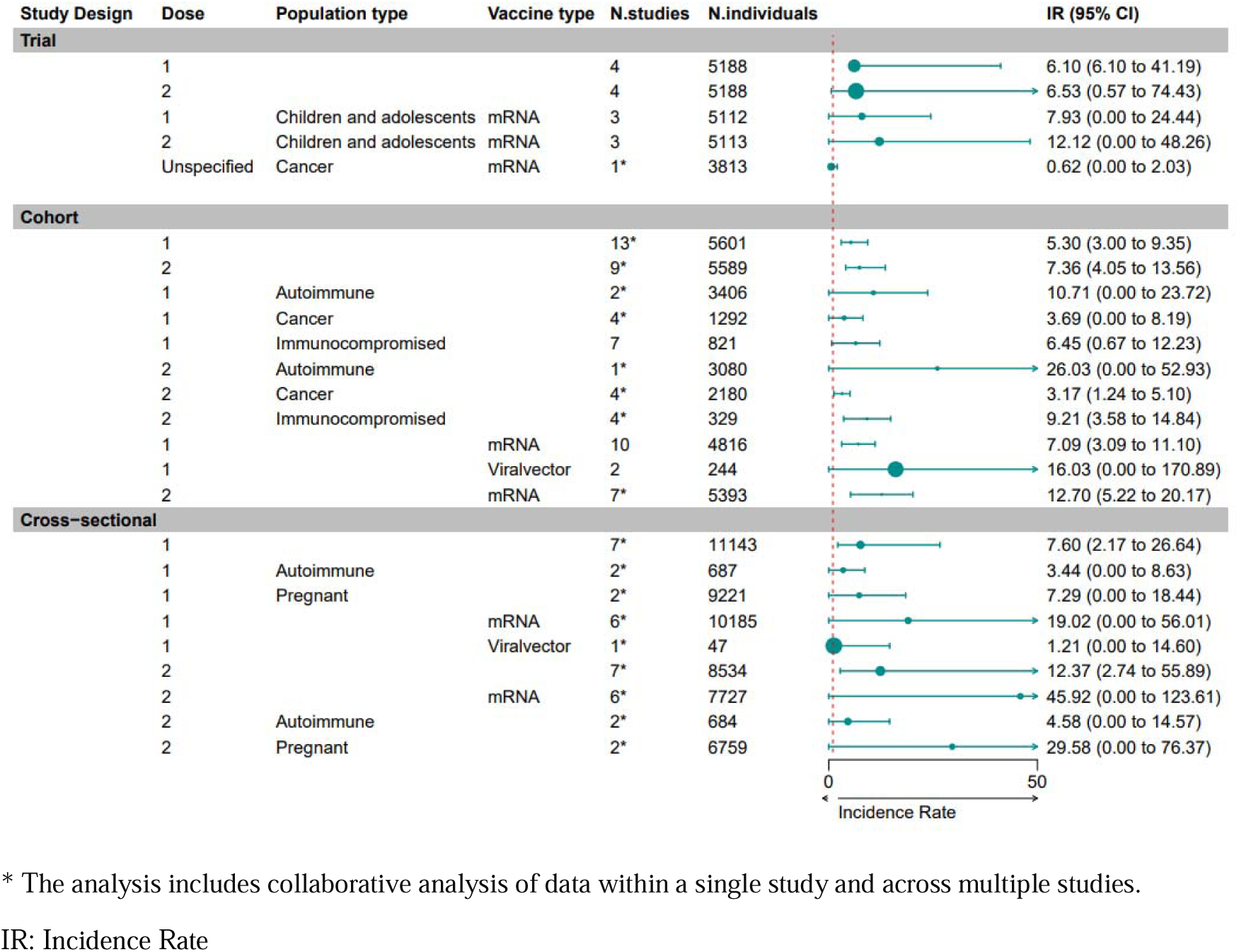
Gastrointestinal Symptoms Pooled Incidence Rate.

**Figure 21.**
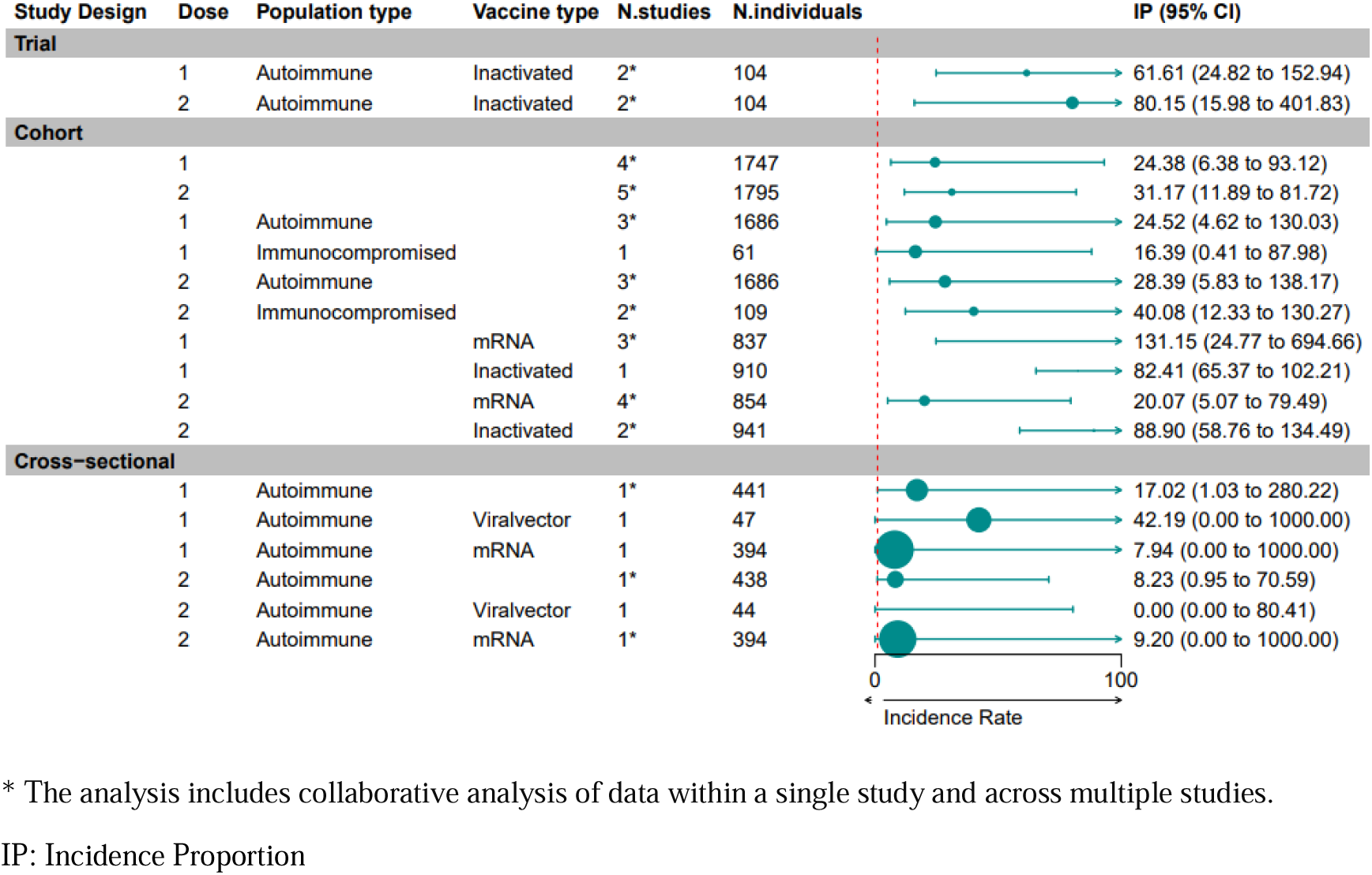
Ear, Nose, Throat symptoms Pooled Incidence Proportion.

**Figure 22.**
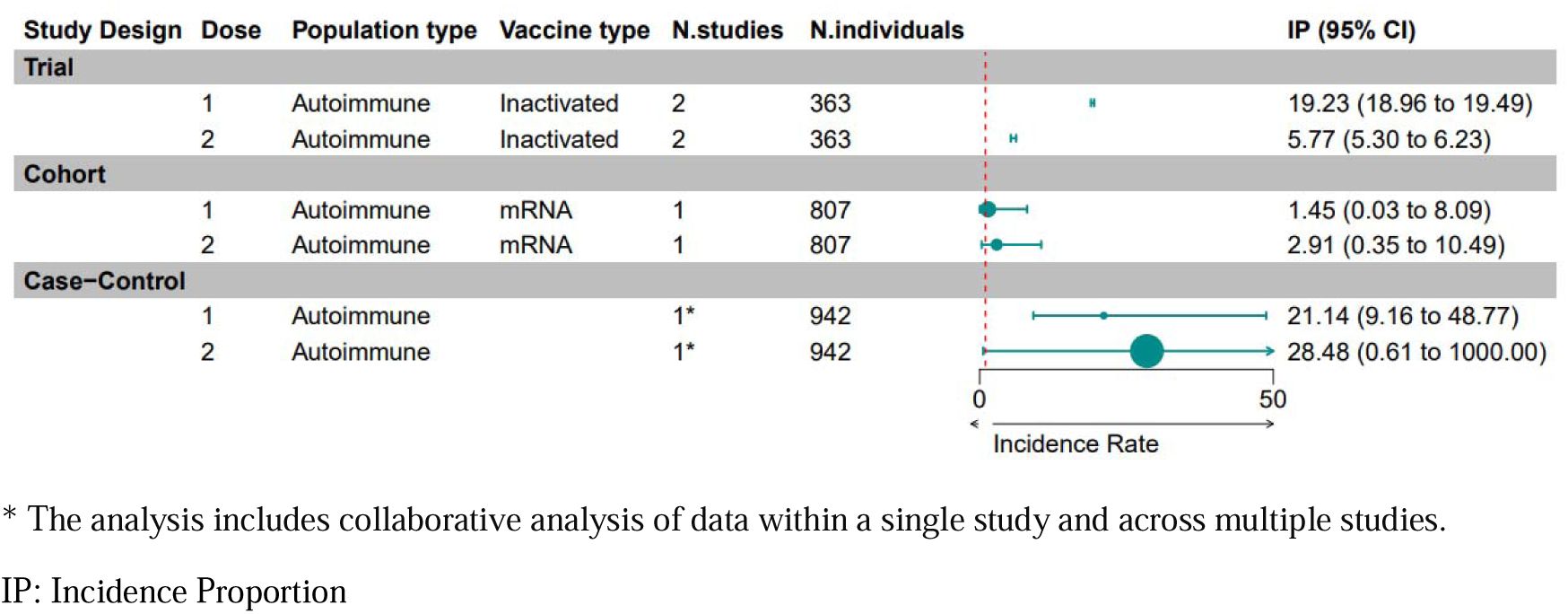
Conjunctivitis and Uveitis Pooled Incidence Proportion.

### Publication Bias

The funnel plots represent each reported effect estimate as an individual point. Each study includes several rates for different outcomes and vaccines, consequently, each study contributes multiple points to the plot. The funnel plots exhibit some skewness, suggesting potential publication bias within this literature, however, due to the generally low standard errors, the likelihood of publication bias attributed to sample size appears to be minimal (Figure S2-3).

## Discussion

The rapid development and widespread deployment of COVID-19 vaccines have been pivotal in mitigating the impact of the global pandemic. However, given the limited representation of vulnerable groups in pivotal clinical trials and RCTs, further investigation into vaccine safety within these populations is imperative to inform vaccination strategies and enhance safety monitoring efforts. This study pooled the IRs and proportions of COVID-19 vaccine-induced outcomes across diverse vaccine platforms, study designs, and populations. Our finding showed that among the reported outcomes, the top four most prevalent severe events were lymphadenopathy, autoimmune disease and MS flare-up, and cardiac symptoms. Allergic reactions were more common among patients with autoimmune diseases, followed by cancer patients, who most often experienced lymphadenopathy. Autoimmune disease patients had more flare-ups than those with MS. Lymphadenopathy, cardiac events, cardiac symptoms, and hospitalizations were more frequent after the second vaccine dose. Cardiac events and hypertensive disorders were more common with mRNA vaccines, while allergic reactions were more frequent with viral vector vaccines. The adverse effects of mRNA vaccines are higher due to their earlier approval and broader distribution^16^. No link was found between COVID-19 vaccination and maternal or neonatal outcomes. Cardiac events were more common with mRNA vaccines in cohort studies, especially after the second dose. However, cross-sectional studies had conflicting results due to smaller sample sizes and vaccine brand variability.

The literature reviews on the cancer population revealed a significant number of cases of lymphadenopathy following COVID-19 vaccination, advising clinicians to rely on the patient’s clinical context and current resources to avoid probable disease progression and therapy changes^136^ ^137^. Negahdaripour et al.’s study supports overall vaccine safety for cancer patients ^138^, supporting our finding that serious adverse events are rare. Our results aligned with this study, showing nuanced susceptibility in immunocompromised individuals. This susceptibility might depend on the medication type administered to the patients, which impacts the activation of immune cells and their seropositivity^138^. Studies recommend avoiding vaccination during immunosuppressive treatment or waiting 4-6 months post-medication to minimize complications and ensure safety^138^.

Our findings revealed a high IR of flare-ups in autoimmune diseases, particularly after the second vaccine dose. This is different from Patrick’s study, where autoimmune-immunocompromised individuals did not experience an exacerbation, possibly due to ongoing immunosuppressive treatment^139^. Shabani et al.’s systematic review shows more flare-ups in immune-mediated diseases compared to MS, and higher flare-ups with mRNA vaccines than with viral vector and inactivated vaccines^140^, aligning with our research findings. It suggests avoiding vaccination during disease flare-ups and tapering steroid therapy^141^. Yang’s study demonstrated disease flares after inactivated COVID-19 vaccination^141^. Our study compared flare-ups across different vaccine types, finding higher rates with mRNA vaccines than inactivated or viral vector vaccines, likely due to most studies focusing on mRNA vaccines and fewer on other vaccine types, though further investigation is needed^142^. In our study, cardiac events were rare in immunocompromised, autoimmune, and cancer patients, aligning with a systematic review that found myocarditis and pericarditis rates post-vaccination similar to the general population ^143^. Thus, emphasizing the significance of the overall advantages of COVID-19 vaccination.

Our analysis showed that severe events in vaccinated children and adolescents are rare, consistent with systematic reviews confirming the rarity of serious adverse events^139,141^. Hause et al. noted a few serious adverse events in children under five, including anaphylaxis and afebrile seizures, with the majority being non-serious events like fever, rash, vomiting, urticaria, and fatigue^144^.

Annamaria et al.’s review identifies common pregnancy-related AESIs ^141,145^. Our findings aligned, showing SGA as the most prevalent fetal adverse event and pregnancy loss as the most common maternal outcome. Unlike the Annamaria study, we found a neonatal death rate of 1.44 per 1000 person days, but no significant link to vaccination (OR: 0.8). Both Kang’s and Li Yang’s studies found no significant difference in adverse event rates post-COVID-19 vaccination between HIV patients and healthy controls^146,147^. Although our study, which included HIV patients in the immunocompromised group, found a high rate of non-severe adverse events, these rates may not differ from those in healthy populations, indicating the need for further investigation.

To our knowledge, this study is the first comprehensive systematic review that addresses a crucial gap in understanding the safety profiles of COVID-19 vaccines, particularly within vulnerable populations. Our study encompassed a diverse range of vulnerable populations, including immunocompromised individuals, cancer patients, pregnant and breastfeeding women, children, adolescents, individuals with disabilities, and migrants. We analysed observational studies and clinical trial data to elucidate these groups’ IRs of adverse events following COVID-19 vaccination. Broad inclusion criteria and detailed subgroup analyses of vaccine types and doses provided valuable insights for vaccination strategies and safety monitoring.

Our study has several limitations that should be acknowledged. We reduced heterogeneity in the overall analysis through subgroup analyses. However, the optimal approach would have been to group individuals with varying immune characteristics, such as those with HIV or drug-induced immunosuppression into a single category. However, separate analyses for these subgroups were not feasible due to the limited number of available studies. This limitation highlights the need for more granular data in future research to allow for more precise subgroup analyses. Limited gender information restricted sex-based analyses, and certain populations, such as refugees and prisoners, were not presented in any study. Furthermore, some studies utilize passive surveillance methods, such as self-reported data, which may not be as reliable as data reported by the healthcare system following direct observation. In addition the predominance of studies from high-income countries limits generalizability to low- and middle-income settings. Finally, small sample sizes, wide confidence intervals, and varying study quality highlight the need for larger and more representative datasets in future research.

## Conclusion

In conclusion, our review provides valuable insights into the safety profiles of COVID-19 vaccines within vulnerable populations. By systematically analyzing various adverse events across diverse populations and vaccine platforms, our findings contribute to the ongoing efforts to optimize vaccination strategies and enhance safety monitoring efforts in the fight against the COVID-19 pandemic. Further research using real-world data is warranted to validate and expand upon our findings, focusing on addressing gaps in vaccine safety knowledge within vulnerable populations.

## Data Availability

All data produced in the present work are contained in the manuscript

