## Supplemental tables for "COVID-19 Vaccine Safety Studies among Vulnerable Populations: A Systematic Review and Meta-analysis of 120 Observational Studies and Randomized Clinical Trials": Supplementary tables.pdf

**Table S1.Search Terms**

| **Population/Domain** | **Exposure** | **Outcomes** |
|---|---|---|
| Immunocompromised | COVID-19 vaccine* | Side effect* |
| Immunosupress* | SARS-COV-2 vaccine* | Advers* |
| HIV | BNT162* | Adverse event* |
| AIDS | Comirnaty | Adverse effect* |
| Solid tumour* | Tozinameran | Adverse events of special interest (AESI) |
| Hematologic malignanc* | mRNA-1273 | Adverse event following Immunization (AEFI) |
| Solid organ transplant* | Spikevax | Reactogenicity |
| Hematopoietic cell transplant* | ZyCov-D | Acute respiratory distress syndrome |
| Pregnan* | Ad5-nCOV | Multisystem inflammatory syndrome |
| Breastfeed* | Ad26.COV2.S | Acute cardiovascular injury |
| Postpartum | ChAdOx1* | Myocarditis |
| Delivery | Vaxzevria | Pericarditis |
| Post-abortion | Covishield | Microangiopathy |
| Post-miscarriage | BBIBP-CorV | Heart Failure |
| Child* | GAM-Covid-VAC | Stress cardiomyopathy |
| Neonat* | Sputnik V | Coronary artery disease |
| Infan* | Covaxin | Arrhythmia |
| Adolescent* | QazCovid-in | Coagulation disorders |
| Refugee | KCONVAC | thrombotic disorders |
| Migrant* | COVIran | bleeding disorders |
| Undocumented migrant* | CoronaVac | Anosmia |
| IDP* | WIBP | Ageusia |
| Internally Displaced People | ZF2001 | Chilblain – like lesions |
| Asylum seeker* | CIGB-66/Abdala | Erythema multiforme |
| Disabled | Soberana 2 | Single Organ Cutaneous Vasculitis |
| Disabilit* | MVC-COV1901 | Acute kidney injury |
| Transgender | NVX-CoV2373 | Acute liver injury |
| Trans-sexual | Covovax | Acute pancreatitis |
| Prisoner* | SpikoGen | Rhabdomyolysis |
| | EpiVacCorona | Subacute thyroiditis |
| | RNA vaccine* | Anaphylaxis |
| | Adenovirus vaccine* | Thrombocytopenia |
| | Inactivated vaccine* | Generalized convulsion. |
| | Subunit vaccine* | Acute disseminated encephalomyelitis |
| | Other international nonproprietary name (e.g., tozinameran) | Guillain Barré Syndrome |
| | | Acute aseptic arthritis |
| | | Aseptic meningitis |
| | | Encephalitis |
| | | Encephalomyelitis |
| | | Idiopathic Peripheral Facial Nerve Palsy |
| | | Vaccine associated enhanced disease |
| | | Joint Pain |
| | | Fatigue |
| | | Headache |
| | | Muscle pain |
| | | Fever |
| | | Chills |
| | | Asthenia |
| | | Tolera* |
| | | Safety assessment |
| | | Safety analysis |
| | | Safety criteria* |
| | | Safety profile* |

### Table S2.Searching String

| Ovid MEDLINE(R) ALL <1946 to February 18, 2022> | Query | Results | Type |
|---|---|---|---|
| 1 | ((2019 nCoV or 2019nCoV or corona virus or corona viruses or coronavirus or coronaviruses or COVID or COVID19 or nCov 2019 or SARS-CoV2 or SARS CoV-2 or SARSCoV2 or SARSCoV-2).ti,ab,kf. or (exp "COVID-19"/ or exp "COVID-19 Testing"/ or exp "COVID-19 Vaccines"/ or "Coronavirus"/ or exp "Receptors, Coronavirus"/ or exp "SARS-CoV-2"/ or exp "Spike Glycoprotein, Coronavirus"/)) not (exp "animals"/ not "humans"/) not (editorial or newspaper article).pt. | 226749 | Advanced |
| 2 | (vaccine or abdavomeran or "ad26.cov2.s" or ag0302-covid19 or bbibp-corv or bnt-162 or bnt162b2 or convidicea or coronavac covaxin or covlp or elasomeran or epivaccorona or ganulameran or nvx-cov2373 or pidacmeran or Pittsburgh-coronavirus-vaccine or reluscovtogene-ralaplasmid or (SCB-2019 adj vaccine) or sputnik or tozinameran or vaxzevria or vidprevtyn or wibp-corv or zifivax or zorecimeran or mRNA-1273 or JNJ-78436735 or "Ad26.COV2.S" or AZD1222 or ChAdOx1 or CoronaVac or GAM-Covid-VAC or NVX-CoV2373).ti,ab,kf. | 224152 | Advanced |
| 3 | exp COVID-19 Vaccines/ | 9094 | Advanced |
| 4 | 2 or 3 | 227180 | Advanced |
| 5 | 1 and 4 | 19399 | Advanced |
| 6 | (safe or safety or side-effect* or undesirable effect* or treatment emergent or tolerability or toxicity or adrs or (adverse adj2 (effect or effects or reaction or reactions or event or events or outcome or outcomes))) .ti,ab. | 1818572 | Advanced |
| 7 | (ae or to or co).fs. | 4174141 | Advanced |
| 8 | 6 or 7 | 5362116 | Advanced |
| 9 | (allergic or anaphylaxi* or thrombo* or (blood adj2 cloth*) or heart or coronary or arrhythmia or myocard* or pericard* or menstrual or period or bleeding or coagulation or anosmia or ageusia or chilblain or erythema or vasculitis or (acute adj2 (kidney or liver or pancreat* or "aseptic arthritis")) or rhabdomyolysis or thyroiditis or convulsion or encephal* or Comirnat*).ti,ab,kf. | 3685495 | Advanced |
| 10 | (cohort or (control and study) or (control and group*)).tw. or exp epidemiologic studies/ or program.tw. or clinical trial.pt. or comparative-stud*.tw. or evaluation-studies.tw. or exp statistics as topic/ or survey*. tw. or follow-up*.tw. or time factors.tw. or ci.tw. | 7848938 | Advanced |
| 11 | case reports.pt. or (case adj (stud* or repor*)).ti,ab. or case.ti. | 2650495 | Advanced |
| 12 | (MEDLINE or systematic-review or (literature adj2 review)).tw. or (search* adj12 (literature or database?)).ti,ab. | 512630 | Advanced |
| 13 | ((randomized controlled trial or controlled clinical trial).pt. or drug therapy.fs. or (randomized or randomized or placebo or randomly or trial or groups).ab.) [illegible] | 4600894 | Advanced |
| 14 | 10 or 12 or 13 | 10394274 | Advanced |
| 15 | (animals/ not humans/) or comment.pt. or editorial.pt. or exp consensus/ | 6283005 | Advanced |
| 16 | 11 or 15 | 8860181 | Advanced |
| 17 | 8 or 9 | 7937672 | Advanced |
| 18 | 5 and 14 and 17 | 3470 | Advanced |
| 19 | 18 not 16 | 3128 | |

| Embase | Query | Results | Date |
|---|---|---|---|
| #13 | #12 AND [embase]/lim | 1635 | 21-Feb-22 |
| #12 | #7 AND #11 | 1744 | 21-Feb-22 |
| #11 | #8 OR #9 OR #10 | 13728750 | 21-Feb-22 |
| #10 | methodology'/exp OR cohort:ti,ab,kw,de OR ((control NEAR/5 study):ti,ab,kw,de) OR ((control NEAR/3 group*):ti,ab,kw,de) OR program:ti,ab OR 'comparative stud*':ti,ab,kw,de OR 'evaluation studies':ti,ab,kw,de OR survey*:ti,ab,kw,de OR 'follow up*':ti,ab,kw,de OR 'time factors':ti,ab,kw,de OR ci:ti,ab,kw,de | 11528926 | 21-Feb-22 |

| | | | |
|---|---|---|---|
| #9 | (medline:ti,ab,kw,de OR systematic) AND 'review'/exp OR 'systematic review':ti,ab,kw,de OR 'meta analysis'/exp OR ((search* NEAR/12 (literature OR database?)):ti,ab) | 646635 | 21-Feb-22 |
| #8 | ('randomized controlled trial'/de OR 'controlled clinical trial'/de OR random*:ti,ab,tt OR 'randomization'/de OR 'intermethod comparison'/de OR placebo:ti,ab,tt OR compare:ti,tt OR compared:ti,tt OR comparison:ti,tt OR ((evaluated:ab OR evaluate:ab OR evaluating:ab OR assessed:ab OR assess:ab) AND (compare:ab OR compared:ab OR comparing:ab OR comparison:ab)) OR ((open NEXT/1 label):ti,ab,tt) OR (((double OR single OR doubly OR singly) NEXT/1 (blind OR blinded OR blindly)):ti,ab,tt) OR 'double blind procedure'/de OR ((parallel NEXT/1 group*):ti,ab,tt) OR crossover:ti,ab,tt OR 'cross over':ti,ab,tt OR (((assign* OR match OR matched OR allocation) NEAR/6 (alternate OR group OR groups OR intervention OR interventions OR patient OR patients OR subject OR subjects OR participant OR participants)):ti,ab,tt) OR assigned:ti,ab,tt OR allocated:ti,ab,tt OR ((controlled NEAR/8 (study OR design OR trial)):ti,ab,tt) OR volunteer:ti,ab,tt OR volunteers:ti,ab,tt OR 'human experiment'/de OR trial:ti,tt) NOT (((random* NEXT/1 sampl* NEAR/8 ('cross section*' OR questionnaire* OR survey OR surveys OR database OR databases)):ti,ab,tt) NOT ('comparative study'/de OR 'controlled study'/de OR 'randomised controlled':ti,ab,tt OR 'randomized controlled':ti,ab,tt OR 'randomly assigned':ti,ab,tt) OR ('cross-sectional study' NOT ('randomized controlled trial'/de OR 'controlled clinical study'/de OR 'controlled study'/de OR 'randomised controlled':ti,ab,tt OR 'randomized controlled':ti,ab,tt OR 'control group':ti,ab,tt OR 'control groups':ti,ab,tt)) OR ('case control*':ti,ab,tt AND random*:ti,ab,tt NOT ('randomised controlled':ti,ab,tt OR 'randomized controlled':ti,ab,tt)) OR ('systematic review':ti,tt NOT (trial:ti,tt OR study:ti,tt)) OR (nonrandom*:ti,ab,tt NOT random*:ti,ab,tt) OR 'random field*':ti,ab,tt OR (('random cluster' NEAR/4 sampl*):ti,ab,tt) OR (review:ab AND review:it NOT trial:ti,tt) OR ('we searched':ab AND (review:ti,tt OR review:it)) OR 'update review':ab OR ((databases NEAR/5 searched):ab) OR ((rat:ti,tt OR rats:ti,tt OR mouse:ti,tt OR mice:ti,tt OR swine:ti,tt OR porcine:ti,tt OR murine:ti,tt OR sheep:ti,tt OR lambs:ti,tt OR pigs:ti,tt OR piglets:ti,tt OR rabbit:ti,tt OR rabbits:ti,tt OR cat:ti,tt OR cats:ti,tt OR dog:ti,tt OR dogs:ti,tt OR cattle:ti,tt OR bovine:ti,tt OR monkey:ti,tt OR monkeys:ti,tt OR trout:ti,tt OR marmoset*:ti,tt) AND 'animal experiment'/de) OR ('animal experiment'/de NOT ('human experiment'/de OR 'human'/de))) | 5066761 | 21-Feb-22 |
| #7 | #3 AND #6 | 2929 | 21-Feb-22 |
| #6 | #4 OR #5 | 7193524 | 21-Feb-22 |
| #5 | allergic:ti,ab,kw OR anaphylaxi*:ti,ab,kw OR thrombo*:ti,ab,kw OR ((blood NEAR/2 cloth*):ti,ab,kw) OR heart:ti,ab,kw OR coronary:ti,ab,kw OR arrhythmia:ti,ab,kw OR myocard*:ti,ab,kw OR pericard*:ti,ab,kw OR menstrual:ti,ab,kw OR period:ti,ab,kw OR bleeding:ti,ab,kw OR coagulation:ti,ab,kw OR anosmia:ti,ab,kw OR ageusia:ti,ab,kw OR chilblain:ti,ab,kw OR erythema:ti,ab,kw OR vasculitis:ti,ab,kw OR ((acute NEAR/2 (kidney OR liver OR pancreat* OR 'aseptic arthritis')):ti,ab,kw) OR rhabdomyolysis:ti,ab,kw OR thyroiditis:ti,ab,kw OR convulsion:ti,ab,kw OR encephal*:ti,ab,kw OR comirnat*:ti,ab,kw | 5193220 | 21-Feb-22 |
| #4 | safe:ti,ab OR safety:ti,ab OR 'side effect*':ti,ab OR 'undesirable effect*':ti,ab OR 'treatment emergent':ti,ab OR tolerability:ti,ab OR toxicity:ti,ab OR adrs:ti,ab OR ((adverse NEAR/2 (effect OR effects OR reaction OR reactions OR event OR events OR outcome OR outcomes)):ti,ab) | 2666918 | 21-Feb-22 |
| #3 | #1 AND #2 | 7333 | 21-Feb-22 |
| #2 | anti-sars-cov-2 agent'/exp OR vaccine:ti,ab,kw OR abdavomeran:ti,ab,kw OR 'ag0302 covid19':ti,ab,kw OR 'bbibp corv':ti,ab,kw OR 'bnt 162':ti,ab,kw OR bnt162b2:ti,ab,kw OR convidicea:ti,ab,kw OR ((coronavac NEAR/1 covaxin):ti,ab,kw) OR covlp:ti,ab,kw OR elasomeran:ti,ab,kw OR epivaccorona:ti,ab,kw OR ganulameran:ti,ab,kw OR pidacmeran:ti,ab,kw OR 'pittsburgh coronavirus vaccine':ti,ab,kw OR 'reluscovtogene ralaplasmid':ti,ab,kw OR (('scb 2019' NEAR/1 vaccine):ti,ab,kw) OR sputnik:ti,ab,kw OR tozinameran:ti,ab,kw OR vaxzevria:ti,ab,kw OR vidprevtyn:ti,ab,kw OR 'wibp corv':ti,ab,kw OR zifivax:ti,ab,kw OR zorecimeran:ti,ab,kw OR 'mrna 1273':ti,ab,kw OR 'jnj 78436735':ti,ab,kw OR 'ad26.cov2.s':ti,ab,kw OR azd1222:ti,ab,kw OR chadox1:ti,ab,kw OR coronavac:ti,ab,kw OR 'gam covid vac':ti,ab,kw OR 'nvx cov2373':ti,ab,kw | 278137 | 21-Feb-22 |

| | | | |
|---|---|---|---|
| #1 | (('coronaviridae'/exp AND 'coronavirinae'/exp OR 'coronaviridae infection'/exp OR 'coronavirus disease 2019'/exp) AND r AND 'coronavirus infection'/exp OR 'covid-19 testing'/exp OR 'sars coronavirus 2 test kit'/exp OR 'sars-related coronavirus'/exp OR 'severe acute respiratory syndrome coronavirus 2'/exp OR '2019 ncov':ti,ab,kw OR 2019ncov:ti,ab,kw OR (((corona* OR corono*) NEAR/1 (virus* OR viral* OR virinae*)):ti,ab,kw) OR coronavir*:ti,ab,kw OR coronovir*:ti,ab,kw OR covid:ti,ab,kw OR covid19:ti,ab,kw OR hcov*:ti,ab,kw OR 'ncov 2019':ti,ab,kw OR 'sars cov2':ti,ab,kw OR 'sars cov 2':ti,ab,kw OR sarscov2:ti,ab,kw OR 'sarscov 2':ti,ab,kw) NOT (('animal experiment'/exp OR 'animal experiment' OR 'animal'/exp OR 'animal') NOT ('human'/exp OR 'human' OR 'human experiment'/exp OR 'human experiment')) NOT 'editorial'/it NOT ([medline]/lim OR [pubmed-not-medline]/lim) AND [1-12-2019]/sd | 73751 | 21-Feb-22 |

### Table S3.Measured Outcomes

**Classification of outcomes**

Accelerated allergic reactions: anaphylaxis and any allaergic reactions occured less than 48 hours

Arthralgia

Cardiac events: myocarditis,pericarditis, arrhythmia

Cardiac symptoms:Chest pain,palpitation

Conjunctivitis and Uveitis.

Death

Disease exacerbation, flare or relaps

Ear-nose-throat symptoms: coryza, sneezing, throat ache, Rhinorrhea, stuffy nose

Fatigue : fatigue,asthenia,weakness,tiredness, malaise

Fever

Gastrointestinal symptoms :Nausea, vomiting, abdominal pain, diarrhea

Gestational hypertension

Hospitalization;

Hypertensive disorder

Induced or spontaneous abortion

Lymphadenopathy : local lymphadenopthy, axilary lymphadenopathy , unspecified lymphadenopathy, hypermetabolic axillary lymph nodes , lymphadenitis, swollen gland, lymph node swelling

Muscle ache and Myalgia

Neurological event : radiculitis , sensory neuropathy, seizure, convulsion

Neurological symptoms : Numbness , paresthesia, Face tingling, tinnitus, tremor

respiratory symptoms : shortness of breath,cough,wheezing,dyspnea ,mechanical ventilation

small for gestational age ; Intrautherin growth restriction

stillbirth

Systemic allergic reactions: allergic reaction unspecified,angioedema,severe allergic reaction, systemic swelling, head and tongue swelling

Thromboembolims

### Table S4.Risk window period

| Measured outcome | Risk window |
|---|---|
| Lymphadenopathy risk window 0-14days [7] | 0-14 Days |
| Allergic reaction 0-14days [8] | 0-14 Days |
| Accelerated reaction 24hours [8] | 1 Day |
| Myocarditis and Pericarditis 0-28days [9] | 0-28 Days |
| Arrhythmia 0-14days [10] | 0-14 Days |
| thrombosis 0-28days[11] | 0-28 Days |
| Arthralgia 21days[12] | 21 Days |
| Respiratory symptoms 0-28days [13] | 0-28 Days |
| Neurological outcome [14] | 0-42 Days |
| Muscle ache and Myalgia 0-7days [15,16] | 0-7 Days |
| GI symptoms 0-14days [17] | 0-14 Days |

**Table S5.Characteristics**

| Author | Refrences | Study design | measured outcome | Series of vaccine dose | Population, Domain | brand | type_vaccine | quality check |
|---|---|---|---|---|---|---|---|---|
| Ali, Bremen et al, 2021 | 18 | Trial | arthralgia, lymphadenopathy, myalgia, fatigue, gastrointestinal | 1 | Children | Moderna | mRNA | Lowrisk |
| Frater et al, 2021 | 19 | Trial | arthralgia, myalgia, fatigue, gastrointestinal symptoms, fever | 1 | HIV | AZ | Viral Vector | Highrisk |
| Frenck et al, 2021 | 20 | Trial | arthralgia, myalgia, fatigue, gastrointestinal symptoms,fever | 1 | Children | Pfizer | mRNA | Some concerns |
| Oosting S F, 2021 | 21 | Trial | cardiac event and, or cardiac symptoms, thromboembolism, | UNS | Cancer | Moderna | mRNA | Lowrisk |
| Sampaio-Barros,et,al, 2021 | 22 | Trial | arthralgia, myalgia, fatigue, fever, respiratory symptoms, gastrointestinal symptoms, | 1 | Autoimmune disease | CV* | Inactivated | Highrisk |
| Shinjo,et,al, 2021 | 23 | Trial | arthralgia, gastrointestinal symptoms, myalgia, fever, fatigue, neurological events | 1 | Autoimmune disease | CV | Inactivated | Highrisk |
| Thomas,et,al,2021 | 24 | Trial | arthralgia, myalgia, fatigue, gastrointestinal symptoms, death | UNS | Cancer | Pfizer | mRNA | Lowrisk |
| Walter,et,al,2022 | 25 | Trial | arthralgia, myalgia, fatigue, gastrointestinal symptoms, fever | 1 | Children | Pfizer | mRNA | Lowrisk |
| Bergman, et al ,2021 | 135 | Trial | serious adverse reaction | 1 and 2 | HIV, Cancer, stem cell transplantation, | Pfizer | mRNA | Highrisk |
| Alamer et al, 2021 | 26 | Cross-sectional | cardiac event, cardiac symptoms, respiratory symptoms, hospitalization, | 1 | Children | Pfizer | mRNA | 2 |
| Ali et al, 2021 | 27 | Cross-sectional | fatigue,fever | 1 | Children, Transplant recepient | Pfizer, Moderna | mRNA | 3 |
| Alonso et al, 2021 | 28 | Cross-sectional | myalgia, fatigue, gastrointestinal symptoms, fever | UNS | MS* | UNS | Inactivated, Viral Vector | 2 |
| Bartels et al, 2021 | 29 | Cross-sectional | arthralgia, myalgia, fatigue, fever | 2 | Autoimmune rheumatic diseases | Pfizer | mRNA | 6 |
| Botwin et al, 2021 | 30 | Cross-sectional | fatigue,gastrointestinal symptoms, fever | 1 | inflammatory bowel disease | Pfizer, Moderna | mRNA | 6 |
| Briggs et al, 2022 | 31 | Cross-sectional | arthralgia, accelerated allergic reaction, myalgia, fatigue, gastrointestinal | 1 | MS | Pfizer, AZ*, JnJ* | mRNA,Viral Vector | 6 |
| Brko et al, 2021 | 32 | Cross-sectional | accelerated allergic reaction, myalgia, fever | 1and 2 | Cancer | Pfizer,Sino*,Sputnik, AZ | mRNA,Viral Vector, Inactivated | 2 |
| Cherian et al, 2021 | 33 | Cross-sectional | arthralgia, myalgia, fatigue, fever | 1 | rheumatoid disease | AZ | Viral Vector | 4 |
| Clayton et al, 2021 | 34 | Cross-sectional | accelerated allergic reaction , disease flare-up, fatigue, fever, | UNS | Darvet syndrome | Pfizer,AZ | mRNA,Viral Vector | 2 |
| Ellul et al, 2021 | 35 | Cross-sectional | arthralgia, lymphadenopathy , fever,gastrointestinal symptoms, myalgia, | 1 | inflammatory bowel disease | Pfizer, Moderna, AZ | mRNA,Viral Vector | 5 |
| Esquivel-Valerio et, 2021 | 36 | Cross-sectional | respiratory symptoms, myalgia, fatigue, fever, gastrointestinal symptoms | 1 | Autoimmune rheumatic diseases | Moderna | mRNA | 1 |
| Foster et al, 2021 | 37 | Cross-sectional | arthralgia, myalgia, fatigue, gastrointestinal symptoms, fever, Vertigo, dizziness | 1 | Cancer | Pfizer, Moderna | mRNA | 5 |
| Fragoulis et al, 2021 | 38 | Cross-sectional | lymphadenopathy, cardiac event and,or cardiac symptoms, respiratory | 1and 2 | Autoimmune disease | AZ | Viral Vector | 8 |
| Haslak et al, 2022 | 39 | Cross-sectional | arthralgia, cardiac event and, or cardiac symptoms, respiratory symptoms, | UNS | inflammatory rheumatic diseases | Pfizer, inactivated | mRNA, inactivated | 3 |

| | | | | | | | | |
|---|---|---|---|---|---|---|---|---|
| K. Allen-Philbey et al, 2021 | 40 | Cross-sectional | disease flare-up, fatigue | 1 | MS | Pfizer, AZ | mRNA,Viral Vector | 2 |
| Kadali R A. K., 2021 | 41 | Cross-sectional | arthralgia, myalgia, fatigue, gastrointestinal symptoms, fever | UNS | Pregnant | Pfizer, Moderna | mRNA | 5 |
| Lechosa-Muniz C., 2021 | 42 | Cross-sectional | lymphadenopathy, gastrointestinal symptoms, fever | UNS | Lactating | Pfizer, Moderna, AZ | mRNA, Inactivated | 7 |
| Liu Y., 2021 | 43 | Cross-sectional | fatigue, fever | UNS | HIV | Sinovac | Inactivated | 7 |
| Lotan I., 2021 | 44 | Cross-sectional | myalgia, fatigue, fever,Vertigo, dizziness | 1 | rare neuroimmunolo-gical diseases | Pfizer, Moderna, AZ, JnJ | mRNA,Viral Vector | 6 |
| Lotan I., 2021 | 45 | Cross-sectional | disease flare-up | UNS | MS | Pfizer | mRNA | 4 |
| McLaurin-Jiang S., 2021 | 46 | Cross-sectional | accelerated allergic reaction, myalgia, fatigue | 1 | lactating | Pfizer, Moderna | mRNA | 7 |
| Orfanoudaki E., 2022 | 47 | Cross-sectional | accelerated allergic reaction, lymphadenopathy, gastrointestinal symptoms, | 1 | inflammatory bowel disease | Pfizer, Moderna, AZ, JnJ | mRNA,Viral Vector | 7 |
| Rotondo,et,al,,2021 | 48 | Cross-sectional | disease flare-up | 1 | Autoimmune disease | Pfizer | mRNA | 4 |
| Sattui,et,al,,2021 | 49 | Cross-sectional | accelerated allergic reaction, fatigue, fever, gastrointestinal symptoms | UNS | Autoimmune disease | Pfizer, Moderna, AZ, JnJ | mRNA,Viral Vector | 2 |
| Shapiro,et,al,,2021 | 50 | Cross-sectional | arthralgia, gastrointestinal symptoms, myalgia, fever, fatigue, lymphadenopathy, | 3 | Immunocompromised | Pfizer | mRNA | 4 |
| Shimabukuro,et,al,,2021 | 51 | Cross-sectional | arthralgia, myalgia, fatigue, fever, gastrointestinal symptoms , SGA*, stillbirth, | 1 and UNS | Pregnant | Pfizer | mRNA | 3 |
| Wang,et,al,,2021 | 52 | Cross-sectional | arthralgia,disease flare-up, gastrointestinal symptoms, fatigue, fever, ear, nose, | UNS | Autoimmune disease | UNS | Inactivated | 2 |
| Zdanowsky,et,al,, 2022 | 53 | Cross-sectional | myalgia, fatigue, fever, gastrointestinal symptoms | 1 | Pregnant | Pfizer | mRNA | 2 |
| Li M 2021 | 54 | Cohort | cardiac event and,or cardiac symptoms | UNS | Children | Pfizer Moderna, JnJ | mRNA,Viral Vector | 5 |
| Achiron et al, 2021 | 55 | Cohort | neurological events and, or symptoms, disease flare-up, death, fatigue, fever | 1and 2 | MS | Pfizer | mRNA | 4 |
| Aharon et al, 2022 | 56 | cohort | biochemical-pregnancy-loss | 2 | Pregnant | Pfizer, Moderna | mRNA | 9 |
| Ariamanesha et al, 2021 | 57 | Cohort | myalgia,fatigue,gastrointestinal symptoms,fever | 2 | Cancer | Sino | Inactivated | 6 |
| Avivi et al, 2021 | 58 | Cohort | arthralgia,lymphadenopathy, myalgia, fatigue, fever,Vertigo, dizziness | 2 | multiple myeloma | Pfizer | mRNA | 7 |
| Benjamini et al, 2022 | 59 | Cohort | myalgia,fever | 2 | Cancer | Pfizer, Moderna, AZ | mRNA,Viral Vector | 6 |
| Blakeway et al, 2022 | 3 | Cohort | small for gestational age, neonatal hospitalization , stillbirth, fever | UNS | Pregnant | UNS | mRNA,Viral Vector | 8 |
| Cavanna et al, 2021 | 60 | Cohort | myalgia, fever | 1and 2 | Cancer | Pfizer, Moderna | mRNA | 8 |
| Chua et al, 2021 | 61 | Cohort | cardiac event and,or cardiac symptoms | 2 | Adolescent | Comirnaty | mRNA | 5 |
| Citu et al, 2022 | 62 | Cohort | arthralgia, lymphadenopathy , fever, fatigue, myalgia | UNS | Pregnant | Pfizer, JnJ | mRNA,Viral Vector | 8 |
| Collier et al, 2021 | 63 | Cohort | fever | 1 | Pregnant | Pfizer, Moderna | mRNA | 7 |

| | | | | | | | | |
|---|---|---|---|---|---|---|---|---|
| Di Noia et al, 2021 | 64 | Cohort | fatigue, gastrointestinal symptoms, fever | 1 | Cancer | Pfizer | mRNA | 8 |
| Dreyer-Alste et al, 2022 | 65 | Cohort | disease flare-up,fatigue,gastrointestinal , symptoms,fever,neurological | 3 | MS | Pfizer | mRNA | 5 |
| Edelman-Klapper et al, 2022 | 66 | Cohort | arthralgia, accelerated allergic reaction, myalgia, fatigue, gastrointestinal | 1and 2 | inflammatory bowel disease | Pfizer | mRNA | 7 |
| Erol et al, 2021 | 67 | Cohort | accelerated allergic reaction, cardiac event and, or cardiac symptoms, fatigue, | 2 | Cancer, Transplant recepient | Sinovac | Inactivated | 7 |
| Furer et al, 2021 | 68 | Cohort | lymphadenopathy, cardiac event and, or cardiac symptoms, respiratory | 1and 2 | Autoimmune rheumatic diseases | Pfizer | mRNA | 7 |
| Golan et al, 2021 | 69 | Cohort | arthralgia, myalgia, fatigue, gastrointestinal symptoms, fever | 1 | lactating | Pfizer, Moderna | mRNA | 4 |
| Goldshtein et al, 2021 | 70 | Cohort | stillbirth, gestational hypertension, abortion, death, SGA, pulmonary | 1 | Pregnant | Pfizer | mRNA | 9 |
| Goldshtein et al, 2022 | 71 | Cohort | preterm birth, SGA, low birth weight, neonatal hospitalization | 1 | Pregnant | Pfizer | mRNA | 9 |
| Goshen-Lago et al, 2021 | 72 | Cohort | lymphadenopathy, fatigue, gastrointestinal symptoms, fever | UNS | Cancer | Pfizer | mRNA | 4 |
| Gray et al, 2021 | 73 | Cohort | accelerated allergic reaction, myalgia, fatigue, fever | 1and 2 | lactating and pregnant | Pfizer, Moderna | mRNA | 6 |
| Greenberg et al, 2021 | 74 | Cohort | myalgia, fatigue, fever, ear, nose, throat symptoms | 1and 2 | multiple myeloma ,Kidn ey transplant | Pfizer, Moderna | mRNA | |
| Hadi et al, 2021 | 75 | Cohort | accelerated allergic reaction | UNS | inflammatory bowel disease | Pfizer, Moderna | mRNA | 6 |
| Hall et al, 2021 | 76 | Cohort | arthralgia, myalgia, fatigue, gastrointestinal symptoms, fever | 1 | Cancer, Transplant recepient | Moderna | mRNA | 7 |
| Heshin-Bekenstein Eet al, 2022 | 77 | Cohort | arthralgia. accelerated allergic reaction, disease flare-up, hospitalization, | 1and 2 | rheumatoid disease | Pfizer | mRNA | |
| Hod et al, 2021 | 78 | Cohort | myalgia, fatigue, gastrointestinal symptoms, fever | 1 | Transplant recepient | Pfizer | mRNA | 8 |
| Jovicevic et al, 2022 | 79 | Cohort | disease flare-up | 2 | Neuromyelitis optica spectrum disorders | Sino | Inactivated | 2 |
| June Choe., 2022 | 80 | Cohort | accelerated allergic reaction, cardiac event and, or cardiac symptoms, fatigue, | 1 | Children | Pfizer | mRNA | 4 |
| Jyssum I., 2021 | 81 | Cohort | accelerated allergic reaction,respiratory symptoms, myalgia, fever, | 1 | rheumatoid arthritis | Pfizer, Moderna, AZ | mRNA, Inactivated | 9 |
| Karacin C, 2021 | 82 | Cohort | myalgia, fatigue, fever | 1 | Cancer | CV | Inactivated | 6 |
| Kian W., 2022 | 83 | Cohort | lymphadenopathy, respiratory symptoms, myalgia, fatigue, fever | 1and 2 | Children | Pfizer | mRNA | 7 |
| Lasagna A., 2021 | 84 | Cohort | arthralgia, myalgia, fever | UNS | Cancer | Pfizer | mRNA | 8 |
| Levy I., 2021 | 85 | Cohort | arthralgia, cardiac event and,or cardiac symptoms, respiratory symptoms, | 1 | HIV | Pfizer | mRNA | 7 |
| Ligumsky H., 2022 | 86 | Cohort | myalgia | 2 | Cancer | Pfizer | mRNA | 9 |
| Linardou H., 2021 | 87 | Cohort | fatigue | 2 | Cancer | Pfizer, Moderna, AZ | mRNA,Viral Vector | 9 |
| Liontos M., 2021 | 88 | Cohort | fatigue, fever | UNS | Cancer | Pfizer, Moderna | mRNA | 8 |

| | | | | | | | | |
|---|---|---|---|---|---|---|---|---|
| Low J M., 2022 | 89 | Cohort | fatigue, gastrointestinal symptoms, fever, lymphadenopathy | 2 | Lactating | Pfizer | mRNA | 5 |
| Ma Y., 2021 | 90 | Cohort | arthralgia | UNS | Cancer,Autoimmune disease | Sino, SinoVac | Inactivated | 8 |
| Machado P M., 2021 | 91 | Cohort | fatigue, gastrointestinal symptoms, fever | UNS | autoimmune rheumatic and musculoskeletal | Pfizer, Moderna, AZ | mRNA, Viral Vector | 8 |
| Massa F., 2021 | 92 | Cohort | myalgia ,fatigue, fever, hypertension crisis, gastrointestinal | 1and 2 | Transplant recepient | Pfizer | mRNA | 6 |
| Matkowska-Kocjan A, 2021 | 93 | Cohort | lymphadenopathy, gastrointestinal symptoms, fever | 1 | Cancer,Transplant recepient | Pfizer | mRNA | 6 |
| Medeiros-Ribeiro A C, 2020 | 94 | Cohort | gastrointestinal symptoms, respiratory symptoms, arthralgia, myalgia, fatigue, | 1and 2 | Autoimmune rheumatic diseases | CV | Inactivated | 9 |
| Medina-Pestana J., 2021 | 95 | Cohort | myalgia, fever | UNS | Transplant recepient | UNS | Inactivated | 9 |
| Nakahara A., 2022 | 96 | Cohort | cardiac event and,or cardiac symptoms,respiratory symptoms, fatigue, fever, | UNS | Pregnant, cancer | Pfizer, Moderna | mRNA | 8 |
| Oster M E., 2022 | 97 | Cohort | cardiac event and, or cardiac symptoms | UNS | Children | Pfizer | mRNA | 6 |
| Palaia I., 2021 | 98 | Cohort | gastrointestinal symptoms, fever | 1 | Cancer | Pfizer | mRNA | 8 |
| Peet C J., 2021 | 99 | Cohort | myalgia, fever, fatigue | UNS | systemic autoinflammato-ry disease | Pfizer, AZ | mRNA,Viral Vector | 5 |
| Peled Y., 2021 | 100 | Cohort | fatigue, gastrointestinal symptoms, fever | 1and 3 | Transplant recepient | Pfizer | mRNA | 6 |
| Pham,et,al,,2021 | 101 | Cohort | disease flare-up,myalgia,fatigue,gastrointestinal | UNS | Immunocompro-mised | UNS | mRNA | 4 |
| Pimpinelli,et,al,,2021 | 102 | Cohort | myalgia, fatigue, fever | 1 | Cancer | Pfizer | mRNA | 3 |
| Pinte,et,al,,2021 | 103 | Cohort | disease flare-up | UNS | Autoimmune disease | Pfizer, Moderna, AZ, JnJ | mRNA,Viral Vector | 4 |
| Rabinowich,et,al,,2021 | 104 | Cohort | accelerated allergic reaction,neurological events and,or symptoms | 2 | Cancer, Transplant recepient | Pfizer | mRNA | 4 |
| Revon-Riviere,et,al,,2021 | 105 | Cohort | gastrointestinal symptoms,fever | 1 | Children, Cancer | Pfizer | mRNA | 3 |
| Rottenstreich,et,al,,2021 | 106 | Cohort | respiratory symptoms, SGA, neonatal hospitalization, neurological events and, or | UNS | Pregnant | Pfizer | mRNA | 7 |
| Russo,et,al,,2021 | 107 | Cohort | fever | UNS | Transplant recepient | Pfizer | mRNA | 4 |
| Sanders,et,al,,2021 | 108 | Cohort | arthralgia,myalgia,fatigue,fever,gastrointestinal symptoms | 1 | Cancer, Transplant recepient | Moderna | mRNA | 6 |
| Shulman,et,al,,2022 | 109 | Cohort | arthralgia, accelerated allergic reaction, myalgia, fatigue, fever, | 1and 2 | Cancer | Pfizer | mRNA | 4 |
| So,et,al,,2021 | 110 | Cohort | arthralgia, fatigue, fever, lymphadenopathy, gastrointestinal symptoms | 1 | Cancer | Pfizer, Moderna, AZ | mRNA, Viral Vector | 5 |
| Spinelli,et,al,,2022 | 111 | Cohort | arthralgia, fatigue, fever | 1 | Autoimmune disease | UNS | mRNA | 6 |
| Strobel,et,al,,2021 | 112 | Cohort | arthralgia, myalgia, fatigue,fever, gastrointestinal symptoms, neurological | 1 | Cancer | Pfizer, Moderna, AZ, JnJ | mRNA, Viral Vector | 4 |
| Tamura,et,al,,2021 | 113 | Cohort | myalgia, fatigue, fever | 1and 2 | Cancer | Pfizer, Moderna | mRNA, unknown | 5 |

| | | | | | | | | |
|---|---|---|---|---|---|---|---|---|
| Tanner,et,al,,2022 | 114 | Cohort | fatigue | 2 | Transplant recepient | AZ | Viral Vector | 3 |
| Theiler,et,al,,2021 | 115 | Cohort | neonatal hospitalization ,stillbirth, gestational hypertension, | UNS | Pregnant | Pfizer, Moderna, JnJ | mRNA,Viral Vector | 2 |
| Trillo,Aliaga,et,al, ,2021 | 116 | Cohort | arthralgia, lymphadenopathy, myalgia, fatigue, fever, | 1 | Cancer | Pfizer, Moderna, AZ | mRNA,Viral Vector | 5 |
| Tsimafeyeu,et,al,2 021 | 117 | Cohort | fever | 2 | Cancer | Sputnik | Viral Vector | 6 |
| Tzioufas,et,al,,20 21 | 118 | Cohort | accelerated allergic reaction, lymphadenopathy, disease flare-up, fatigue, | 2 | Autoimmune disease | UNS | mRNA | 5 |
| Valentini,et,al,,20 22 | 119 | Cohort | respiratory symptoms, myalgia, fatigue, fever, Vertigo, dizziness | 1 | Down syndrome | Pfizer | mRNA | 4 |
| Wainstock,et,al,2 021 | 120 | Cohort | small for gestational age, gestational hypertension | 1 | Pregnant | Pfizer | mRNA | 7 |
| Watcharananan,et ,al,,2022 | 121 | Cohort | cardiac event and, or cardiac symptoms, fatigue, fever, gastrointestinal symptoms, | 1 | Cancer, Transplant recepient | AZ | Viral Vector | 4 |
| Watanabe,et,al,20 22 | 122 | Cohort | arthralgia, myalgia, fatigue, gastrointestinal symptoms, fever | 1 | Cancer, Transplant recepient | Pfizer | mRNA | 5 |
| Weaver,at,al,2021 | 123 | Cohort | arthralgia accelerated allergic reaction, disease flare-up, myalgia, fatigue, | 1and 2 | Autoimmune disease | Pfizer | mRNA | 4 |
| Yasin,et,al,2022 | 124 | Cohort | myalgia, fatigue, fever, gastrointestinal symptoms | 1 | Cancer | CV | Inactivated | 4 |
| Zagouri,et,al,2021 | 125 | Cohort | arthralgia, fatigue | 1 | Cancer | Pfizer | mRNA | 4 |
| Zauche,et,al,2021 | 126 | cohort | abortion | UNS | Pregnant | UNS | mRNA | 5 |
| Bernstine et al, 2021 | 127 | cohort | lymphadenopathy | 1and 2 | Cancer | Pfizer | mRNA | 7 |
| Bleicher et al, 2021 | 128 | cohort | SGA, pregnancy loss, other maternal complication | UNS | Pregnant | Pfizer | mRNA | 5 |
| Mark C., 2021 | 129 | cohort | allergic reaction | 1 | Leukemia | Pfizer | mRNA | 5 |
| Naranbhai V., 2022 | 130 | cohort | fatigue, myalgia, gastrointestinal symptoms | 1and 2 | Cancer | Pfizer, Moderna | mRNA | 9 |
| Peled Y., 2022 | 131 | cohort | arthralgia, fatigue, myalgia, gastrointestinal symptoms | 1 and 2 and 3 | Transplant recepient | Pfizer | mRNA | 6 |
| Skroza,et,al,,2021 | 132 | cohort | disease flare-up | UNS | Autoimmune disease | Pfizer | mRNA | 5 |
| Aikawa et al, 2021 | 133 | Case-control | arthralgia, respiratory symptoms, fatigue, fever, conjunctivitis, ear, nose, and | 1and 2 | Autoimmune rheumatic diseases | CV | Inactivated | 8 |
| Bookstein Peretz et al, 2021 | 134 | Case-control | arthralgia, lymphadenopathy , fever, neurological events and, or symptoms | 1 | Pregnant | Pfizer | mRNA | 8 |

UNS* : Unspecified

SGA*: Small for gestational age

AZ*: AZ

CV*: CV

JnJ*: JnJ

Sino*: SinoPharm

MS*: Multiple sclerosis

### Table S6.Characteristics Frequency

| Variable | Category | Percentage.Freq (%) |
|---|---|---|
| Publication Year | 2021 | 80.65 |
| | 2022 | 19.35 |
| Population_group | Autoimmune disease | 24.1 |
| | Cancer | 24.1 |
| | Immunocompromised | 20 |
| | Pregnant | 13.3 |
| | Children and adolescent | 7.5 |
| | Multiple sclerosis | 5 |
| | lactating | 4.1 |
| | Disability(Down syndrom / Darvet syndrome) | 1.6 |
| Study design | Cohort | 67.8 |
| | Cross-sectional | 25 |
| | Clinical Trial | 6.6 |
| | Case-control | 1.6 |
| type_vaccine | mRNA | 64.1 |
| | reported together | 20 |
| | inactivated | 10.8 |
| | viralvector | 5 |
| Vaccine brand | pfizer | 40.8 |
| | reported together | 41.6 |
| | Moderna | 4.1 |
| | CoronaVac | 5 |
| | AstraZeneca | 4.1 |
| | SinoVac | 0.8 |
| | SinoPharm | 1.6 |
| | Comirnaty | 0.8 |

| | | |
|---|---|---|
| | Sputnik | 0.8 |
| Measeured outcome | Fever | 17.07 |
| | Fatigue | 16.26 |
| | GI | 12.52 |
| | Myalgia | 12.03 |
| | Arthralgia | 7.64 |
| | Allergic | 4.72 |
| | Lymphadenopathy | 3.58 |
| | Neurological | 3.58 |
| | Respiratory symptoms | 3.58 |
| | Deterioration | 3.25 |
| | Vertigo | 3.25 |
| | Cardiac symtoms and events | 3.09 |
| | ENT | 2.76 |
| | Pregnancy loss and abortion | 0.98 |
| | SGA | 0.81 |
| | Death | 0.65 |
| | Eye complication | 0.65 |
| | Hospitalization | 0.65 |
| | Hypertensive | 0.65 |
| | Neonatal Hospitalization | 0.65 |
| | Stillbirth | 0.65 |
| | Gestational hypertension | 0.49 |
| | Thromboembolism | 0.49 |
| age_group | Adults (18-50) | 85.4 |
| | Older adults (>50) | 8.8 |
| | Children and Adolescents (<18 years) | 5.8 |
| Income_Status | High income | 70.41 |

| | | |
|---|---|---|
| | Upper middle income | 22.28 |
| | Other | 6.02 |
| | Lower middle income | 1.3 |
| Country | USA | 21.14 |
| | Isreal | 15.77 |
| | Greece | 8.78 |
| | Brazil | 5.69 |
| | Turkey | 5.69 |
| | Italy | 5.37 |
| | Mexico | 5.2 |
| | UK | 4.39 |
| | Germany | 4.07 |
| | global | 3.41 |
| | China | 2.28 |
| | Serbia | 2.28 |
| | LatinAmerica | 1.79 |
| | Netherland | 1.46 |
| | France | 1.3 |
| | Japan | 1.3 |
| | EU | 1.14 |
| | Poland | 1.14 |
| | KSA | 0.98 |
| | Romania | 0.81 |
| | Thailand | 0.81 |
| | Argentina,Brazil,SouthAfrica,USA | 0.65 |
| | Denmark | 0.65 |
| | India | 0.65 |
| | Iran | 0.65 |

| | | |
|---|---|---|
| | South Korea | 0.65 |
| | Singapore | 0.49 |
| | Spain | 0.49 |
| | Canada | 0.33 |
| | Asia | 0.16 |
| | Hong kong | 0.16 |
| | Ireland | 0.16 |
| | Russia | 0.16 |
| number of injection | Not specified | 28.3 |
| | 1 | 55.83 |
| | 2 | 27.5 |
| | 3 | 2.5 |

### Table S7.Sever Outcomes

| Design | Dose | Population | vaccine | OR | IR | IP | low | high | I2 | P | Outcome |
|---|---|---|---|---|---|---|---|---|---|---|---|
| Case-Control | 1 | Autoimmune | Inactivated | | | 42.60 | 0.08 | 1000 | 92% | < 0.01 | Respiratory symptoms |
| Case-Control | 2 | Autoimmune | Inactivated | | | 37.83 | 0.15 | 1001 | 88% | < 0.01 | Respiratory symptoms |
| Case-Control | 1 | Pregnant | mRNA | | | 23.08 | 10.61 | 43.35 | | | Neurological symptoms |
| Case-Control | 1 | Autoimmune | Inactivated | | | 25.48 | 14.63 | 41.05 | | | Neurological symptoms |
| Case-Control | 2 | Pregnant | mRNA | | | 46.15 | 27.58 | 71.96 | | | Neurological symptoms |
| Case-Control | 2 | Autoimmune | Inactivated | | | 15.92 | 7.66 | 29.09 | | | Neurological symptoms |
| Case-Control | 2 | | | | | 27.69 | 0.03 | 1000 | 87% | <0.01 | Neurological symptoms |
| Case-Control | 1 | | | | | 24.59 | 13.45 | 44.95 | 0 | 0.81 | Neurological symptoms |
| Case-Control | 2 | Pregnant | mRNA | | 2.93 | | 0.90 | 4.96 | | | Lymphadenopathy |
| Case-Control | 1 | Pregnant | mRNA | | 0.37 | | 0.00 | 1.08 | | | Lymphadenopathy |
| Case-Control | 1 | Pregnant | mRNA | | | 2.56 | 0.06 | 14.20 | | | Lymphadenopathy |
| Case-Control | 2 | Pregnant | mRNA | | | 20.51 | 8.90 | 40.02 | | | Lymphadenopathy |
| Cohort | 1 | Pregnant | mRNA, viralvector | | | 0.00 | 0.00 | 24.71 | | | Thromboembolism |
| Cohort | any | Pregnant | | | | 3.65 | 2.64 | 5.05 | 0 | 0.98 | Stillbirth |
| Cohort | 1 | Pregnant | | | | 0.13 | 0.00 | 0.74 | | | Stillbirth |
| Cohort | any | Pregnant | | 2.05 | | | 0.18 | 23.25 | 0 | 0.62 | Stillbirth |
| Cohort | any | | | | | 73.18 | 9.67 | 553.68 | 95% | <0.01 | SGA |
| Cohort | 1 | | mRNA | | | 62.93 | 44.77 | 88.45 | 0 | 0.37 | SGA |
| Cohort | any | | mRNA | | | 57.46 | 0.01 | 1000 | 97% | <0.01 | SGA |
| Cohort | 2 | | mRNA | | | 25.07 | 15.16 | 38.87 | | | SGA |
| Cohort | 1 | | mRNA | 0.66 | | | 0.4 | 1.07 | | | SGA |
| Cohort | 1 | | mRNA | 0.97 | | | 0.87 | 1.08 | | | SGA |
| Cohort | 2 | | mRNA | 1.21 | | | 0.56 | 2.64 | | | SGA |
| Cohort | any | | | 1.02 | | | 0.82 | 1.26 | 21% | 0.28 | SGA |
| Cohort | 1 | Disable | mRNA | | 3.57 | | 0 | 10.57 | | | Respiratory symptoms |
| Cohort | 1 | | mRNA | | 1.73 | | 0.28 | 10.41 | 0 | 0.42 | Respiratory symptoms |
| Cohort | 1 | IMC* | mRNA | | 1.07 | | 0 | 2.28 | | | Respiratory symptoms |

| | | | | | | | | | | |
|---|---|---|---|---|---|---|---|---|---|---|
| Cohort | 2 | | mRNA | 0.06 | | 0 | 0.19 | 0 | 0.69 | Respiratory symptoms |
| Cohort | 2 | Autoimmune | mRNA | 0.05 | | 0 | 0.15 | | | Respiratory symptoms |
| Cohort | 2 | IMC | mRNA | 0 | | 0 | 0.74 | | | Respiratory symptoms |
| Cohort | 2 | Disable | mRNA | 0 | | 0 | 6.74 | | | Respiratory symptoms |
| Cohort | 1 | Autoimmune | | | 20.71 | 0.39 | 1000 | 92% | < 0.01 | Respiratory symptoms |
| Cohort | 2 | Autoimmune | | | 15.89 | 0.15 | 1000 | 92% | < 0.01 | Respiratory symptoms |
| Cohort | 1 | Disable | | | 25 | 0.63 | 131.5 | | | Respiratory symptoms |
| Cohort | 1 | IMC | | | 22.56 | 4.68 | 64.5 | | | Respiratory symptoms |
| Cohort | 2 | Disable | | | 0 | 0 | 88.10 | | | Respiratory symptoms |
| Cohort | 2 | IMC | | | 0 | 0 | 30.03 | | | Respiratory symptoms |
| Cohort | 2 | Children/ Adolescents | | | 9.52 | 1.16 | 33.98 | | | Respiratory symptoms |
| Cohort | 1 | | Inactivated | | 47.54 | 0.34 | 1000 | 92% | < 0.01 | Respiratory symptoms |
| Cohort | 1 | | mRNA | | 13.24 | 2.34 | 75.5 | 52% | 0.1 | Respiratory symptoms |
| Cohort | 2 | | Inactivated | | 40.36 | 0.13 | 1000 | 93% | < 0.01 | Respiratory symptoms |
| Cohort | 2 | | mRNA | | 5.56 | 1.22 | 25.34 | 0 | 0.43 | Respiratory symptoms |
| Cohort | 1 | | | | 24.11 | 7.91 | 73.47 | 85% | < 0.01 | Respiratory symptoms |
| Cohort | 2 | | | | 14.10 | 3.42 | 58.20 | 86% | < 0.01 | Respiratory symptoms |
| Cohort | 2 | IMC | | | 38.51 | 0.02 | 1000 | 0 | 0.44 | Neurological symptoms |
| Cohort | 1 | MS* | mRNA | | 5.41 | 1.12 | 15.71 | | | Neurological symptoms |
| Cohort | 1 | IMC | Viralvector | | 24.10 | 2.93 | 84.35 | | | Neurological symptoms |
| Cohort | 2 | MS* | | | 11.49 | 3.74 | 26.62 | | | Neurological symptoms |
| Cohort | 2 | | Inactivated | | 0.00 | 0.00 | 112.19 | | | Neurological symptoms |
| Cohort | 2 | | mRNA | | 20.71 | 0.00 | 1000 | 57% | 0.13 | Neurological symptoms |
| Cohort | 2 | | | | 16.43 | 2.32 | 116.19 | 15% | 0.31 | Neurological symptoms |
| Cohort | 1 | | | | 10.83 | 0.00 | 1000 | 63% | 0.10 | Neurological symptoms |
| Cohort | 2 | Cancer | | | 5.34 | 1.69 | 16.90 | 0 | 0.93 | Neurological event |
| Cohort | 1 | Cancer | | | 5.05 | 1.40 | 18.19 | 0 | 0.92 | Neurological event |
| Cohort | 2 | IMC | mRNA | | 0.00 | 0.00 | 45.06 | | | Neurological event |
| Cohort | Any | Cancer | | | 0.00 | 0.00 | 40.60 | | | Neurological event |

| | | | | | | | | | | |
|---|---|---|---|---|---|---|---|---|---|---|
| Cohort | Any | Pregnant | mRNA | | 1.40 | 0.04 | 7.80 | | | Neurological event |
| Cohort | Any | | | | 2.81 | 0.34 | 22.92 | 0 | 0.62 | Neurological event |
| Cohort | 2 | | | | 5.62 | 4.11 | 7.68 | 0 | 0.99 | Neurological event |
| Cohort | any | | mRNA | | 40.73 | 27.44 | 57.97 | | | Neonatal hospitalization |
| Cohort | any | | | | 33.89 | 4.10 | 280.14 | 43% | 0.17 | Neonatal hospitalization |
| Cohort | 2 | | | 1.95 | | 1.20 | 3.19 | 0 | 0.94 | Lymphadenopathy |
| Cohort | 2 | | mRNA | 1.82 | | 0.00 | 5.74 | 0 | 0.77 | Lymphadenopathy |
| Cohort | 1 | Cancer | mRNA | 4.39 | | 0.00 | 10.48 | | | Lymphadenopathy |
| Cohort | 2 | Cancer | | 1.80 | | 0.00 | 3.67 | 0 | 0.81 | Lymphadenopathy |
| Cohort | 2 | IMC | | 1.67 | | 0.00 | 3.99 | | | Lymphadenopathy |
| Cohort | 1 | Cancer | | 0.77 | | 0.00 | 1.66 | 0 | 0.70 | Lymphadenopathy |
| Cohort | 1 | Cancer | mRNA | | 30.77 | 3.75 | 106.77 | | | Lymphadenopathy |
| Cohort | 2 | | mRNA | | 11.91 | 3.89 | 36.47 | 70% | <0.01 | Lymphadenopathy |
| Cohort | 2 | Autoimmune | | | 6.16 | 0.00 | 1000 | 56% | 0.13 | Lymphadenopathy |
| Cohort | 2 | Cancer | | | 15.53 | 10.54 | 22.88 | 0 | 0.97 | Lymphadenopathy |
| Cohort | 1 | Cancer | | | 10.39 | 4.35 | 24.83 | 0 | 0.54 | Lymphadenopathy |
| Cohort | 2 | Pregnant | | | 54.35 | 17.88 | 122.29 | | | Lymphadenopathy |
| Cohort | 2 | Children/ Adolescents | | | 4.76 | 0.12 | 26.24 | | | Lymphadenopathy |
| Cohort | 2 | IMC | | | 11.70 | 1.42 | 41.61 | | | Lymphadenopathy |
| Cohort | 2 | | | | 13.44 | 7.19 | 25.12 | 48% | 0.05 | Lymphadenopathy |
| Cohort | 1 | IMC | mRNA | | 16.39 | 0.41 | 87.99 | | | Hypertensive disorder |
| Cohort | 2 | IMC | | | 18.71 | 8.84 | 39.59 | 0 | 0.94 | Hypertensive disorder |
| Cohort | 2 | Autoimmune | | | 3.31 | 0.40 | 11.89 | | | Hypertensive disorder |
| Cohort | 2 | | Inactivated | | 0.00 | 0.00 | 112.19 | | | Hypertensive disorder |
| Cohort | 2 | | mRNA | | 8.19 | 0.49 | 135.66 | 31% | 0.24 | Hypertensive disorder |
| Cohort | 2 | | | | 8.09 | 1.59 | 41.14 | 5% | 0.37 | Hypertensive disorder |
| Cohort | 2 | | mRNA | | 1.19 | 0.00 | 1000 | 94% | <0.01 | Hospitalization |
| Cohort | 2 | Children/ Adolescents | mRNA | | 0.16 | 0.13 | 0.20 | | | Hospitalization |
| Cohort | 2 | Autoimmune | mRNA | | 11.11 | 0.28 | 60.36 | | | Hospitalization |

| | | | | | | | | | | |
|---|---|---|---|---|---|---|---|---|---|---|
| Cohort | 1 | | mRNA | | 1.29 | 0.00 | 1000 | 98% | <0.01 | Hospitalization |
| Cohort | 1 | Children/ Adolescents | mRNA | | 0.08 | 0.06 | 0.11 | | | Hospitalization |
| Cohort | 1 | Autoimmune | mRNA | | 22.22 | 2.70 | 77.98 | | | Hospitalization |
| Cohort | 2 | | mRNA | | 50.13 | 35.72 | 68.16 | | | Gestational hypertension |
| Cohort | any | | | | 36.57 | 0.00 | 1000 | 88% | <0.01 | Gestational hypertension |
| Cohort | 1 | | mRNA | | 14.28 | 0.00 | 1000 | 99% | <0.01 | Gestational hypertension |
| Cohort | 2 | Autoimmune | mRNA | 2.73 | | 0.56 | 4.89 | 97% | <0.01 | Flare up |
| Cohort | 2 | | mRNA | 2.40 | | 0.81 | 3.99 | 96% | <0.01 | Flare up |
| Cohort | 1 | | | 1.13 | | 0.47 | 2.68 | 63% | 0.03 | Flare up |
| Cohort | 2 | MS* | mRNA | 1.42 | | 0.00 | 12.55 | 90% | <0.01 | Flare up |
| Cohort | 1 | Autoimmune | | 0.92 | | 0.00 | 2.01 | 33% | 0.23 | Flare up |
| Cohort | 1 | | Viralvector | 0.88 | | 0.00 | 2.62 | | | Flare up |
| Cohort | 1 | | mRNA | 0.85 | | 0.36 | 1.34 | 18% | 0.30 | Flare up |
| Cohort | 1 | MS* | | 0.79 | | 0.00 | 2.40 | 0 | 0.49 | Flare up |
| Cohort | 2 | Autoimmune | | | 45.01 | 17.28 | 117.24 | 97% | <0.01 | Flare up |
| Cohort | 1 | | Viralvector | | 6.21 | 0.16 | 34.12 | | | Flare up |
| Cohort | 1 | Autoimmune | | | 25.88 | 2.48 | 270.11 | 60% | 0.08 | Flare up |
| Cohort | 2 | | mRNA | | 38.58 | 16.31 | 91.22 | 97% | <0.01 | Flare up |
| Cohort | 2 | MS* | | | 24.84 | 0.00 | 1000 | 88% | <0.01 | Flare up |
| Cohort | 1 | MS* | | | 17.34 | 2.35 | 127.73 | 0 | 0.49 | Flare up |
| Cohort | 1 | | mRNA | | 24.00 | 10.40 | 55.39 | 52% | 0.10 | Flare up |
| Cohort | 2 | | Inactivated | | 0.00 | 0.00 | 369.42 | | | Flare up |
| Cohort | 2 | | | | 39.27 | 18.08 | 85.31 | 96% | <0.01 | Flare up |
| Cohort | 1 | | | | 22.13 | 10.22 | 47.93 | 50% | 0.09 | Flare up |
| Cohort | 1 | | | 0.26 | | 0.006 | 10.58 | 43% | 0.17 | Cardiac symptoms |
| Cohort | 2 | | | 0.75 | | 0.00 | 5175.62 | 0 | 0.39 | Cardiac symptoms |
| Cohort | 2 | IMC | mRNA | 0.46 | | 0.00 | 1.36 | | | Cardiac symptoms |
| Cohort | 1 | | mRNA | 0.11 | | 0.00 | 0.48 | 0 | 0.69 | Cardiac symptoms |
| Cohort | 1 | Autoimmune | | 0.10 | | 0.00 | 0.24 | | | Cardiac symptoms |

| | | | | | | | | | | |
|---|---|---|---|---|---|---|---|---|---|---|
| Cohort | 1 | Cancer | | 0.00 | | 0.00 | 0.57 | | | Cardiac symptoms |
| Cohort | 1 | IMC | | 0.00 | | 0.00 | 0.94 | | | Cardiac symptoms |
| Cohort | 2 | Cancer | Viralvector | 0.00 | | 0.00 | 7.57 | | | Cardiac symptoms |
| Cohort | 1 | | Viralvector | 0.00 | | 0.00 | 7.57 | | | Cardiac symptoms |
| Cohort | 2 | | | | 6.11 | 2.05 | 18.22 | 0 | 0.59 | Cardiac symptoms |
| Cohort | 1 | | | | 3.78 | 2.53 | 5.65 | 0 | 0.99 | Cardiac symptoms |
| Cohort | 2 | | mRNA | | 3.48 | 0.00 | 1000 | 34% | 0.22 | Cardiac symptoms |
| Cohort | 1 | | mRNA | | 3.07 | 0.86 | 10.95 | 0 | 0.87 | Cardiac symptoms |
| Cohort | 2 | Cancer | | | 9.56 | 4.51 | 20.28 | 0 | 0.93 | Cardiac symptoms |
| Cohort | 1 | | Viralvector | | 0.00 | 0.00 | 43.47 | | | Cardiac symptoms |
| Cohort | 1 | Cancer | | | 5.53 | 3.77 | 7.58 | 0 | 0.99 | Cardiac symptoms |
| Cohort | 2 | IMC | | | 8.26 | 0.21 | 45.19 | | | Cardiac symptoms |
| Cohort | 2 | | Viralvector | | 0.00 | 0.00 | 43.47 | | | Cardiac symptoms |
| Cohort | 1 | Autoimmune | | | 2.92 | 0.35 | 10.49 | | | Cardiac symptoms |
| Cohort | 2 | Autoimmune | | | 1.46 | 0.04 | 8.09 | | | Cardiac symptoms |
| Cohort | 1 | IMC | | | 0.00 | 0.00 | 27.35 | | | Cardiac symptoms |
| Cohort | 2 | Children/ Adolescents | mRNA | 0.01 | | 0.01 | 0.01 | | | Cardiac event |
| Cohort | 1 | Children/ Adolescents | mRNA | 0.0024 | | 0.00048 | 0.004 | | | Cardiac event |
| Cohort | 1 | | | | 6.08 | 4.09 | 9.06 | 0 | 0.47 | Allergic reaction |
| Cohort | 2 | | | | 5.61 | 4.18 | 7.53 | 0 | 0.87 | Allergic reaction |
| Cohort | 1 | Lactating | | | 0.00 | 0.00 | 112.19 | | | Allergic reaction |
| Cohort | 1 | Pregnant | | | 0.00 | 0.00 | 42.96 | 0 | 0.62 | Allergic reaction |
| Cohort | 1 | Autoimmune | | | 7.03 | 4.10 | 12.06 | 0 | 0.43 | Allergic reaction |
| Cohort | 2 | Autoimmune | | | 5.53 | 3.60 | 8.48 | 0 | 0.66 | Allergic reaction |
| Cohort | 2 | Cancer | | | 4.87 | 2.21 | 10.76 | 0 | 0.68 | Allergic reaction |
| Cohort | 1 | Cancer | | | 2.68 | 1.01 | 10.00 | 0 | 0.77 | Allergic reaction |
| Cohort | 2 | | mRNA | | 5.53 | 3.93 | 7.77 | 0 | 0.72 | Allergic reaction |
| Cohort | 1 | | mRNA | | 5.39 | 3.02 | 9.62 | 21% | 0.27 | Allergic reaction |
| Cohort | 1 | | Viralvector | | 12.42 | 1.51 | 44.15 | | | Allergic reaction |

| | | | | | | | | | | |
|---|---|---|---|---|---|---|---|---|---|---|
| Cohort | 2 | Lactating | | | 0.00 | 0.00 | 112.19 | | | Allergic reaction |
| Cohort | 2 | Pregnant | | | 11.90 | 0.30 | 64.55 | 0 | 0.87 | Allergic reaction |
| Cohort | 2 | | Inactivated | | 0 | 0 | 112.19 | | | Allergic reaction |
| Cohort | 1 | Pregnant | mRNA | 1.62 | | 1.18 | 2.22 | | | Abortion |
| Cohort | Any | Pregnant | mRNA | 0.17 | | 0.06 | 0.50 | 95% | <0.01 | Abortion |
| Cohort | Any | Pregnant | mRNA | | 8.55 | 3.59 | 20.37 | 96% | <0.01 | Abortion |
| Cohort | 1 | Pregnant | mRNA | | 51.86 | 0 | 1000 | 99% | <0.01 | Abortion |
| Cohort | 2 | | | | | | | | | Cardiac event |
| Cohort | 1 | | | | | | | | | Cardiac event |
| Cross-sectional | 1 | Autoimmune | mRNA, viralvector | | 0.7707 | 0.1927 | 30816.00 | | | Thromboembolism |
| Cross-sectional | 2 | Autoimmune | mRNA, viralvector | | 1156.00 | 0.3728 | 35843.00 | | | Thromboembolism |
| Cross-sectional | 2 | | mRNA | 74.52 | | 0 | 394.91 | 97% | < 0.01 | Respiratory symptoms |
| Cross-sectional | 1 | | mRNA | 60.58 | | 0 | 320.09 | 96% | < 0.01 | Respiratory symptoms |
| Cross-sectional | 2 | Children/ Adolescents | | 227.11 | | 170.58 | 283.64 | | | Respiratory symptoms |
| Cross-sectional | 1 | Children/ Adolescents | | 184.56 | | 135.78 | 233.34 | | | Respiratory symptoms |
| Cross-sectional | 1 | | Viralvector | 1.20 | | 0 | 14.60 | 0 | 0.51 | Respiratory symptoms |
| Cross-sectional | 1 | Autoimmune | | 0.29 | | 0 | 0.81 | 0 | 0.75 | Respiratory symptoms |
| Cross-sectional | 2 | | | 4.13 | | 0.02 | 712.07 | 94% | < 0.01 | Respiratory symptoms |
| Cross-sectional | 2 | Autoimmune | | 0.15 | | 0 | 0.67 | 0 | 0.66 | Respiratory symptoms |
| Cross-sectional | 2 | | Viralvector | 0 | | 0 | 4.29 | | | Respiratory symptoms |
| Cross-sectional | 2 | Autoimmune | | | 8.09 | 0.13 | 498.89 | 28% | 0.25 | Respiratory symptoms |
| Cross-sectional | 1 | Autoimmune | | | 6.11 | 2.59 | 14.45 | 7% | 0.38 | Respiratory symptoms |
| Cross-sectional | 1 | Children/ Adolescents | | | 184.56 | 142.17 | 233.36 | | | Respiratory symptoms |
| Cross-sectional | 2 | Children/ Adolescents | | | 227.11 | 178.78 | 281.46 | | | Respiratory symptoms |
| Cross-sectional | 1 | | | 5.83 | | 0.21 | 158.58 | 94% | < 0.01 | Respiratory symptoms |
| Cross-sectional | 1 | | Inactivated | | 4.44 | 0.11 | 24.51 | | | Respiratory symptoms |
| Cross-sectional | 2 | | Viralvector | | 0 | 0 | 80.42 | | | Respiratory symptoms |
| Cross-sectional | 2 | | | | 24.89 | 0.72 | 859.33 | 85% | < 0.01 | Respiratory symptoms |
| Cross-sectional | 1 | | mRNA | | 14.68 | 1.37 | 157.53 | 91% | < 0.01 | Respiratory symptoms |

| | | | | | | | | | | |
|---|---|---|---|---|---|---|---|---|---|---|
| Cross-sectional | 1 | | Viralvector | | 8.12 | 0.97 | 68.03 | 42% | 0.14 | Respiratory symptoms |
| Cross-sectional | 2 | | mRNA | | 27.81 | 0.04 | 1000 | 87% | < 0.01 | Respiratory symptoms |
| Cross-sectional | 1 | | | | 10.86 | 3.43 | 34.44 | 89% | < 0.01 | Respiratory symptoms |
| Cross-sectional | 1 | Autoimmune | mRNA | | 9.20 | 0.00 | 1000 | 31% | 0.23 | Neurological symptoms |
| Cross-sectional | 2 | Autoimmune | | | 8.23 | 0.96 | 70.59 | 0 | 47% | Neurological symptoms |
| Cross-sectional | 1 | Autoimmune | Viralvector | | 42.19 | 0.00 | 1000 | 43% | 0.19 | Neurological symptoms |
| Cross-sectional | 1 | Autoimmune | | | 17.69 | 1.68 | 185.93 | 45% | 0.14 | Neurological symptoms |
| Cross-sectional | 2 | Autoimmune | mRNA | | 9.20 | 0.00 | 1000 | 31% | 0.23 | Neurological symptoms |
| Cross-sectional | 2 | Autoimmune | Viralvector | | 0.00 | 0.00 | 80.42 | | | Neurological symptoms |
| Cross-sectional | 1 | Autoimmune | Viralvector | 1.20 | | 0.00 | 14.60 | 0 | 0.51 | Lymphadenopathy |
| Cross-sectional | 2 | Autoimmune | | 0.57 | | 0.00 | 1.34 | 0 | 0.78 | Lymphadenopathy |
| Cross-sectional | 2 | Autoimmune | mRNA | 0.54 | | 0.00 | 3.38 | 0 | 0.55 | Lymphadenopathy |
| Cross-sectional | 1 | Autoimmune | | 0.15 | | 0.00 | 0.54 | 0 | 0.72 | Lymphadenopathy |
| Cross-sectional | 1 | Autoimmune | mRNA | 0.13 | | 0.00 | 1.74 | 0 | 0.50 | Lymphadenopathy |
| Cross-sectional | 2 | Autoimmune | Viralvector | 0.00 | | 0.00 | 4.29 | | | Lymphadenopathy |
| Cross-sectional | 2 | Autoimmune | | | 21.53 | 8.50 | 54.58 | 27% | <0.01 | Lymphadenopathy |
| Cross-sectional | 1 | Autoimmune | | | 19.51 | 6.49 | 58.68 | 25% | 0.25 | Lymphadenopathy |
| Cross-sectional | 1 | Autoimmune | mRNA | | 6.80 | 0.00 | 1000 | 63% | 0.10 | Lymphadenopathy |
| Cross-sectional | 1 | Autoimmune | Viralvector | | 42.19 | 0.00 | 1000 | 43% | 0.19 | Lymphadenopathy |
| Cross-sectional | 2 | Autoimmune | mRNA | | 9.20 | 0.00 | 1000 | 31% | 0.23 | Lymphadenopathy |
| Cross-sectional | 2 | Autoimmune | Viralvector | | 0.00 | 0.00 | 80.42 | | | Lymphadenopathy |
| Cross-sectional | 2 | Children/ Adolescents | mRNA | | 120.88 | 84.69 | 165.55 | | | Hospitalization |
| Cross-sectional | 1 | Children/ Adolescents | mRNA | | 53.69 | 31.00 | 85.73 | | | Hospitalization |
| Cross-sectional | 1 | MS* | | 4.32 | | 0.00 | 12.81 | | | Flare up |
| Cross-sectional | 1 | Autoimmune | mRNA | 4.02 | | 0.00 | 8.92 | 0 | 0.67 | Flare up |
| Cross-sectional | 2 | Autoimmune | mRNA | 1.61 | | 0.42 | 2.80 | | | Flare up |
| Cross-sectional | 1 | | | 4.09 | | 2.98 | 5.63 | | | Flare up |
| Cross-sectional | 1 | Autoimmune | | | 79.98 | 21.22 | 301.41 | 57% | 0.07 | Flare up |
| Cross-sectional | 2 | Autoimmune | | | 44.54 | 10.99 | 180.48 | 0 | 0.41 | Flare up |

| | | | | | | | | | | |
|---|---|---|---|---|---|---|---|---|---|---|
| Cross-sectional | 1 | | | | 73.44 | 26.52 | 203.37 | 52% | 0.08 | Flare up |
| Cross-sectional | 1 | MS* | | | 30.30 | 0.77 | 157.59 | | | Flare up |
| Cross-sectional | 2 | Autoimmune | mRNA | | 31.69 | 0.00 | 1000 | 38% | 0.21 | Flare up |
| Cross-sectional | 1 | | Viralvector | | 0.00 | 0.00 | 115.70 | | | Flare up |
| Cross-sectional | 2 | Autoimmune | Viralvector | | 0 | 0 | 459.26 | | | Flare up |
| Cross-sectional | 1 | | mRNA | | 91.00 | 15.32 | 540.69 | 62% | 0.07 | Flare up |
| Cross-sectional | 2 | Children/ Adolescents | mRNA | 227.11 | | 170.58 | 283.64 | | | Cardiac symptoms |
| Cross-sectional | 1 | Children/ Adolescents | mRNA | 184.56 | | 135.78 | 233.34 | | | Cardiac symptoms |
| Cross-sectional | 2 | Children/ Adolescents | mRNA | | 227.11 | 178.78 | 281.46 | | | Cardiac symptoms |
| Cross-sectional | 1 | Children/ Adolescents | mRNA | | 184.56 | 142.17 | 233.36 | | | Cardiac symptoms |
| Cross-sectional | 1 | Autoimmune | mRNA | 7.23 | | 0.51 | 102.50 | 40% | 0.17 | Cardiac event |
| Cross-sectional | 1 | Autoimmune | Viralvector | 43.30 | | 4.21 | 445.75 | 14% | 0.32 | Cardiac event |
| Cross-sectional | 1 | Autoimmune | | | 17.04 | 4.00 | 72.66 | 45% | 0.08 | Cardiac event |
| Cross-sectional | 2 | Autoimmune | | | 7.84 | 2.85 | 21.57 | 0 | 0.56 | Cardiac event |
| Cross-sectional | 2 | Autoimmune | | 0.39 | | 0.08 | 0.69 | 0 | 0.90 | Cardiac event |
| Cross-sectional | 2 | Autoimmune | mRNA | 0.36 | | 0 | 0.78 | 0 | 0.77 | Cardiac event |
| Cross-sectional | 2 | Autoimmune | Viralvector | 0 | | 0 | 4.29 | 0 | 100 | Cardiac event |
| Cross-sectional | 2 | Autoimmune | Viralvector | | 0 | 0 | 80.42 | | | Cardiac event |
| Cross-sectional | 2 | Autoimmune | mRNA | | 7.89 | 1.18 | 52.74 | 20% | 0.29 | Cardiac event |
| Cross-sectional | 2 | Autoimmune | | | 7.84 | 2.85 | 21.57 | 0 | 0.56 | Cardiac event |
| Cross-sectional | 1 | | | | 8.47 | 3.61 | 19.89 | 0 | 0.47 | Allergic reaction |
| Cross-sectional | 1 | | mRNA | | 0 | 0 | 46.78 | | | Allergic reaction |
| Cross-sectional | 1 | | Inactivated | | 0 | 0 | 17.83 | | | Allergic reaction |
| Cross-sectional | 2 | | mRNA | | 0 | 0 | 46.78 | | | Allergic reaction |
| Cross-sectional | 2 | | Inactivated | | 0 | 0 | 17.83 | | | Allergic reaction |
| Cross-sectional | 1 | Autoimmune | | | 9.93 | 4.77 | 18.19 | | | Allergic reaction |
| Cross-sectional | 2 | Autoimmune | | | 8.94 | 4.09 | 16.90 | | | Allergic reaction |
| Cross-sectional | 1 | Cancer | | | 2.91 | 0.48 | 17.67 | 0 | 0.77 | Allergic reaction |
| Cross-sectional | 2 | Cancer | | | 2.91 | 0.48 | 17.67 | 0 | 0.77 | Allergic reaction |

| | | | | | | | | | | | |
|---|---|---|---|---|---|---|---|---|---|---|---|
| Cross-sectional | 1 | MS* | | | 27.81 | | 15.62 | 40.00 | | | Accelerated allergic reaction |
| Cross-sectional | 2 | MS* | | | 13.91 | | 5.29 | 22.53 | | | Accelerated allergic reaction |
| Cross-sectional | Any | Autoimmune | | | 1.82 | | 0.81 | 2.83 | 0 | 0.80 | Accelerated allergic reaction |
| Cross-sectional | Any | | Viralvector | | 1.66 | | 0 | 19.72 | 0 | 0.36 | Accelerated allergic reaction |
| Cross-sectional | Any | | mRNA | | 1.59 | | 0 | 10.21 | 0 | 0.43 | Accelerated allergic reaction |
| Cross-sectional | 1 | Lactating | mRNA | | 1.28 | | 0.00 | 16.39 | 91% | <0.01 | Accelerated allergic reaction |
| Cross-sectional | 1 | | | | 2.35 | | 0.002 | 2267.10 | 96% | <0.01 | Accelerated allergic reaction |
| RCT | 1 | Cancer | mRNA | | | 9.19 | 3.83 | 22.08 | | | Thromboembolism |
| RCT | 1 | Autoimmune | Inactivated | | 2.14 | | 0.008 | 4.29 | 56% | 0.08 | Respiratory symptoms |
| RCT | 1 | Autoimmune | Inactivated | | | 73.29 | 31.95 | 168.08 | 0 | 0.43 | Respiratory symptoms |
| RCT | 2 | Autoimmune | Inactivated | | | 67.43 | 15.33 | 296.48 | 67% | 0.03 | Respiratory symptoms |
| RCT | 1 | Autoimmune | Inactivated | | | 47.63 | 0.00 | 1000 | 67% | 0.08 | Neurological symptoms |
| RCT | 2 | Autoimmune | Inactivated | | | 36.89 | 0.00 | 1000 | 32% | 0.22 | Neurological symptoms |
| RCT | 1 | Children/ Adolescents | mRNA | | | 200.32 | 184.75 | 216.61 | | | Lymphadenopathy |
| RCT | 2 | Children/ Adolescents | mRNA | | | 185.04 | 169.95 | 200.87 | | | Lymphadenopathy |
| RCT | any | Cancer | mRNA | 0.5 | | | 0.05 | 5.51 | | | Death |
| RCT | any | | | | 0.13 | | 0.03 | 0.53 | | | Cardiac event |
| RCT | any | | | | | 3.67 | 0.59 | 6.75 | | | Cardiac event |

IMC*: Immunocompromied

MS*: Multiple sclerosis

### Table S8.Non-Sever Outcomes

| Design | References | Dose | Population | vaccine | IR | IP | low | high | I2 | P | Outcome |
|---|---|---|---|---|---|---|---|---|---|---|---|
| RCT | 22.23 | 1 | | | 16.30 | | 2.21 | 120.14 | 0.99 | < 0.01 | Myalgia |
| RCT | 18,19,20,25 | 1 | | | 16.30 | | 2.21 | 120.14 | 0.99 | < 0.01 | Myalgia |
| RCT | 18,19,20,25 | 2 | | | 17.05 | | 2.77 | 104.89 | | | Myalgia |
| RCT | 18,20,25 | 1 | Children/ Adolescents | mRNA | 17.18 | | 0.00 | 58.77 | 1 | < 0.01 | Myalgia |
| RCT | 19 | 2 | IMC | Viralvector | 18.52 | | 4.80 | 32.24 | | | Myalgia |
| RCT | 18,20,25 | 2 | Children/ Adolescents | mRNA | 28.42 | | 0.00 | 106.87 | 1 | < 0.01 | Myalgia |
| RCT | 19 | 1 | IMC | Viralvector | 50.26 | | 27.66 | 72.87 | | | Myalgia |
| RCT | 22.23 | 2 | Autoimmune | Inactivated | | 79.36 | 4.41 | 1000.00 | 0 | 0.50 | Myalgia |
| RCT | 18,20,25 | 2 | Children/ Adolescents | mRNA | | 115.99 | 3.75 | 1000.00 | 1 | < 0.01 | Myalgia |
| RCT | 22,23,18,20,25 | 2 | | | | 104.98 | 38.55 | 285.93 | 0.99 | < 0.01 | Myalgia |
| RCT | 19 | 2 | IMC | Viralvector | | 129.63 | 53.74 | 249.01 | | | Myalgia |
| RCT | 22,23,18,20,25 | 1 | | | | 97.21 | 31.35 | 301.41 | 0.98 | < 0.01 | Myalgia |
| RCT | 22.23 | 1 | Autoimmune | Inactivated | | 72.13 | 0.42 | 1000.00 | 0.14 | 0.28 | Myalgia |
| RCT | 18,20,25 | 1 | Children/ Adolescents | mRNA | | 79.09 | 3.67 | 1000.00 | 0.99 | < 0.01 | Myalgia |
| RCT | 19 | 1 | IMC* | Viralvector | | 351.85 | 226.81 | 493.76 | | | Myalgia |
| RCT | 24 | any | Cancer | mRNA | 0.62 | | 0.00 | 2.03 | 0.06 | 0.30 | GI symptoms |
| RCT | 19 | 1 | IMC | Viralvector | 10.58 | | 0.21 | 20.95 | | | GI symptoms |
| RCT | 19 | 2 | IMC | Viralvector | 10.58 | | 0.21 | 20.95 | | | GI symptoms |
| RCT | 18,20,25 | 2 | Children/ Adolescents | mRNA | 12.12 | | 0.00 | 48.26 | 100 | < 0.01 | GI symptoms |
| RCT | 18,19,20,25 | 1 | | | 6.10 | | 6.10 | 41.19 | 0.95 | < 0.01 | GI symptoms |
| RCT | 18,19,20,25 | 2 | | | 6.53 | | 0.57 | 74.43 | 0.98 | < 0.01 | GI symptoms |
| RCT | 18,20,25 | 1 | Children/ Adolescents | mRNA | 7.93 | | 0.00 | 24.44 | 0.99 | < 0.01 | GI symptoms |
| RCT | 18,19,20,22,23,25 | 2 | | | | 51.32 | 21.65 | 121.62 | 0.96 | < 0.01 | GI symptoms |
| RCT | 18,19,20,22,23,25 | 1 | | | | 40.37 | 21.86 | 74.55 | 0.88 | < 0.01 | GI symptoms |
| RCT | 22.23 | 1 | Autoimmune | Inactivated | | 38.14 | 24.97 | 58.25 | 0 | 0.91 | GI symptoms |
| RCT | 18,20,25 | 1 | Children/ Adolescents | mRNA | | 35.98 | 1.14 | 1000 | 0.97 | < 0.01 | GI symptoms |
| RCT | 19 | 1 | IMC | Viralvector | | 74.07 | 20.55 | 178.93 | | | GI symptoms |
| RCT | 22.23 | 2 | Autoimmune | Inactivated | | 65.33 | 23.79 | 179.39 | 0.42 | 0.13 | GI symptoms |
| RCT | 18,20,25 | 2 | Children/ Adolescents | mRNA | | 38.82 | 0.42 | 1000 | 0.99 | < 0.01 | GI symptoms |
| RCT | 19 | 2 | IMC | Viralvector | | 74.07 | 20.55 | 178.93 | | | GI symptoms |

| | | | | | | | | | | | |
|---|---|---|---|---|---|---|---|---|---|---|---|
| RCT | 18,20,25 | 2 | Children/ Adolescents | mRNA | | 53.96 | 4.68 | 621.56 | 0.98 | < 0.01 | Fever |
| RCT | 19 | 2 | IMC | Viralvector | | 35.86 | 0.00 | 1000 | 0.29 | 0.23 | Fever |
| RCT | 22.23 | 2 | Autoimmune | Inactivated | | 36.89 | 0.00 | 1000 | 0.32 | 0.22 | Fever |
| RCT | 18,19,20,22,23,25 | 2 | | | | 47.20 | 20.84 | 106.90 | 0.94 | < 0.01 | Fever |
| RCT | 18,19,20,22,23,25 | 1 | | | | 37.10 | 14.26 | 96.52 | 0.89 | < 0.01 | Fever |
| RCT | 18,20,25 | 1 | Children/ Adolescents | mRNA | | 23.00 | 3.24 | 163.09 | 0.91 | < 0.01 | Fever |
| RCT | 19 | 1 | IMC | Viralvector | | 56.55 | 0.00 | 1000 | 0.77 | 0.04 | Fever |
| RCT | 22.23 | 1 | Autoimmune | Inactivated | | 49.37 | 3.13 | 777.75 | 0 | 0.62 | Fever |
| RCT | 19,20,25 | 2 | | | 25.70 | | 6.68 | 98.85 | 0.99 | < 0.01 | Fatigue |
| RCT | 19 | 2 | IMC | Viralvector | 30.86 | | 0.00 | 131.08 | 0.33 | 0.22 | Fatigue |
| RCT | 20.25 | 1 | Children/ Adolescents | mRNA | 31.84 | | 0.00 | 244.25 | 0.99 | < 0.01 | Fatigue |
| RCT | 20.25 | 2 | Children/ Adolescents | mRNA | 31.96 | | 0.00 | 332.68 | 1 | < 0.01 | Fatigue |
| RCT | 19,20,25 | 1 | | | 38.47 | | 13.57 | 109.11 | 0.98 | < 0.01 | Fatigue |
| RCT | 19 | 1 | IMC | Viralvector | 55.37 | | 0.00 | 171.45 | 0.12 | 0.29 | Fatigue |
| RCT | 18,20,25 | 2 | Children/ Adolescents | mRNA | | 246.38 | 10.55 | 1000.00 | 1 | < 0.01 | Fatigue |
| RCT | 19 | 2 | IMC | Viralvector | | 224.73 | 9.18 | 1000.00 | 0.46 | 0.17 | Fatigue |
| RCT | 18,20,25 | 2 | Autoimmune | Inactivated | | 77.37 | 42.03 | 142.39 | 0 | 0.62 | Fatigue |
| RCT | 18,19,20,22,23,25 | 2 | | | | 148.10 | 70.63 | 310.57 | 0.99 | < 0.01 | Fatigue |
| RCT | 18,19,20,22,23,25 | 1 | | | | 190.29 | 95.59 | 378.80 | 0.98 | < 0.01 | Fatigue |
| RCT | 18,20,25 | 1 | Children/ Adolescents | mRNA | | 259.56 | 36.50 | 1000.00 | 0.99 | < 0.01 | Fatigue |
| RCT | 19 | 1 | IMC | Viralvector | | 402.25 | 51.00 | 1000.00 | 0.46 | 0.17 | Fatigue |
| RCT | 18,20,25 | 1 | Autoimmune | Inactivated | | 99.70 | 48.81 | 203.65 | 0 | 0.39 | Fatigue |
| RCT | 22.23 | 1 | Autoimmune | Inactivated | | 19.23 | 18.96 | 19.49 | | | Conjuctivitis and Uveitis |
| RCT | 22.23 | 2 | Autoimmune | Inactivated | | 5.77 | 5.30 | 6.23 | | | Conjuctivitis and Uveitis |
| Cross-sectional | 39 | 1 | | Inactivated | 105.26 | | 21.04 | 189.49 | | | Myalgia |
| Cross-sectional | 46,51,53 | 1 | | mRNA | 12.69 | | 0.00 | 33.49 | 0.99 | < 0.01 | Myalgia |
| Cross-sectional | 33 | 1 | | Viralvector | 14.60 | | 0.00 | 36.13 | 0 | 0.32 | Myalgia |
| Cross-sectional | 33 | 1 | Autoimmune | | 14.60 | | 0.00 | 36.13 | 0 | 0.32 | Myalgia |
| Cross-sectional | 31 | 1 | MS* | | 183.58 | | 152.26 | 214.90 | | | Myalgia |
| Cross-sectional | 31 | 2 | MS | | 194.71 | | 162.46 | 226.97 | | | Myalgia |
| Cross-sectional | 31,33,39,46,51,53 | 1 | | | 24.64 | | 7.47 | 81.21 | 0.99 | < 0.01 | Myalgia |
| Cross-sectional | 29 | 2 | Autoimmune | | 59.78 | | 48.99 | 70.56 | | | Myalgia |

| | | | | | | | | | | | |
|---|---|---|---|---|---|---|---|---|---|---|---|
| Cross-sectional | 29,46,51,53 | 2 | | mRNA | 66.75 | | 34.72 | 98.77 | 0.99 | < 0.01 | Myalgia |
| Cross-sectional | 51.53 | 2 | Pregnant | | 68.74 | | 9.25 | 128.24 | 0.99 | < 0.01 | Myalgia |
| Cross-sectional | 29,46,51,53 | 2 | | | 80.35 | | 40.36 | 159.95 | 0.99 | < 0.01 | Myalgia |
| Cross-sectional | 28,39,44 | any | | | | 215.82 | 121.57 | 383.14 | 0.90 | < 0.01 | Myalgia |
| Cross-sectional | 28 | any | | Inactivated | | 113.33 | 67.42 | 175.25 | | | Myalgia |
| Cross-sectional | 28 | any | | Viralvector | | 286.46 | 223.66 | 355.98 | | | Myalgia |
| Cross-sectional | 28.44 | any | | mRNA | | 284.76 | 5.57 | 1000.00 | 0.78 | 0.03 | Myalgia |
| Cross-sectional | 28.44 | any | MS | | | 227.98 | 100.67 | 516.29 | 0.89 | < 0.01 | Myalgia |
| Cross-sectional | 39 | any | Autoimmune | | | 170.40 | 123.50 | 226.31 | | | Myalgia |
| Cross-sectional | 29,31,32,33,35,36,39,<br>44,46,51,53 | 2 | | | | 136.08 | 56.80 | 326.03 | 0.99 | 0 | Myalgia |
| Cross-sectional | 29,31,44,46,51,53 | 2 | | mRNA | | 220.42 | 86.59 | 561.13 | 0.99 | < 0.01 | Myalgia |
| Cross-sectional | 32 | 2 | | Inactivated | | 9.76 | 1.18 | 34.80 | | | Myalgia |
| Cross-sectional | 32 | 2 | Cancer | | | 35.34 | 6.37 | 1000.00 | 0.87 | < 0.01 | Myalgia |
| Cross-sectional | 29,35,44 | 2 | Autoimmune | | | 180.88 | 23.24 | 1407.55 | 0.99 | < 0.01 | Myalgia |
| Cross-sectional | 31 | 2 | MS | | | 194.71 | 166.38 | 225.58 | | | Myalgia |
| Cross-sectional | 51.53 | 2 | Pregnant | | | 464.42 | 197.06 | 1094.51 | 1 | < 0.01 | Myalgia |
| Cross-sectional | 31,33,35,36,44,46,51,<br>53 | 1 | | | | 59.91 | 37.09 | 96.76 | 0.96 | < 0.01 | Myalgia |
| Cross-sectional | 32 | 1 | Cancer | | | 28.60 | 6.53 | 125.27 | 0.82 | < 0.01 | Myalgia |
| Cross-sectional | 33,35,36,44 | 1 | Autoimmune | | | 54.73 | 28.48 | 105.16 | 0.94 | < 0.01 | Myalgia |
| Cross-sectional | 31 | 1 | MS | | | 183.59 | 155.94 | 213.86 | | | Myalgia |
| Cross-sectional | 51.53 | 1 | Pregnant | | | 78.97 | 11.35 | 549.30 | 0.99 | < 0.01 | Myalgia |
| Cross-sectional | 32,36,44,46,51,53 | 1 | | mRNA | | 61.03 | 31.75 | 117.32 | 0.97 | < 0.01 | Myalgia |
| Cross-sectional | 32,36,39 | 1 | | Inactivated | | 40.27 | 0.38 | 1000.00 | 0.94 | < 0.01 | Myalgia |
| Cross-sectional | 33.36 | 1 | | Viralvector | | 55.37 | 12.02 | 255.03 | 0.85 | < 0.01 | Myalgia |
| Cross-sectional | 38 | 1 | | Viralvector | 1.21 | | 0.00 | 14.60 | 0 | 0.51 | GI symptoms |
| Cross-sectional | 26 | 1 | Children/<br>Adolescents | | 140.93 | | 98.31 | 183.56 | | | GI symptoms |
| Cross-sectional | 26,30,37,51,53 | 1 | | mRNA | 19.02 | | 0.00 | 56.01 | 0.99 | < 0.01 | GI symptoms |
| Cross-sectional | 37 | 1 | Cancer | | 2.29 | | 0.59 | 3.99 | | | GI symptoms |
| Cross-sectional | 30.38 | 1 | Autoimmune | | 3.44 | | 0.00 | 8.63 | 0.74 | < 0.01 | GI symptoms |
| Cross-sectional | 51.53 | 1 | Pregnant | | 7.29 | | 0.00 | 18.44 | 0.97 | < 0.01 | GI symptoms |
| Cross-sectional | 26,30,31,37,38,51,53 | 1 | | | 7.60 | | 2.17 | 26.64 | 0.98 | < 0.01 | GI symptoms |
| Cross-sectional | 31 | 1 | MS | | 77.88 | | 57.48 | 98.28 | | | GI symptoms |

| | | | | | | | | | | |
|---|---|---|---|---|---|---|---|---|---|---|
| Cross-sectional | 28.49 | any | | mRNA | 72.95 | 17.44 | 305.13 | 0.96 | < 0.01 | GI symptoms |
| Cross-sectional | 49.52 | any | | Viralvector | 70.51 | 26.22 | 189.61 | 0.83 | < 0.01 | GI symptoms |
| Cross-sectional | 52 | any | | Inactivated | 20.30 | 2.45 | 168.36 | 0.93 | < 0.01 | GI symptoms |
| Cross-sectional | 28,34,42,49,52 | any | | | 50.43 | 27.70 | 91.79 | 0.92 | < 0.01 | GI symptoms |
| Cross-sectional | 49.52 | any | Autoimmune | | 80.86 | 28.50 | 229.40 | 0.95 | < 0.01 | GI symptoms |
| Cross-sectional | 28 | any | MS | | 36.08 | 15.19 | 85.67 | 0.84 | < 0.01 | GI symptoms |
| Cross-sectional | 34 | any | Disable | | 0.00 | 0.00 | 336.27 | | | GI symptoms |
| Cross-sectional | 42 | any | Lactating | | 7.52 | 0.19 | 41.18 | | | GI symptoms |
| Cross-sectional | 26,30,35,37,38,47,51,53,63 | 2 | | | 74.70 | 33.88 | 164.72 | 0.99 | < 0.01 | GI symptoms |
| Cross-sectional | 26,30,37,38,51,63 | 2 | | mRNA | 83.02 | 23.58 | 292.25 | 0.98 | < 0.01 | GI symptoms |
| Cross-sectional | 38 | 2 | | Viralvector | 0.00 | 0.00 | 80.42 | | | GI symptoms |
| Cross-sectional | 30,35,38,47 | 2 | Autoimmune | | 36.93 | 9.66 | 141.13 | 0.79 | < 0.01 | GI symptoms |
| Cross-sectional | 26 | 2 | Children/ Adolescents | | 285.71 | 232.88 | 343.27 | | | GI symptoms |
| Cross-sectional | 31 | 2 | MS | | 84.84 | 65.52 | 107.65 | | | GI symptoms |
| Cross-sectional | 37 | 2 | Cancer | | 26.79 | 5.56 | 76.29 | | | GI symptoms |
| Cross-sectional | 51.53 | 2 | Pregnant | | 181.08 | 27.94 | 1000 | 0.99 | < 0.01 | GI symptoms |
| Cross-sectional | 26,30,36,37,38,51,53 | 1 | | | 32.23 | 18.96 | 54.79 | 0.88 | < 0.01 | GI symptoms |
| Cross-sectional | 26,30,36,37,38,51,53 | 1 | | mRNA | 37.48 | 16.38 | 85.76 | 0.91 | < 0.01 | GI symptoms |
| Cross-sectional | 36.38 | 1 | | Viralvector | 12.57 | 3.50 | 45.13 | 0.16 | 0.31 | GI symptoms |
| Cross-sectional | 36 | 1 | | Inactivated | 8.89 | 1.08 | 31.74 | | | GI symptoms |
| Cross-sectional | 30,36,37,38,51,53 | 1 | Autoimmune | | 21.57 | 10.75 | 43.25 | 0.71 | < 0.01 | GI symptoms |
| Cross-sectional | 26 | 1 | Children/ Adolescents | | 140.94 | 103.50 | 185.70 | | | GI symptoms |
| Cross-sectional | 31 | 1 | MS | | 77.89 | 59.37 | 99.95 | | | GI symptoms |
| Cross-sectional | 37 | 1 | Cancer | | 32.11 | 13.01 | 65.04 | | | GI symptoms |
| Cross-sectional | 51.53 | 1 | Pregnant | | 49.56 | 8.72 | 281.73 | 0.96 | < 0.01 | GI symptoms |
| Cross-sectional | 28,34,39,44,45,49,52 | any | | | 120.98 | 67.98 | 215.32 | 0.95 | < 0.01 | Fever |
| Cross-sectional | 43.52 | any | | Inactivated | 24.60 | 0.01 | 1000.00 | 0.85 | < 0.01 | Fever |
| Cross-sectional | 44.49 | any | | mRNA | 199.88 | 57.65 | 693.05 | 0.98 | < 0.01 | Fever |
| Cross-sectional | 39,49,52 | any | Autoimmune | | 136.67 | 56.32 | 331.65 | 0.96 | < 0.01 | Fever |
| Cross-sectional | 34 | any | Disable | | 555.56 | 212.01 | 863.00 | | | Fever |
| Cross-sectional | 43 | any | IMC | | 46.51 | 22.53 | 83.87 | | | Fever |
| Cross-sectional | 28.45 | any | MS | | 96.58 | 20.62 | 452.40 | 0.92 | < 0.01 | Fever |

| | | | | | | | | | | | |
|---|---|---|---|---|---|---|---|---|---|---|---|
| Cross-sectional | 26,27,29,30,32,33,36, 38,44,51,53 | 2 | | | | 113.99 | 71.06 | 182.83 | 0.99 | 0 | Fever |
| Cross-sectional | 38 | 2 | | Viralvector | | 113.64 | 37.94 | 245.58 | | | Fever |
| Cross-sectional | 32 | 2 | | Inactivated | | 0.00 | 0.00 | 17.83 | | | Fever |
| Cross-sectional | 26 | 2 | Children/ Adolescents | | | 589.74 | 528.85 | 648.67 | | | Fever |
| Cross-sectional | 27 | 2 | IMC | | | 17.70 | 2.15 | 62.47 | | | Fever |
| Cross-sectional | 26,27,29,30,31,32,33, 35,36,38,44,47,51,53 | 2 | | mRNA | | 128.00 | 74.99 | 218.46 | 100 | 0 | Fever |
| Cross-sectional | 31 | 2 | MS | | | 184.98 | 157.24 | 215.33 | | | Fever |
| Cross-sectional | 29,30,33,35,36,38,44, 47 | 2 | Autoimmune | | | 122.76 | 69.55 | 216.68 | 0.94 | < 0.01 | Fever |
| Cross-sectional | 32.37 | 2 | Cancer | | | 41.27 | 3.78 | 451.09 | 0.83 | < 0.01 | Fever |
| Cross-sectional | 51.53 | 2 | Pregnant | | | 148.52 | 56.86 | 387.91 | 100 | 0 | Fever |
| Cross-sectional | 27,30,31,32,33,35,37, 36,38,44,47,51,53 | 1 | | | | 38.81 | 23.63 | 63.73 | 0.98 | 0 | Fever |
| Cross-sectional | 27,30,32,33,37,36,38, 44,51,53 | 1 | | mRNA | | 28.08 | 14.84 | 53.14 | 0.99 | < 0.01 | Fever |
| Cross-sectional | 32.36 | 1 | | Inactivated | | 7.51 | 0.07 | 830.40 | 0 | 0.52 | Fever |
| Cross-sectional | 27 | 1 | IMC | | | 8.85 | 0.22 | 48.32 | | | Fever |
| Cross-sectional | 33,36,38 | 1 | | Viralvector | | 83.71 | 15.32 | 457.21 | 0.86 | < 0.01 | Fever |
| Cross-sectional | 31 | 1 | MS | | | 147.43 | 122.31 | 175.48 | | | Fever |
| Cross-sectional | 30,33,35,36,38,44,47 | 1 | Autoimmune | | | 47.37 | 24.18 | 92.81 | 0.92 | < 0.01 | Fever |
| Cross-sectional | 32.37 | 1 | Cancer | | | 37.70 | 11.74 | 121.04 | 0.60 | 0.06 | Fever |
| Cross-sectional | 51.53 | 1 | Pregnant | | | 14.73 | 4.72 | 45.99 | 0.86 | < 0.01 | Fever |
| Cross-sectional | 26,29,30,38,46,51,53 | 2 | | mRNA | 120.63 | | 0.00 | 273.61 | 1 | 0 | Fatigue |
| Cross-sectional | 30,33,38 | 1 | Autoimmune | | 18.29 | | 0.00 | 26.45 | 0.90 | < 0.01 | Fatigue |
| Cross-sectional | 33.38 | 1 | | Viralvector | 20.40 | | 6.50 | 34.30 | 0.71 | 0.02 | Fatigue |
| Cross-sectional | 31 | 2 | MS | | 211.39 | | 0.00 | 608.91 | 0.85 | < 0.01 | Fatigue |
| Cross-sectional | 29,30,38,47, | 2 | Autoimmune | | 35.75 | | 0.00 | 73.56 | 0.98 | < 0.01 | Fatigue |
| Cross-sectional | 26,30,31,33,37,38,40, 46,47,51,53, | 1 | | | 36.20 | | 18.27 | 71.73 | 0.99 | 0 | Fatigue |
| Cross-sectional | 51.53 | 1 | Pregnant | | 44.84 | | 27.88 | 61.80 | 0.97 | < 0.01 | Fatigue |
| Cross-sectional | 26,29,30,31,38,46,47, 51,53 | 2 | | | 52.84 | | 22.68 | 123.11 | 0.99 | 0 | Fatigue |
| Cross-sectional | 26 | 1 | Children/ Adolescents | | 607.38 | | 518.90 | 695.87 | | | Fatigue |
| Cross-sectional | 26 | 2 | Children/ Adolescents | | 736.26 | | 634.48 | 838.05 | | | Fatigue |
| Cross-sectional | 26,30,37,38,,51,53 | 1 | | mRNA | 86.77 | | 0.00 | 233.44 | 0.99 | < 0.01 | Fatigue |
| Cross-sectional | 51.53 | 2 | Pregnant | | 96.29 | | 50.61 | 141.97 | 0.98 | < 0.01 | Fatigue |
| Cross-sectional | 28,34,39,43,45,49,52 | any | | | | 193.78 | 113.51 | 330.83 | 0.96 | < 0.01 | Fatigue |

| | | | | | | | | | | | |
|---|---|---|---|---|---|---|---|---|---|---|---|
| Cross-sectional | 39,49,52 | any | Autoimmune | | | 246.07 | 145.86 | 415.12 | 0.97 | < 0.01 | Fatigue |
| Cross-sectional | 28.45 | any | MS | | | 86.65 | 12.30 | 610.31 | 0.96 | < 0.01 | Fatigue |
| Cross-sectional | 34 | any | Disable | | | 666.67 | 299.30 | 925.15 | | | Fatigue |
| Cross-sectional | 43 | any | IMC | | | 134.88 | 92.23 | 187.94 | | | Fatigue |
| Cross-sectional | 28,45,49 | any | | mRNA | | 221.25 | 57.69 | 848.56 | 0.98 | < 0.01 | Fatigue |
| Cross-sectional | 28,49,52 | any | | Viralvector | | 207.07 | 57.01 | 752.13 | 0.93 | < 0.01 | Fatigue |
| Cross-sectional | 28,43,52 | any | | Inactivated | | 67.63 | 6.62 | 691.10 | 0.84 | < 0.01 | Fatigue |
| Cross-sectional | 26,30,31,33,36,38,40, 44,47,51,53 | 2 | | | | 250.47 | 143.70 | 436.56 | 0.99 | 0 | Fatigue |
| Cross-sectional | 26,30,36,38,44,51,53 | 2 | | mRNA | | 274.79 | 139.90 | 539.73 | 0.99 | < 0.01 | Fatigue |
| Cross-sectional | 33,36,38 | 2 | | Viralvector | | 22.73 | 0.58 | 120.24 | | | Fatigue |
| Cross-sectional | 27 | 2 | IMC | | | 61.95 | 25.27 | 123.47 | | | Fatigue |
| Cross-sectional | 30,33,36,38,44,47 | 2 | Autoimmune | | | 177.01 | 55.64 | 563.11 | 0.98 | < 0.01 | Fatigue |
| Cross-sectional | 26 | 2 | Children/ Adolescents | | | 736.26 | 679.77 | 787.55 | | | Fatigue |
| Cross-sectional | 31.4 | 2 | MS | | | 210.48 | 31.86 | 1000.00 | 0.88 | < 0.01 | Fatigue |
| Cross-sectional | 37 | 2 | Cancer | | | 205.79 | 0.19 | 1000.00 | 0.93 | < 0.01 | Fatigue |
| Cross-sectional | 51.53 | 2 | Pregnant | | | 663.28 | 412.71 | 1065.98 | 1 | < 0.01 | Fatigue |
| Cross-sectional | 26,30,31,33,36,38,40, 44,47,51,53 | 1 | | | | 124.02 | 78.19 | 196.72 | 0.97 | < 0.01 | Fatigue |
| Cross-sectional | 26,30,36,38,44,51,53 | 1 | | mRNA | | 129.45 | 64.54 | 259.62 | 0.98 | < 0.01 | Fatigue |
| Cross-sectional | 33,36,38 | 1 | | Viralvector | | 87.81 | 35.63 | 216.44 | 0.88 | < 0.01 | Fatigue |
| Cross-sectional | 36 | 1 | | Inactivated | | 13.33 | 2.76 | 38.47 | | | Fatigue |
| Cross-sectional | 26 | 1 | Children/ Adolescents | | | 607.38 | 549.42 | 663.19 | | | Fatigue |
| Cross-sectional | 31.4 | 1 | MS | | | 268.46 | 141.69 | 1000.00 | 0.93 | < 0.01 | Fatigue |
| Cross-sectional | 37 | 1 | Cancer | | | 138.64 | 0.24 | 80041.55 | 0.93 | < 0.01 | Fatigue |
| Cross-sectional | 51.53 | 1 | Pregnant | | | 311.38 | 211.62 | 458.19 | 0.98 | < 0.01 | Fatigue |
| Cross-sectional | 30,33,36,38,44,47 | 1 | Autoimmune | | | 79.20 | 42.14 | 148.82 | 0.96 | < 0.01 | Fatigue |
| Cross-sectional | 38 | 2 | Autoimmune | Viralvector | | 0 | 0 | 80.4199 | | | ENT symptoms |
| Cross-sectional | 38 | 2 | Autoimmune | mRNA | | 9.2004 | 0.0002 | 1000 | 0.31 | 0.23 | ENT symptoms |
| Cross-sectional | 38 | 1 | | Viralvector | | 42.1926 | 0 | 1000 | 0.43 | 0.19 | ENT symptoms |
| Cross-sectional | 38 | 1 | | mRNA | | 7.9453 | 0 | 1000 | 0.56 | 0.13 | ENT symptoms |
| Cross-sectional | 38 | 1 | Autoimmune | | | 17.0284 | 1.0348 | 280.2269 | 0.53 | 0.09 | ENT symptoms |
| Cross-sectional | 38 | 2 | Autoimmune | | | 8.2314 | 0.9598 | 70.5944 | 0 | 0.47 | ENT symptoms |
| Cross-sectional | 50 | 3 | IMC | mRNA | 1.2185 | | 1.0889 | 1.3635 | | | Arthralgia |

| | | | | | | | | | | | |
|---|---|---|---|---|---|---|---|---|---|---|---|
| Cross-sectional | 31 | 2 | MS | | 108.484 | | 84.409 | 132.559 | | | Arthralgia |
| Cross-sectional | 31 | 1 | MS | | 152.9902 | | 124.4001 | 181.5803 | | | Arthralgia |
| Cross-sectional | 37 | 1 | Cancer | | 4.9148 | | 2.4276 | 7.402 | | | Arthralgia |
| Cross-sectional | 29 | 2 | Autoimmune | | 49.1388 | | 39.36 | 58.9176 | | | Arthralgia |
| Cross-sectional | 33 | 1 | Autoimmune | Viralvector | 3.4559 | | 0 | 10.7664 | 0 | 0.49 | Arthralgia |
| Cross-sectional | 29 | 2 | | mRNA | 32.6893 | | 1.6523 | 63.7264 | 0.99 | < 0.01 | Arthralgia |
| Cross-sectional | 29.31 | 2 | | | 35.8149 | | 10.8019 | 118.7477 | 0.99 | < 0.01 | Arthralgia |
| Cross-sectional | 37 | 2 | Cancer | | 7.6531 | | 3.323 | 11.9831 | | | Arthralgia |
| Cross-sectional | 35 | 2 | Autoimmune | | | 127.9383 | 115.3193 | 141.4032 | | | Arthralgia |
| Cross-sectional | 35 | 1 | Autoimmune | | | 111.368 | 99.5151 | 124.1055 | | | Arthralgia |
| Cross-sectional | 52 | any | Autoimmune | | | 18.0204 | 4.12 | 78.8179 | 0 | 0.72 | Arthralgia |
| Cross-sectional | 51 | 2 | Pregnant | | 37.3339 | | 0 | 165.4497 | 1 | < 0.01 | Arthralgia |
| Cross-sectional | 51 | 1 | Pregnant | | 4.7292 | | 0 | 22.9159 | 0.98 | < 0.01 | Arthralgia |
| Cross-sectional | 37.51 | 1 | | mRNA | 4.7904 | | 1.2152 | 8.3656 | 0.96 | < 0.01 | Arthralgia |
| Cross-sectional | 31,33,37,51 | 1 | | | 7.5327 | | 1.5217 | 37.2862 | 1 | < 0.01 | Arthralgia |
| Cohort | 66,68,77,94,123 | 1 | Autoimmune | | | 126.13 | 62.00 | 256.60 | 0.97 | < 0.01 | Myalgia |
| Cohort | 60,82,83,84,102,109, 112,124 | 1 | Cancer | | | 51.87 | 33.88 | 79.41 | 0.86 | < 0.01 | Myalgia |
| Cohort | 74,76,78,85,92,108,122 | 1 | IMC | | | 96.61 | 31.89 | 292.69 | 0.93 | < 0.01 | Myalgia |
| Cohort | 123 | 1 | Autoimmune | | 36.33 | | 0.00 | 74.54 | 0.92 | < 0.01 | Myalgia |
| Cohort | 110,116,130 | 1 | Cancer | | 5.72 | | 0.00 | 12.76 | 0.84 | < 0.01 | Myalgia |
| Cohort | 69.73 | 1 | Lactating | | | 201.05 | 100.83 | 400.88 | 0.21 | 0.28 | Myalgia |
| Cohort | 73 | 1 | Pregnant | | | 23.81 | 2.90 | 83.37 | | | Myalgia |
| Cohort | 60,66,68,69,73,74,76 ,77,78,84,85,102,108, [illegible] | 1 | | mRNA | | 80.92 | 52.65 | 124.37 | 0.95 | < 0.01 | Myalgia |
| Cohort | 119 | 1 | Disable | | 3.57 | | 0.00 | 10.57 | | | Myalgia |
| Cohort | 74,76,92,108 | 1 | IMC | | 32.48 | | 0.00 | 71.26 | 0.94 | < 0.01 | Myalgia |
| Cohort | 74,76,92,108,119,123,130 | 1 | | mRNA | 23.86 | | 7.01 | 40.71 | 0.98 | < 0.01 | Myalgia |
| Cohort | 74,76,92,108,110,116,119,123,130 | 1 | | | 15.70 | | 7.64 | 32.25 | 0.96 | < 0.01 | Myalgia |
| Cohort | 60,66,68,69,73,74,76, 77,78,84,85,94,102, [illegible] | 1 | | | | 78.24 | 55.06 | 111.18 | 0.96 | < 0.01 | Myalgia |
| Cohort | 66,68,77,94,123 | 2 | Autoimmune | | | 190.94 | 87.66 | 415.90 | 0.99 | < 0.01 | Myalgia |
| Cohort | 60,82,83,84,102,109, 112,124 | 2 | Cancer | | | 76.13 | 39.49 | 146.75 | 0.97 | < 0.01 | Myalgia |
| Cohort | 123 | 2 | Autoimmune | | 63.51 | | 0.00 | 210.87 | 0.98 | < 0.01 | Myalgia |
| Cohort | 113,116,130 | 2 | Cancer | | 25.90 | | 0.00 | 79.61 | 0.97 | < 0.01 | Myalgia |

| | | | | | | | | | | | |
|---|---|---|---|---|---|---|---|---|---|---|---|
| Cohort | 58,74,76,78,85,92,108,122 | 2 | IMC | | | 139.04 | 64.90 | 297.86 | 0.93 | < 0.01 | Myalgia |
| Cohort | 69.73 | 2 | Lactating | | | 665.20 | 493.94 | 895.84 | 0.50 | 0.11 | Myalgia |
| Cohort | 59,60,66,68,69,73,74, 76,77,78,85,102,108, [illegible] | 2 | | mRNA | | 164.75 | 102.99 | 263.56 | 0.98 | < 0.01 | Myalgia |
| Cohort | 58,59,60,66,68,69,73, 74,76,77,78,85,94,102,1 [illegible] | 2 | | | | 138.85 | 92.26 | 208.98 | 0.98 | < 0.01 | Myalgia |
| Cohort | 99 | any | Autoimmune | | | 200.00 | 143.44 | 267.01 | | | Myalgia |
| Cohort | 84.13 | any | Cancer | | | 23.99 | 2.76 | 208.54 | 0 | 0.44 | Myalgia |
| Cohort | 95,101,108 | any | IMC | | | 104.84 | 2.14 | 1000.00 | 1 | < 0.01 | Myalgia |
| Cohort | 62 | any | Pregnant | | | 66.08 | 37.45 | 106.65 | | | Myalgia |
| Cohort | 62,84,95,101,108,130 | any | | | | 70.60 | 21.18 | 235.36 | 0.99 | < 0.01 | Myalgia |
| Cohort | 84,101,108,130 | any | | mRNA | | 53.91 | 3.31 | 878.77 | 0.98 | < 0.01 | Myalgia |
| Cohort | 95 | any | | Inactivated | | 47.72 | 40.73 | 55.51 | | | Myalgia |
| Cohort | 119 | 2 | Disable | | 3.57 | | 0.00 | 10.57 | | | Myalgia |
| Cohort | 58,74,76,92 | 2 | IMC | | 28.42 | | 1.30 | 55.55 | 0.64 | 0.04 | Myalgia |
| Cohort | 58,74,76,92,119,123, 130 | 2 | | mRNA | 33.48 | | 12.52 | 54.43 | 0.98 | < 0.01 | Myalgia |
| Cohort | 58,74,76,92,113,116, 119,123,130 | 2 | | | 24.05 | | 11.19 | 51.68 | 0.96 | < 0.01 | Myalgia |
| Cohort | 66,68,77,94,123 | 1 | Autoimmune | | | 34.96 | 15.93 | 76.73 | 0.94 | < 0.01 | GI symptoms |
| Cohort | 57,64,98,105,109,112,116,124,130 | 1 | Cancer | | | 19.21 | 11.61 | 31.76 | 0.67 | < 0.01 | GI symptoms |
| Cohort | 91 | any | Autoimmune | | | 12.60 | 9.58 | 16.26 | | | GI symptoms |
| Cohort | 123 | 2 | Autoimmune | | 26.03 | | 0.00 | 52.93 | 0.71 | 0.06 | GI symptoms |
| Cohort | 77.123 | 1 | Autoimmune | | 10.71 | | 0.00 | 23.72 | 0.98 | < 0.01 | GI symptoms |
| Cohort | 91 | any | Autoimmune | | 1.80 | | 1.34 | 2.26 | | | GI symptoms |
| Cohort | 72.13 | any | Cancer | | | 9.58 | 0.00 | 1000 | 0.89 | < 0.01 | GI symptoms |
| Cohort | 67,76,78,92,93,108, 121,122 | 1 | IMC | | | 39.24 | 16.42 | 93.75 | 0.73 | < 0.01 | GI symptoms |
| Cohort | 69.89 | 1 | Lactating | | | 31.19 | 21.54 | 45.17 | 0 | 0.99 | GI symptoms |
| Cohort | 98,110,116,130 | 1 | Cancer | | 3.69 | | 0.00 | 8.19 | 0.55 | 0.04 | GI symptoms |
| Cohort | 109.116 | 2 | Cancer | | 3.17 | | 1.24 | 5.10 | 0.70 | < 0.01 | GI symptoms |
| Cohort | 130 | any | Cancer | | 0.37 | | 0.00 | 0.89 | | | GI symptoms |
| Cohort | 121 | 1 | | Viralvector | | 58.11 | 0.00 | 1000 | 0.87 | < 0.01 | GI symptoms |
| Cohort | 57,67,94,124 | 1 | | Inactivated | | 30.98 | 10.36 | 92.62 | 0.91 | < 0.01 | GI symptoms |
| Cohort | 101.108 | any | IMC | | | 113.96 | 0.00 | 1000 | 0.80 | 0.03 | GI symptoms |
| Cohort | 76,92,93,121 | 2 | IMC | | 9.21 | | 3.58 | 14.84 | 0 | 0.49 | GI symptoms |
| Cohort | 76,78,108,92,93,121, 131 | 1 | IMC | | 6.45 | | 0.67 | 12.23 | 0.87 | < 0.01 | GI symptoms |

| | | | | | | | | | | | |
|---|---|---|---|---|---|---|---|---|---|---|---|
| Cohort | 64,66,69,77,92,93,98,105,108,109,123,130 | 1 | | mRNA | | 30.54 | 20.95 | 44.53 | 0.88 | < 0.01 | GI symptoms |
| Cohort | 57,64,66,67,69,77,92,93,94,98,105,108,109,1 | 1 | | | | 29.46 | 21.51 | 40.35 | 0.89 | < 0.01 | GI symptoms |
| Cohort | 96 | any | Pregnant | | | 48.19 | 13.29 | 118.82 | | | GI symptoms |
| Cohort | 66,68,77,94,123 | 2 | Autoimmune | | | 40.74 | 16.06 | 103.33 | 0.97 | < 0.01 | GI symptoms |
| Cohort | 57,64,98,105,109,112,116,124,130 | 2 | Cancer | | | 32.18 | 19.10 | 54.22 | 0.87 | < 0.01 | GI symptoms |
| Cohort | 67,76,78,92,93,108,121,122 | 2 | IMC | | | 53.72 | 25.98 | 111.07 | 0.75 | < 0.01 | GI symptoms |
| Cohort | 69.89 | 2 | Lactating | | | 50.24 | 21.64 | 116.68 | 0.48 | 0.05 | GI symptoms |
| Cohort | 64,66,69,77,92,93,98,105,108,109,123,130 | 2 | | mRNA | | 45.38 | 29.89 | 68.91 | 0.93 | < 0.01 | GI symptoms |
| Cohort | 57,64,66,67,69,77,92,93,94,98,105,108,109,1 | 2 | | | | 40.37 | 29.28 | 55.65 | 0.93 | < 0.01 | GI symptoms |
| Cohort | 57,67,94,124 | 2 | | Inactivated | | 32.85 | 14.30 | 75.47 | 0.92 | < 0.01 | GI symptoms |
| Cohort | 72,91,96,101,108,130 | any | | mRNA | | 24.06 | 5.99 | 96.59 | 0.99 | < 0.01 | GI symptoms |
| Cohort | 121 | 2 | | Viralvector | | 12.05 | 0.30 | 65.31 | | | GI symptoms |
| Cohort | 76,92,93,109,116,121,123 | 2 | | | 7.36 | | 4.05 | 13.56 | 0.96 | < 0.01 | GI symptoms |
| Cohort | 76,77,78,92,93,98,108,130,131 | 1 | | mRNA | 7.09 | | 3.09 | 11.10 | 0.96 | < 0.01 | GI symptoms |
| Cohort | 76,77,78,92,93,98,108,110,116,121,123,130, | 1 | | | 5.30 | | 3.00 | 9.35 | 0.85 | < 0.01 | GI symptoms |
| Cohort | 121 | 2 | | Viralvector | 4.02 | | 0.00 | 11.89 | | | GI symptoms |
| Cohort | 121.123 | 1 | | Viralvector | 16.03 | | 0.00 | 170.89 | 0.93 | < 0.01 | GI symptoms |
| Cohort | 76,92,93,109,123 | 2 | | mRNA | 12.70 | | 5.22 | 20.17 | 0.98 | < 0.01 | GI symptoms |
| Cohort | 91.13 | any | | | 1.11 | | 0.18 | 6.99 | 0.63 | 0.06 | GI symptoms |
| Cohort | 91.99 | any | Autoimmune | | | 86.68 | 4.82 | 1000.00 | 0.78 | 0.03 | Fever |
| Cohort | 66,77,94,111,121,123 | 1 | Autoimmune | | | 44.99 | 23.36 | 86.63 | 0.93 | < 0.01 | Fever |
| Cohort | 18 | 1 | Children/ Adolescents | | | 4.76 | 0.12 | 26.24 | | | Fever |
| Cohort | 83 | 2 | Children/ Adolescents | | | 33.33 | 13.50 | 67.47 | | | Fever |
| Cohort | 66,68,77,94,111,118,123 | 2 | Autoimmune | | | 117.23 | 63.33 | 217.04 | 0.98 | < 0.01 | Fever |
| Cohort | 57,59,60,64,82,98,105,102,,109,112,113,117,12 | 2 | Cancer | | | 81.27 | 50.22 | 131.52 | 0.95 | < 0.01 | Fever |
| Cohort | 64,82,84,98,105,102,109,110,112,113,116, | 1 | Cancer | | | 33.43 | 22.09 | 50.61 | 0.71 | < 0.01 | Fever |
| Cohort | 119 | 1 | Disable | | | 50.00 | 6.11 | 169.20 | | | Fever |
| Cohort | 119 | 2 | Disable | | | 100.00 | 27.93 | 236.64 | | | Fever |
| Cohort | 72,84,130, | any | Cancer | | | 14.33 | 6.68 | 30.73 | 0 | 0.62 | Fever |
| Cohort | 67,76,78,85,92,107,108,121,122 | 2 | IMC | | | 59.55 | 31.28 | 113.36 | 0.91 | < 0.01 | Fever |
| Cohort | 95,101,104,107 | any | IMC | | | 57.17 | 11.59 | 282.06 | 0.98 | < 0.01 | Fever |
| Cohort | 85 | 1 | IMC | | | 37.05 | 18.09 | 75.89 | 0.67 | < 0.01 | Fever |

| | | | | | | | | | | | |
|---|---|---|---|---|---|---|---|---|---|---|---|
| Cohort | 69,73,89 | 2 | Lactating | | | 645.60 | 414.21 | 1000.00 | 0.39 | 0.19 | Fever |
| Cohort | 69 | 1 | Lactating | | | 138.09 | 46.65 | 408.74 | 0 | 0.41 | Fever |
| Cohort | 3.96 | any | Pregnant | | | 42.01 | 8.75 | 201.74 | 0 | 0.71 | Fever |
| Cohort | 63.73 | 2 | Pregnant | | | 224.02 | 1.70 | 1000.00 | 0.62 | 0.10 | Fever |
| Cohort | 55 | 1 | MS | | | 19.82 | 9.93 | 35.19 | | | Fever |
| Cohort | 55 | 2 | MS | | | 119.54 | 90.58 | 153.79 | | | Fever |
| Cohort | 63.73 | 1 | Pregnant | | | 13.25 | 1.95 | 90.17 | 0 | 0.85 | Fever |
| Cohort | 72,84,91,101,104,107,130 | any | | mRNA | | 47.27 | 18.87 | 118.37 | 0.95 | < 0.01 | Fever |
| Cohort | 3,72,84,91,95,96,99, 101,104,107,130 | any | | | | 45.35 | 22.03 | 93.35 | 0.96 | < 0.01 | Fever |
| Cohort | 57,66,82,94,124 | 2 | | Inactivated | | 43.41 | 4.10 | 459.15 | 0.99 | < 0.01 | Fever |
| Cohort | 55,63,64,66,69,73,77, 82,84,85,94,98,105,102, [illegible] | 1 | | | | 39.04 | 29.88 | 51.00 | 0.84 | < 0.01 | Fever |
| Cohort | 55,63,64,66,69,73,77, 84,85,98,105,102,109,1 [illegible] | 1 | | mRNA | | 37.92 | 27.39 | 52.50 | 0.78 | < 0.01 | Fever |
| Cohort | 82,94,124 | 1 | | Inactivated | | 24.57 | 4.15 | 145.35 | 0 | 0.36 | Fever |
| Cohort | 55,59,60,63,64,67,69, 73,76,78,83,85,89,92, [illegible] | 2 | | mRNA | | 117.38 | 83.03 | 165.94 | 0.95 | < 0.01 | Fever |
| Cohort | 55,57,59,60,63,64,66, 67,69,73,76,78,82,83, [illegible] | 2 | | | | 101.29 | 74.77 | 137.23 | 0.96 | < 0.01 | Fever |
| Cohort | 117 | 2 | | Viralvector | | 26.79 | 5.56 | 76.29 | | | Fever |
| Cohort | 95 | any | | Inactivated | | 13.21 | 9.61 | 17.69 | | | Fever |
| Cohort | 58,66,68,74,94,111, 118,123 | 2 | Autoimmune | | | 198.50 | 95.47 | 412.71 | 0.99 | < 0.01 | Fatigue |
| Cohort | 91.99 | Any | Autoimmune | | | 174.56 | 0.81 | 1000.00 | 0.98 | < 0.01 | Fatigue |
| Cohort | 66,68,74,94,111,123 | 1 | Autoimmune | | | 136.04 | 46.41 | 398.74 | 0.99 | < 0.01 | Fatigue |
| Cohort | 91.99 | Any | Autoimmune | | 73.75 | | 0.00 | 821.92 | 0.97 | < 0.01 | Fatigue |
| Cohort | 58,74,111,123 | 2 | Autoimmune | | 68.70 | | 0.00 | 198.95 | 1 | < 0.01 | Fatigue |
| Cohort | 74,111,123 | 1 | Autoimmune | | 57.08 | | 0.00 | 117.69 | 0.99 | < 0.01 | Fatigue |
| Cohort | 83 | 2 | Children/ Adolescents | | | 85.71 | 51.59 | 132.08 | | | Fatigue |
| Cohort | 83 | 1 | Children/ Adolescents | | | 57.14 | 29.87 | 97.69 | | | Fatigue |
| Cohort | 64,82,102,109,110,112, 116,124,125,130 | 1 | Cancer | | | 77.82 | 45.95 | 131.77 | 0.89 | < 0.01 | Fatigue |
| Cohort | 57,64,82,87,102,109, 110,112,116,124,125, [illegible] | 2 | Cancer | | | 176.81 | 114.21 | 273.72 | 0.95 | < 0.01 | Fatigue |
| Cohort | 72.13 | Any | Cancer | | | 157.00 | 0.67 | 1000.00 | 0.97 | < 0.01 | Fatigue |
| Cohort | 109,113,116,130 | 2 | Cancer | | 29.13 | | 0.89 | 57.37 | 0.94 | < 0.01 | Fatigue |
| Cohort | 130 | Any | Cancer | | 14.62 | | 11.38 | 17.87 | | | Fatigue |
| Cohort | 110,116,130 | 1 | Cancer | | 14.35 | | 0.00 | 39.01 | 0.92 | < 0.01 | Fatigue |
| Cohort | 119 | 2 | Disable | | | 50.00 | 6.11 | 169.20 | | | Fatigue |

| | | | | | | | | | | | |
|---|---|---|---|---|---|---|---|---|---|---|---|
| Cohort | 119 | 1 | Disable | | | 25.00 | 0.63 | 131.59 | | | Fatigue |
| Cohort | 101.108 | Any | IMC | | | 73.48 | 0.11 | 1000.00 | 0.95 | < 0.01 | Fatigue |
| Cohort | 67,76,78,85,92,93,108,1 21,122,131 | 2 | IMC | | | 231.12 | 141.65 | 377.12 | 0.92 | < 0.01 | Fatigue |
| Cohort | 76,78,85,92,93,108, 121,122,131 | 1 | IMC | | | 157.67 | 83.78 | 296.75 | 0.92 | < 0.01 | Fatigue |
| Cohort | 76,93,114,121,131 | 2 | IMC | | 39.99 | | 17.88 | 62.11 | 0.89 | < 0.01 | Fatigue |
| Cohort | 76,78,92,93,108,121, 131 | 1 | IMC | | 30.66 | | 9.97 | 51.35 | 0.95 | < 0.01 | Fatigue |
| Cohort | 69,73,89 | 2 | Lactating | | | 700.59 | 484.59 | 1000.00 | 0.92 | < 0.01 | Fatigue |
| Cohort | 69.73 | 1 | Lactating | | | 359.25 | 154.90 | 833.21 | 0.55 | 0.08 | Fatigue |
| Cohort | 73 | 2 | Lactating | | 32.26 | | 15.36 | 49.16 | | | Fatigue |
| Cohort | 55 | 1 | MS | | | 91.89 | 69.19 | 119.05 | | | Fatigue |
| Cohort | 55 | 2 | MS | | | 158.62 | 125.56 | 196.40 | | | Fatigue |
| Cohort | 62.96 | Any | Pregnant | | | 86.15 | 0.00 | 1000.00 | 0.92 | < 0.01 | Fatigue |
| Cohort | 73 | 2 | Pregnant | | | 488.10 | 377.41 | 599.65 | | | Fatigue |
| Cohort | 73 | 1 | Pregnant | | | 142.86 | 76.05 | 236.25 | | | Fatigue |
| Cohort | 73 | 2 | Pregnant | | 34.86 | | 24.19 | 45.54 | | | Fatigue |
| Cohort | 82,94,124 | 1 | | Inactivated | | 85.69 | 53.37 | 137.59 | 0.73 | 0.01 | Fatigue |
| Cohort | 72,91,96,101,108,130 | Any | | mRNA | | 84.19 | 16.11 | 439.93 | 0.99 | < 0.01 | Fatigue |
| Cohort | 121,123,125 | 2 | | Viralvector | | 58.92 | 0.00 | 1000.00 | 0.87 | < 0.01 | Fatigue |
| Cohort | 55,58,64,66,67,68,69, 73,74,76,78,83,85,89, [illegible] | 2 | | mRNA | | 268.91 | 193.95 | 372.85 | 0.99 | 0 | Fatigue |
| Cohort | 55,57,58,64,66,67,68, 69,73,74,76,78,82,83, [illegible] | 2 | | | | 223.37 | 171.69 | 290.61 | 0.99 | 0 | Fatigue |
| Cohort | 121,123,125 | 1 | | Viralvector | | 154.31 | 0.76 | 1000.00 | 0.88 | < 0.01 | Fatigue |
| Cohort | 55,64,66,68,69,73,74, 76,78,83,85,92,93,102,1 [illegible] | 1 | | mRNA | | 141.42 | 99.30 | 201.39 | 0.98 | < 0.01 | Fatigue |
| Cohort | 62,72,91,96,99,101, 108,130 | Any | | | | 128.51 | 33.78 | 488.88 | 1 | 0 | Fatigue |
| Cohort | 55,64,66,68,69,73,74, 76,78,82,83,85,92,93, [illegible] | 1 | | | | 119.23 | 85.73 | 165.81 | 0.98 | 0 | Fatigue |
| Cohort | 57,67,82,94,124 | 2 | | Inactivated | | 102.85 | 62.82 | 168.37 | 0.92 | < 0.01 | Fatigue |
| Cohort | 121.123 | 1 | | Viralvector | 49.11 | | 0.00 | 626.75 | 0.99 | < 0.01 | Fatigue |
| Cohort | 58,74,76,93,109,111, 123,130,131, | 2 | | mRNA | 43.32 | | 24.73 | 61.90 | 0.99 | < 0.01 | Fatigue |
| Cohort | 58,74,76,93,109,111, 113,114,116,121,123, [illegible] | 2 | | | 33.06 | | 22.23 | 49.17 | 0.98 | < 0.01 | Fatigue |
| Cohort | 74,76,78,92,93,108, 111,130,131 | 1 | | mRNA | 32.72 | | 15.60 | 49.83 | 0.99 | < 0.01 | Fatigue |
| Cohort | 91,99,130 | Any | | | 31.73 | | 1.44 | 699.64 | 0.99 | < 0.01 | Fatigue |
| Cohort | 114.121 | 2 | | Viralvector | 26.51 | | 0.00 | 328.54 | 0.93 | < 0.01 | Fatigue |
| Cohort | 74,76,78,92,93,108, 110,111,116,121,123, [illegible] | 1 | | | 22.78 | | 12.48 | 41.59 | 0.96 | < 0.01 | Fatigue |

| | | | | | | | | | | | |
|---|---|---|---|---|---|---|---|---|---|---|---|
| Cohort | 91.13 | Any | | mRNA | 16.19 | | 7.61 | 24.76 | 0.05 | 0.30 | Fatigue |
| Cohort | 68,77,94 | 2 | Autoimmune | | | 28.399 | 5.8367 | 138.1776 | 0.9 | P < 0.01 | ENT symptoms |
| Cohort | 68,77,94 | 1 | Autoimmune | | | 24.5251 | 4.6256 | 130.033 | 0.89 | P < 0.01 | ENT symptoms |
| Cohort | 67.92 | 2 | IMC | | | 40.0876 | 12.3355 | 130.2752 | 0 | 0.73 | ENT symptoms |
| Cohort | 67.94 | 2 | | Inactivated | | 88.9019 | 58.7648 | 134.4948 | 0.14 | 0.31 | ENT symptoms |
| Cohort | 92 | 3 | IMC | mRNA | | 32.7868 | 8.1996 | 131.0993 | | | ENT symptoms |
| Cohort | 92 | 1 | IMC | | | 16.3934 | 0.415 | 87.9881 | | | ENT symptoms |
| Cohort | 67,68,77,92,94 | 2 | | | | 31.1798 | 11.896 | 81.723 | 0.85 | P < 0.01 | ENT symptoms |
| Cohort | 68,77,92,94 | 1 | | | | 24.3842 | 6.385 | 93.1226 | 0.87 | P < 0.01 | ENT symptoms |
| Cohort | 68,77,92 | 2 | | mRNA | | 20.0768 | 5.0703 | 79.4982 | 0.73 | P < 0.01 | ENT symptoms |
| Cohort | 68,77,92 | 1 | | mRNA | | 13.1154 | 2.4773 | 69.4366 | 0.64 | 0.02 | ENT symptoms |
| Cohort | 94 | 1 | | Inactivated | | 82.4176 | 65.3761 | 102.2173 | | | ENT symptoms |
| Cohort | 68 | 1 | Autoimmune | mRNA | | 1.4577 | 0.0369 | 8.0949 | | | Conjuctivitis and Uveitis |
| Cohort | 68 | 2 | | mRNA | | 2.9155 | 0.3533 | 10.4916 | | | Conjuctivitis and Uveitis |
| Cohort | 125 | 1 | | Viralvector | 0 | | 0 | 89.8049 | | | Arthralgia |
| Cohort | 66,77,111.123 | 1 | Autoimmune | | 13.0062 | | 0 | 26.8834 | 0.97 | < 0.01 | Arthralgia |
| Cohort | 58,69,76,85,109,111, 123 | 2 | | mRNA | 14.2072 | | 2.1629 | 26.2514 | 0.99 | < 0.01 | Arthralgia |
| Cohort | 69 | 2 | Lactating | | 14.5936 | | 9.854 | 19.3333 | 0 | 0.9 | Arthralgia |
| Cohort | 58,111,123 | 2 | Autoimmune | | 29.1958 | | 0 | 76.6425 | 0.99 | < 0.01 | Arthralgia |
| Cohort | 76,109,116 | 2 | Cancer | | 3.3593 | | 0.3763 | 6.3423 | 0.92 | < 0.01 | Arthralgia |
| Cohort | 69 | 1 | Lactating | | 3.9374 | | 2.8786 | 4.9963 | 0 | 0.97 | Arthralgia |
| Cohort | 76,84,85,109,108,110,1 23,125 | 1 | | | 5.1923 | | 3.0696 | 8.7827 | 0.96 | < 0.01 | Arthralgia |
| Cohort | 76,84,109,110,116,125 | 1 | Cancer | | 5.62565 239 | | 0 | 11.6937 | 0.85 | < 0.01 | Arthralgia |
| Cohort | 76,84,85,109,108,110,1 23 | 1 | | mRNA | 7.1216 | | 2.4753 | 11.7679 | 0.96 | < 0.01 | Arthralgia |
| Cohort | 58,69,76,85,109,111, 116,123 | 2 | | | 7.4803 | | 2.8422 | 19.6869 | 0.98 | < 0.01 | Arthralgia |
| Cohort | 123 | 1 | | Recombina nt | 37.2671 | | 25.9964 | 48.5377 | | | Arthralgia |
| Cohort | 66,68,77,94 | 2 | Autoimmune | | | 80.7067 | 44.2359 | 147.2461 | 0.68 | 0.02 | Arthralgia |
| Cohort | 68.94 | 1 | Autoimmune | | | 68.1685 | 0.0097 | 1000 | 0.97 | < 0.01 | Arthralgia |
| Cohort | 112.122 | 1 | Cancer | | | 54.8959 | 30.3908 | 99.1602 | 0 | 0.79 | Arthralgia |
| Cohort | 108,112,122 | 2 | Cancer | | | 153.826 4 | 69.5531 | 340.2085 | 0.69 | 0.02 | Arthralgia |
| Cohort | 66,68,77,108,122 | 2 | | mRNA | | 91.8697 | 37.6889 | 223.94 | 0.93 | < 0.01 | Arthralgia |
| Cohort | 68,94,112,122 | 1 | | | | 63.8125 | 29.8589 | 136.3759 | 0.91 | < 0.01 | Arthralgia |

| | | | | | | | | | | |
|---|---|---|---|---|---|---|---|---|---|---|
| Cohort | 62,90,108 | any | | | 50.4877 | 5.8293 | 437.2722 | 0.98 | < 0.01 | Arthralgia |
| Cohort | 90 | any | | Inactivated | 37.4449 | 21.9616 | 59.2774 | | | Arthralgia |
| Cohort | 108 | any | | mRNA | 343.75 | 289.0207 | 401.7328 | | | Arthralgia |
| Cohort | 66,68,77,94,108,112,122 | 2 | | | 105.2421 | 66.7934 | 165.8234 | 0.89 | < 0.01 | Arthralgia |
| Cohort | 94 | 2 | | Inactivated | 102.1978 | 83.2808 | 123.7333 | | | Arthralgia |
| Cohort | 94 | 1 | | Inactivated | 1.6896 | 1.391 | 1.9881 | | | Arthralgia |
| Cohort | 68.122 | 1 | | mRNA | 1.2224 | 0 | 3.7894 | 0 | 0.41 | Arthralgia |
| Case-Control | 133 | 2 | Autoimmune | Inactivated | 23.22 | 6.25 | 86.20 | 0 | 0.57 | Fever |
| Case-Control | 134 | 1 | Pregnant | mRNA | 15.38 | 5.67 | 33.18 | | | Fever |
| Case-Control | 134 | 2 | Pregnant | mRNA | 89.74 | 63.31 | 122.60 | | | Fever |
| Case-Control | 133 | 1 | Autoimmune | Inactivated | 111.46 | 87.93 | 138.72 | | | Fatigue |
| Case-Control | 133 | 2 | Autoimmune | Inactivated | 84.39 | 63.85 | 108.94 | | | Fatigue |
| Case-Control | 133 | 2 | Autoimmune | | 28.488 | 0.6189 | 1000 | 0.88 | < 0.01 | Conjuctivitis and Uveitis |
| Case-Control | 133 | 1 | Autoimmune | | 21.1482 | 9.1698 | 48.774 | 0 | 0.56 | Conjuctivitis and Uveitis |

IMC*: Immunocompromied

MS*: Multiple sclerosis

### References


1. World Health O. WHO SAGE roadmap for prioritizing uses of COVID-19 vaccines in the context of limited supply: an approach to inform planning and subsequent recommendations based on epidemiological setting and vaccine supply scenarios, first issued 20 October 2020, latest update 16 July 2021. CC BY-NC-SA 3.0 IGO. Geneva: World Health Organization, 2021.

144. Akhtar Z, Trent M, Moa A, Tan TC, Fröbert O, MacIntyre CR. The impact of COVID-19 and COVID vaccination on cardiovascular outcomes. *Eur Heart J Suppl* 2023; 25(Suppl A): A42-a9.

145. Hause AM, Marquez P, Zhang B, et al. COVID-19 mRNA Vaccine Safety Among Children Aged 6 Months-5 Years - United States, June 18, 2022-August 21, 2022. *MMWR Morb Mortal Wkly Rep* 2022; 71(35): 1115-20.

146. Mascolo A, di Mauro G, Fraenza F, et al. Maternal, fetal and neonatal outcomes among pregnant women receiving COVID-19 vaccination: The preg-co-vax study. *Front Immunol* 2022; 13: 965171.

147. Kang L, Shang W, Gao P, Wang Y, Liu J, Liu M. Immunogenicity and Safety of COVID-19 Vaccines among People Living with HIV: A Systematic Review and Meta-Analysis. *Vaccines (Basel)* 2022; 10(9).

148. Li HJ, Yang QC, Yao YY, et al. COVID-19 vaccination effectiveness and safety in vulnerable populations: a meta-analysis of 33 observational studies. *Front Pharmacol* 2023; 14: 1144824.
