## Supplemental figures for "COVID-19 Vaccine Safety Studies among Vulnerable Populations: A Systematic Review and Meta-analysis of 120 Observational Studies and Randomized Clinical Trials": supplementary Figures.docx

**Figure S1. Proportion of Studies Per Country**

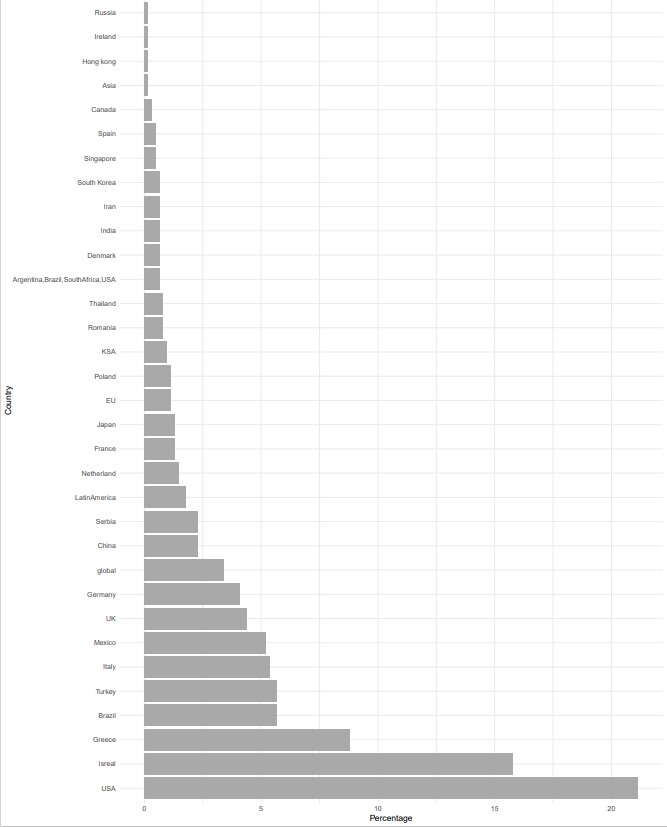

**Figure S2. Funnel Plot for Observational Studies**

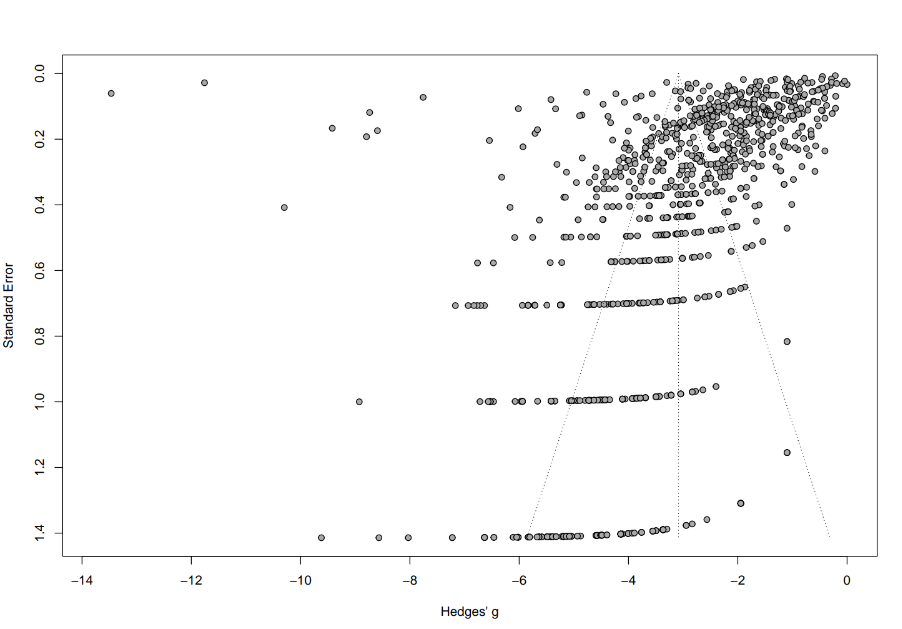

**Figure S3. Funnel Plot for Randomized Clinical Trials**

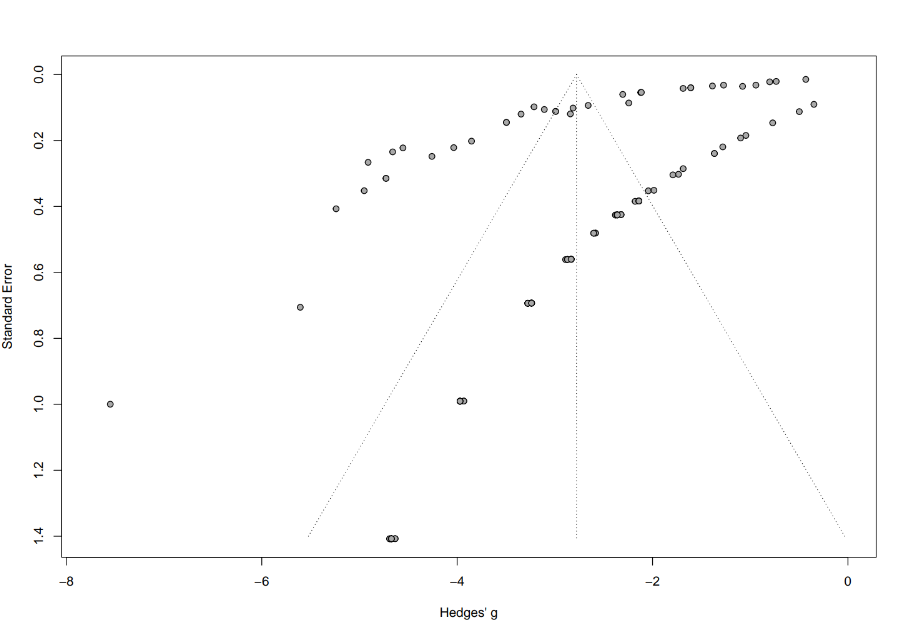
